# Multi-omics in the All Of Us Research Program expands mapping genetic mechanisms in health and disease

**DOI:** 10.64898/2026.10.29.26366507

**Authors:** Evin M. Padhi, Elizabeth Kiernan, Jonathan Nguyen, Marianne K. DeGorter, Victoria Rosa, Yulia Mostovoy, Alex Yenkin, Hang Su, Lee Lichtenstein, Andrew R. Marderstein, Josh Gillard, Wail Baalawi, Sophie Parsa, Mine S. Cicek, Jeffrey G. Meyer, Ashley L. Blegen, Victoria A. McNally, Jolene J. Lane, Zachary S. Fredricksen, All of Us Research Program Multi-omics Working Group, The All of Us Research Program, Namrata Gupta, Harrison Brand, Kiran Garimella, Konrad J. Karczewski, Michael E. Talkowski, Niall Lennon, Stephen B. Montgomery

## Abstract

The All of Us (AoU) Research Program multi-omics cohort was developed to expand discovery of the molecular mechanisms underlying health and disease. Here, we report on the AoU multi-omics cohort comprising proteomic (N = 10,056) and transcriptomic (N = 9,130) sequencing data generated from ancestrally diverse AoU participants. We provide comprehensive expression, splicing, and protein quantitative trait locus (e/s/pQTL) analyses and detected 182,246 QTLs, of which 54,255 finemapped to rare variants and 59.4% (32,235) resolve to a single causal variant. We provide multiple insights into rare variant effects and demonstrate that they are more likely to be ancestrally-restricted, replicate across molecular layers and impact known Mendelian disease genes (OR = 1.4) and approved drug targets (OR = 1.3). Rare variant QTLs from AoU further provided evidence for 5,186 variants, many absent from ClinVar, affecting established disease genes. Combined, the availability of multi-omics in AoU provides a key resource to identify the genetic and molecular mechanisms impacting health and disease across multiple ancestry groups in the United States.

## Introduction

A major challenge in human genomics is mapping the molecular effects of genetic variation (*1*, *2*). Previous studies have associated genetic variants to molecular phenotypes, such as gene expression, splicing, and protein-levels, to uncover functional effects impacting the majority of human genes (*3–9*). However, increasingly large studies have indicated that discovery of independent effects and rarer variant effects are far from saturated (*8*, *10*). To overcome this, existing molecular reference datasets have traditionally encountered a trade-off between scale and diversity: large cohorts have been disproportionately European-ancestry (*5*, *10*), whereas more ancestrally diverse cohorts have often been too small to provide adequate power to identify rare variants effects (*6*, *7*). As a consequence, much of the molecular landscape of rare genetic variation has remained uncharacterized, especially within diverse ancestry groups, limiting both biological discovery and clinical use.

Here, we report on biobank-scale genome sequencing and multi-omic profiling from 10,000 ancestrally diverse participants in the All of Us (AoU) Research Program (*11*) to deeply characterize the molecular effects of both common and rare genetic variation. We observe that ancestral diversity of AoU multi-omics substantially improves fine-mapping variants to a single likely-causal variant. The AoU multi-omics cohort also provides an unprecedented opportunity to study molecular effects across both transcriptome and proteome. Using this design, we reveal a vast repertoire of rare variants with large molecular effects that propagate across multi-omic layers. Further, we observe that rare variant effects were predominantly discovered in a single ancestry group and contained a subset of thousands of uncharacterized variants in well-established disease genes. We further show that variant effects can combine to produce dosage-like effects in every individual’s genome. Together, these findings uncover a previously inaccessible set of rare and ancestry-enriched molecular effects and demonstrate the essential role of biobank-scale multi-omics to enable their discovery and interpretation in health and disease.

## Results

### Ancestral diversity improves fine-mapping of molecular trait loci

The AoU multi-omics cohort was designed to maximize representation of diverse computed genetic ancestry groups within the United States (N = 10,056; Fig. 1A) (Supplemental Table S1), providing an unprecedented opportunity to characterize the molecular effects of human genetic variation. Whole-genome sequencing and plasma proteomics were generated for all participants, with total RNA sequencing available for 9,130 individuals. The use of total RNA sequencing substantially expanded quantification of noncoding transcripts and genes involved in transcriptional regulation relative to poly(A)-selected whole-blood datasets (*10*) (Supplementary Figure 1). After quality control, 8,895 participants with total RNA-sequencing data and 9,581 participants with plasma proteomic data were retained for QTL mapping (Methods; Fig. 1A). The use of Olink Explore HT quantified 5,415 circulating proteins with high precision (Methods) (Supplemental Figure 2), with strongest coverage among extracellular and secreted proteins (Supplementary Fig. 3). Together, these complementary molecular measurements provide a foundation for systematic characterization of molecular traits across several diverse ancestry groups.

**Figure 1.**
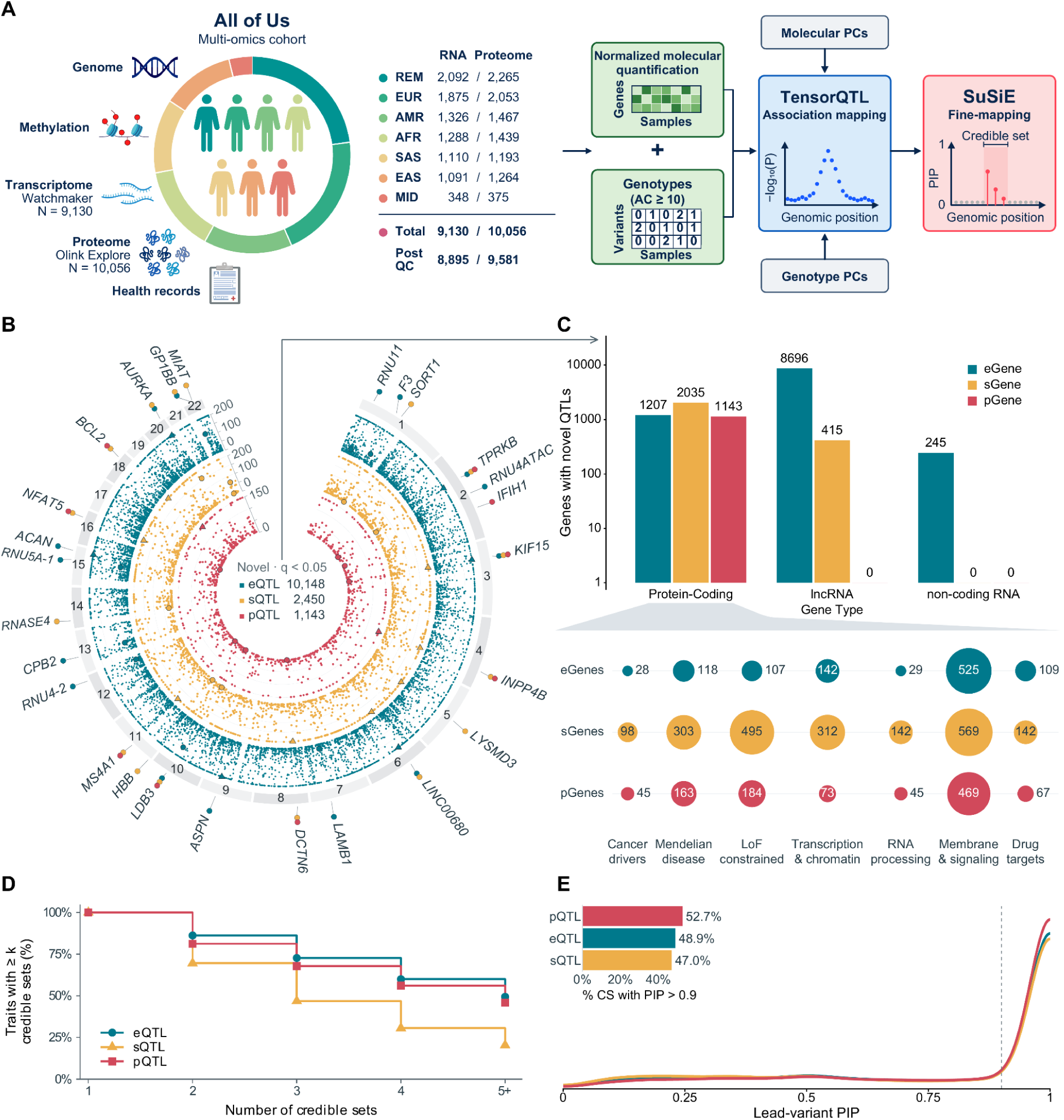
Multi-omic cis-QTL discovery and fine-mapping in the All of Us Research Program. **A.** Overview of the multi-omics cohort and workflow for mapping expression, splicing, and protein quantitative trait loci (eQTLs, sQTLs, and pQTLs). Participant counts are shown as RNA/proteome, with totals of 9,130 and 10,056, respectively. Predicted ancestry groups are AFR (African-like), AMR (Admixed American-like), EAS (East Asian-like), EUR (European-like), MID (Middle Eastern-like), SAS (South Asian-like), and REM (not belonging to one of the other ancestries or is an admixture). Normalized molecular phenotypes and genotypes with allele count ≥10 were used for tensorQTL association mapping, with molecular and genotype principal components as covariates, followed by SuSiE fine-mapping. **B.** Genome-wide distribution of novel significant cis-QTL associations (*q* < 0.05). The outer, middle, and inner tracks show eQTLs, sQTLs, and pQTLs, respectively. Points represent lead cis-variants, positioned by genomic coordinate and nominal −log₁₀(*P*). Selected genes are labelled, and colored dots beside each label identify the QTL types with novel associations at that gene. Outlined markers highlight selected associations; triangles indicate highlighted associations exceeding the plotted radial limits. **C.** Numbers of novel significant QTL target genes by GENCODE v48 biotype. Novel genes have combined-analysis *q* < 0.05 and are absent from the significant reference gene sets for the corresponding assay: GTEx whole blood v10, INTERVAL, and TOPMed for eQTLs; GTEx whole blood v10 and INTERVAL for sQTLs; and UK Biobank Pharma Proteomics Project for pQTLs. Bubble area and labels indicate unique gene counts among cancer drivers, ClinGen Mendelian disease genes, loss-of-function-constrained genes, and transcription/chromatin regulators. ClinGen membership requires Moderate, Strong, or Definitive evidence; constraint requires unflagged gnomAD v2.1.1 LOEUF <0.35. The regulation category combines transcription factors, chromatin remodelers, and histone/DNA modification factors. **D.** Percentage of molecular traits with at least *k* credible sets in the combined analysis. Denominators are 29,088 expression genes, 5,763 individual splicing traits, and 2,529 protein assay targets.The final point, labelled 5+, represents traits with at least five credible sets. **E.** Distributions of lead-variant posterior inclusion probability (PIP), defined as the maximum variant PIP within each credible set, in the combined analysis. The dashed line marks PIP = 0.9. The inset shows the percentage of credible sets containing a variant with PIP >0.9.

We mapped expression (e), splicing (s), and protein (p) quantitative trait loci (QTL) across all samples (minor allele frequency; MAF > 0.05%) and within individual genetic similarity groups (Methods) (Supplemental Tables 2-4), identifying eQTLs, sQTLs, and pQTLs for 96.8%, 100%, and 58.7% of genes, respectively (Supplemental Figure 4). We observed that 93.8% of QTLs had reproducible effects in at least three computed genetic ancestry groups (Methods) (Supplemental Figure 5) and strongly replicated QTLs from external efforts (*4*, *8*, *10*) (Supplemental Figure 6-7). Lower pQTL discovery rates were predominantly attributable to protein detection rates (Supplemental Figure 8). Comparison against existing molecular reference resources such as GTEx, TOPMed, INTERVAL, and UK Biobank (*4*, *5*, *8*, *10*) demonstrated that the AoU QTLs expand the catalog of molecular trait associations, identifying 10,148, 2,450, and 1,143 previously unreported expression, splicing, and proteomics QTLs, respectively (Fig. 1B). Notably, 88% of novel eGenes and 16.9% of novel sGenes corresponded to lncRNAs and other noncoding RNAs (Fig 1C), indicating the increased sensitivity of total RNA sequencing relative to poly(A)-selected protocols used in most studies (Supplementary Figure 1, Supplementary Figure 9-10). Of the novel protein coding QTLs we identified, 159 were cancer drivers, 541 were Mendelian disease genes, 741 were genes under strong loss of function constraint, 475 were transcription and chromatin regulators, 199 were involved in RNA processing and splicing and 304 were drug targets (Fig 1C).

We performed statistical fine-mapping using SuSiE (*12*) in population-specific and combined analyses (Methods). Across all three QTL classes, the combined analysis identified more credible sets per molecular trait than the population-specific analyses (Supplemental Fig. S11), and achieved better fine-mapping resolution, with smaller credible sets and higher lead-variant posterior inclusion probabilities (PIPs) (Supplemental Figures S12–S13). The combined analysis identified 143,512, 17,674, and 11,922 credible sets for expression, splicing, and protein abundance, respectively, with 41–132% more eQTL credible sets than two other large blood eQTL studies when each dataset was restricted to protein-coding genes (Supplemental Figure S14). Molecular QTLs exhibited extensive allelic heterogeneity, with nearly half of eGenes harboring at least five independent eQTL credible sets (Fig. 1D; Supplemental Table 5). Across molecular phenotypes, 48.9%, 47%, and 52.7% of eQTL, sQTL, and pQTL credible sets, respectively, contained a variant with PIP > 0.9 (Fig. 1E; Supplemental Figure S13), yielding 84,831 high-confidence putative causal variants. Despite its smaller sample size, AoU also produced smaller pQTL credible sets than UK Biobank (mean sizes: 5.1 versus 9.7; (*5*); Supplemental Figure S12). Fine-mapped variants were enriched for genomic features consistent with their expected molecular mechanisms (Supplemental Figure S15; Supplemental Table 6). eQTLs were strongly enriched within promoter and enhancer elements as well as splice regions. sQTLs showed pronounced enrichment at splice donor and acceptor sites, whereas pQTLs were enriched for protein-coding variants and other coding-disrupting annotations.

### Biobank-scale multi-omics reveals the landscape of rare molecular effects

Multi-omics in a large, biobank-scale cohort enabled more systematic characterization of increasingly rare variation. Of 30,719,753 variants with MAF > 0.05% we identified 47,572 unique rare fine-mapped variants (max ancestry group MAF <1%) across eQTLs, sQTLs and pQTLs (Supplemental Table 7). When restricted to single variant credible sets with high posterior support (PIP > 0.9) we identified 9,523 rare variants, which represent 15.7% of all putative causal variants across the molecular phenotypes (Fig 2A).

**Figure 2.**
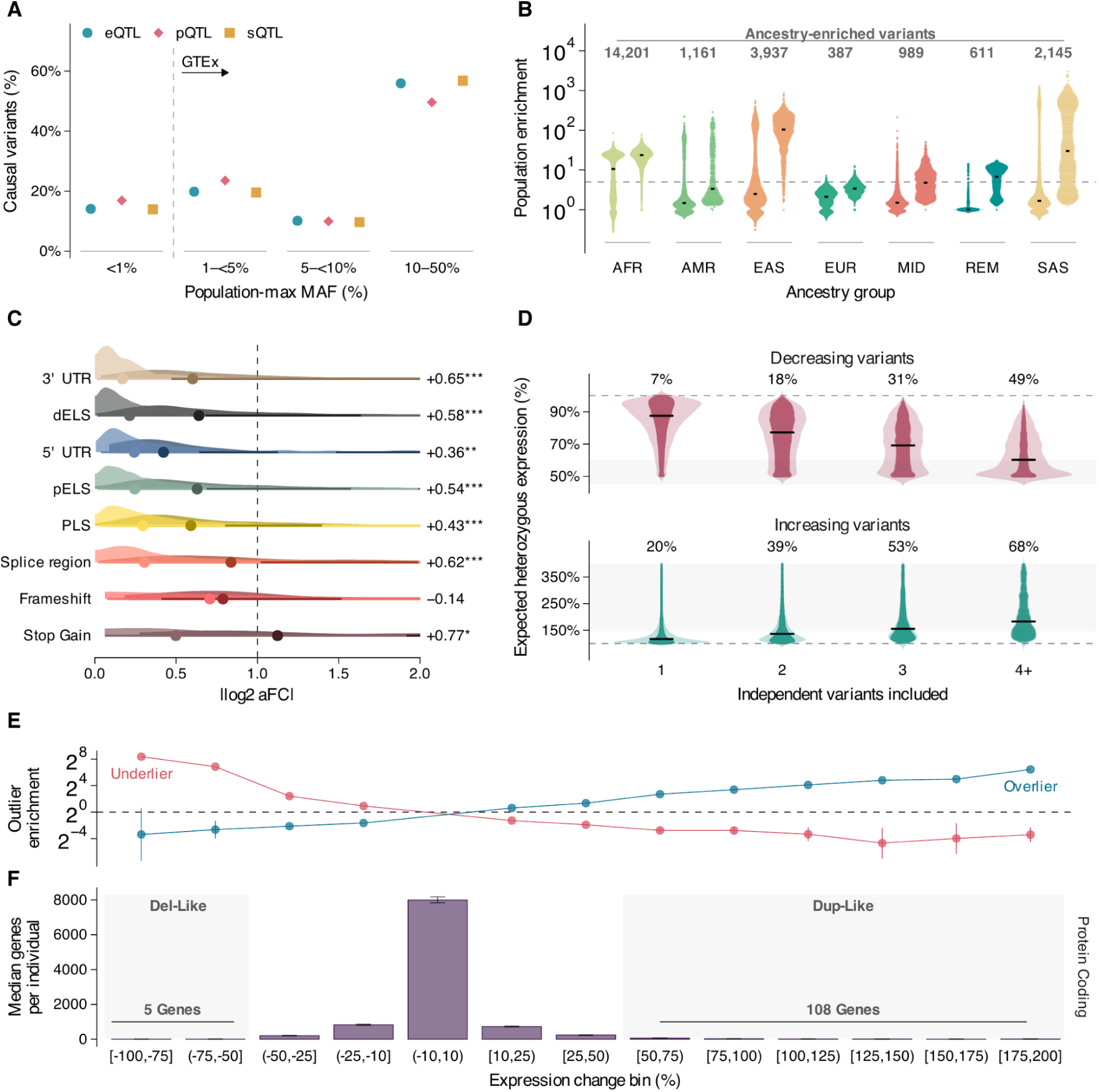
Allele frequencies, ancestry enrichment, and cumulative expression effects of molecular quantitative trait loci. **A.** Percentages of expression (eQTL), protein (pQTL), and splicing (sQTL) variant–trait pairs with PIP > 0.9 across population-maximum allele-frequency bins: <1%, 1–<5%, 5–<10%, and 10–50%. Pairs The dashed divider marks 1% allele frequency. **B.** Population-enrichment odds ratios (ORs) for 67,568 distinct high-confidence variants pooled across assays. Variants are grouped by the ancestry in which their allele frequency is highest. Paired distributions show common (population-maximum allele frequency ≥1%; left) and rare (<1%; right) variants. Points represent variants, violins show densities, and black bars mark medians. The dashed line marks OR = 5. Counts indicate ancestry-enriched variants (OR >5 and Benjamini–Hochberg false discovery rate [FDR] <0.05). **C.** Absolute log₂ allelic fold change (aFC) for 21,000 high-confidence eQTL variants, grouped by functional annotation. Dark and light shades indicate rare and common variants, respectively. Points mark medians; thick and thin intervals show central 66% and 95% quantile ranges. The display is limited to 0–2; the dashed line marks a twofold allelic effect. Right-hand labels give rare-minus-common mean differences from linear regression. Asterisks denote FDR-adjusted *P*values: *P <0.05, **P <0.01, and ***P <0.001. UTR, untranslated region; PLS, promoter-like signature; pELS, proximal enhancer-like signature; dELS, distal enhancer-like signature **D.** Predicted heterozygous expression after sequentially combining expression-decreasing or expression-increasing QTL effects, ordered by decreasing magnitude. Each point represents a gene at a cumulative step; 4+ pools steps with at least four variants. Black bars mark medians, and dashed lines indicate 100% reference expression. Percentages give the fractions of displayed observations reaching ≤60% or ≥150% expression, respectively. Shading marks 45–60% and 145–400% reference expression. **E.** Enrichment of observed low-expression outliers (underliers, red) and high-expression outliers (overliers, blue) across predicted expression-change bins from QTL burden. Points show ORs and vertical bars show 95% Wald confidence intervals. The dashed line marks OR = 1. The central (−10%, 10%) bin is used as the reference. **F.** Median numbers of protein-coding genes per individual in the corresponding predicted expression-change bins; error bars show interquartile ranges. Shading marks deletion-like (−100% to −50%) and duplication-like (50–200%) ranges. The displayed bin medians sum to 5 and 108 genes within these ranges, respectively.

Rare variants and, likewise rare functional variants are typically enriched within specific computed genetic similarity groups (*13*). The AoU multi-omics cohort provided a unique opportunity to identify rare functional variants that were enriched within a similarity group. To examine this, we compared group enrichment between rare and common variants within our combined fine-mapped analyses across QTL modalities (Methods). As expected, group-enrichment of QTLs increased dramatically with decreasing allele frequency: 78% of variants with global allele frequency below 0.5% were group-enriched, compared to 16% of variants with global allele frequency greater than 1% (Supplemental Figure 16). The AFR-like similarity group contributed the largest fraction of group-enriched variants (44.7%), followed by EAS-like (22.2%) and SAS-like (15.5%) ancestries (Methods). Quantifying the magnitude of this enrichment further demonstrated that rare causal QTL variants showed greater ancestry enrichment than common variants, with a mean difference in enrichment odds ratio of 39.6 after adjusting for ancestry (95% CI, 38.1–41.0; P < 2.2 × 10⁻¹⁶). The level of enrichment varied dramatically across the similarity groups with the greatest enrichment in our cohort seen in the rare QTL variants in the EAS-like (median rare variant enrichment OR = 104) and SAS-like similarity groups (mean rare variant enrichment OR = 29.9) (Fig 2B). Rare QTL variants also showed large allelic effects (|log2 aFC|) across all genomic contexts with the majority of the large effect variants being found in promoter-like regions, proximal and distal enhancers and coding disrupting variants (stop-gain, frameshift) (Fig 2C) (Supplemental Table 8).

### Cumulative genetic effects drive extreme molecular phenotypes in individual genomes

The presence of multiple independent eQTLs enabled high resolution analysis of how regulatory effects combine within individual genomes. Using an additive model, we calculated the expected heterozygous expression level resulting from increasing numbers of independent eQTLs affecting the same gene (Methods). Under this system, increasing the number of directionally consistent expression-decreasing alleles produced progressively larger cumulative effects, with the proportion of genes whose modeled expression reached a deletion-like range rising from 19%, when only the strongest allele was considered, to 72% when four or more alleles were considered (Fig. 2D) (Supplemental Table 9). Similarly, for expression-increasing alleles, the proportion of genes whose cumulative effects reached a duplication-like range increased from 34% when one allele was considered to 77% when four or more alleles were considered. Notably, these dosage-like expression states were markedly less prevalent when only common variants were considered, indicating the disproportionately large effects of rare variants and their contribution to cumulative molecular burden (Supplemental Figure 17). These results show that relatively small numbers of SNVs can combine to produce expression changes comparable in magnitude to those caused by gene deletions or duplications.

To quantify the impact of cumulative burden at the individual level, we estimated predicted gene expression using allelic fold changes (*14*) from high-confidence causal variants (PIP > 0.9) (Methods). Across all protein-coding genes, individuals carried a median of five genes with predicted expression decreases greater than 50% and 108 genes with predicted expression increases greater than 150% (Fig 2E) (Supplemental Table 10). Further, we directly observed that individuals with expected dosage-like effects were enriched among expression and protein underliers (Fig 2F; Supplemental Figure S18).

Genes that are under strong selective constraint were significantly depleted among the largest predicted perturbations in both deletions and duplications, with 18.9-fold and 30.7-fold reductions (Methods), respectively (*p* = 3.79 × 10⁻⁴ and *p* = 0.0033; Supplemental Figure S19). ClinGen annotations were also significantly depleted in both deletions and duplications (1.4-fold and 1.5-fold reductions, respectively; *p* = 3.95 × 10⁻⁴ and *p* = 0.0017). Burden and drug-interaction genes were significantly depleted only among deletions (1.4-fold and 1.2-fold reductions, respectively; *p* = 0.0149 and *p* = 0.0480). Despite this relative depletion, we identified thousands of strong cumulative QTL effects involving constrained genes.

### Coordinated effects of rare variants across the regulatory cascade

The cumulative effects observed at the level of gene expression suggested that rare variants may perturb multiple stages of gene regulation. We therefore tested for coordinated effects across expression, splicing, and protein abundance. To characterize the propagation of cis-regulatory effects across molecular layers (expression, splicing and protein), we performed pairwise colocalization of fine-mapped molecular QTLs (*15*). Across 755 genes with detectable eQTLs, sQTLs, and pQTLs (Supplemental Tables 11-12), 86.2% had at least one regulatory effect shared across molecular layers (Fig. 3A). Cross-omic colocalization rates varied across molecular phenotype pairs, ranging from 39.4% to 60.2% (Supplemental Fig S21), and approximately 25% of assayed proteins showed evidence of colocalization with blood eQTL or sQTL signals, substantially higher than previous estimates of transcript–protein sharing (*16*). These patterns were robust across a range of colocalization probability thresholds (Supplemental Figure S20).

**Figure 3.**
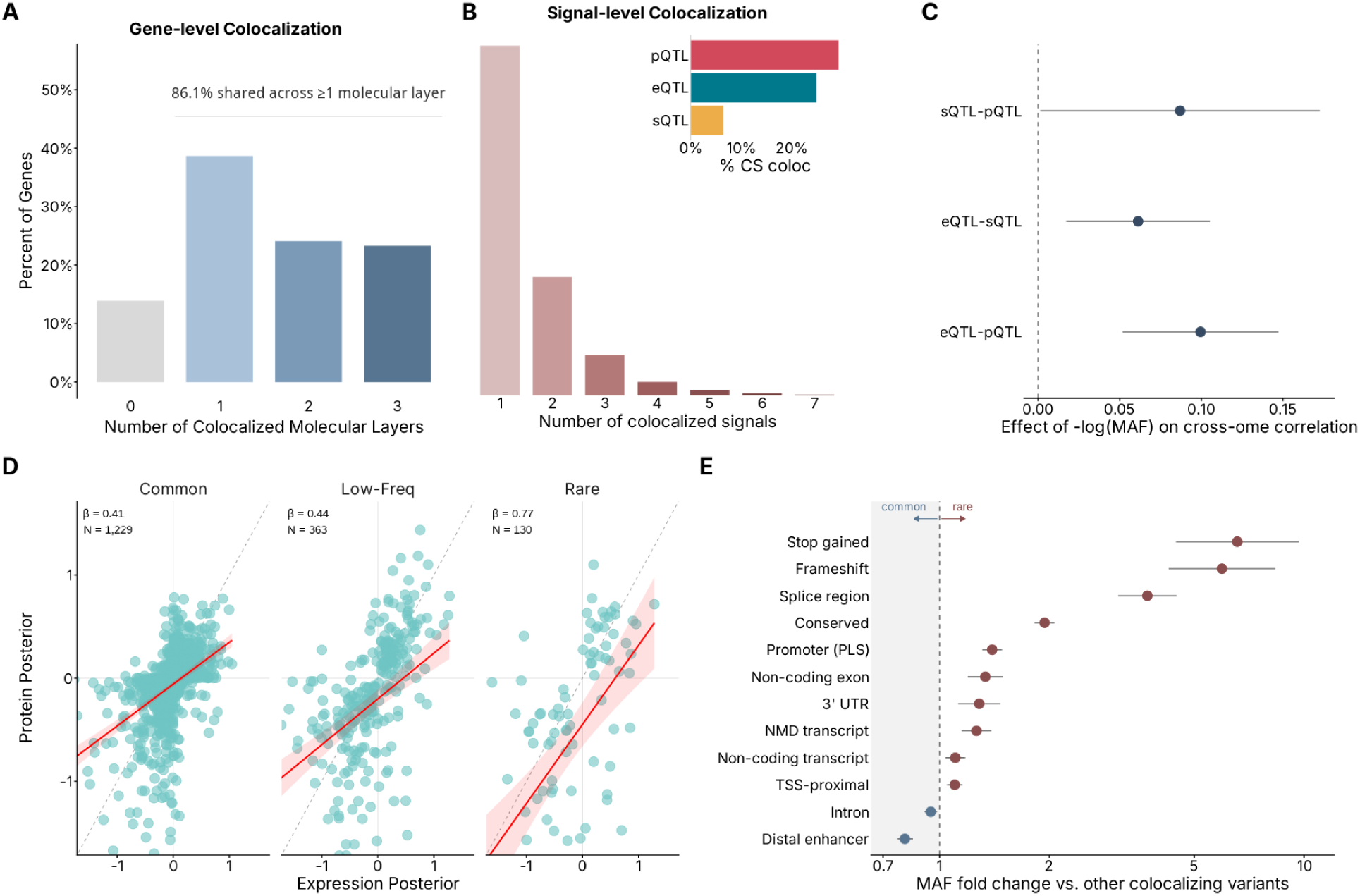
Frequency-dependent sharing of regulatory signals across molecular layers. **A.** For every gene fine-mapped in all three assays (n = 755), the number of assays pairs showing a gene-level colocalization is shown on the x axis, and the percentage of genes in that category on the y axis. **B.** Same as in A) but for credible set colocalization. Inset shows the percentage of total credible sets for each QTL type that has a colocalization **C.** Dependence of cross layer effect-size concordance on allele frequency. For each assay pair, points give the interaction between the two assays fine-mapped effect size and allele frequency across colocalized variants, with 95% confidence intervals **D.** Concordance of fine-mapped variant effect sizes between pQTLs and eQTLs, stratified by allele frequency. Each point is a colocalized eQTL-pQTL variant. **E.** Allele-frequency shift associated with each functional annotation among colocalized variants between eQTL and pQTLs. Points give the fold difference in minor allele frequency between variants belonging to that annotation group and other colocalizing variants, with 95% confidence intervals.

Despite extensive sharing found at the gene level, sharing between specific credible sets was limited. We identified 7,132 colocalized credible sets, representing 13% of all possible overlapping credible sets. Colocalization rates also differed across molecular phenotype pairs, with sQTL–pQTL signals showing the highest sharing rate (50.6%), compared with eQTL–pQTL (16.8%) and eQTL–sQTL (12.0%) signals (Supplemental Figure S22). Consistent with this, most genes contained only one or two colocalized signals despite frequently containing multiple independent QTL credible sets (Fig. 3B).

We next evaluated features that distinguish effects that propagate across the molecular layers. Allele frequency was a major driver of cross-omic sharing with colocalization rates between eQTL-pQTL and sQTL-pQTL both increasing markedly as allele frequency decreased; the most pronounced effect was seen for the sQTL-pQTL signals (Supplemental Figure S23). Consistent with this, the molecular effects of the colocalizing variants became increasingly concordant across the assays as allele frequency decreased and effect size increased (Fig 3C,D). Rare colocalizing variants were also enriched for coding and splice disrupting annotations (frameshift, stop-gained, splice region variants) whereas common colocalizing variants were comparatively enriched in distal enhancer elements (Fig 3E) (Supplemental Table 13). Altogether this suggests that rare variants are more likely to give rise to effects that propagate across the molecular layers.

### Fine-mapped molecular QTLs reveal convergent trait associations and improve effector-gene prioritization

To investigate the phenotypic consequences of fine-mapped QTL variants, we first integrated our molecular QTL fine-mapping with fine-mapped GWAS for 102 complex traits (Supplemental Table 14). Colocalization identified 9,222 gene–trait associations across many physiological systems (Fig. 4A).The molecular signatures of these colocalizations varied across trait categories. eQTLs provided the broadest coverage, with an average of 25–30% of GWAS (Supplemental Table 15) credible sets per trait colocalizing with an eQTL for neurodegenerative, dermatological, autoimmune, and allergic-respiratory traits. pQTL colocalization rates were highest for autoimmune and cardiometabolic traits (8%) and neurodegenerative traits (7%), whereas sQTL rates were highest for neurodegenerative traits (7%), followed by allergic-respiratory and autoimmune traits (5% each). Both pQTLs and sQTLs provided nonredundant information relative to eQTLs: 68% of pQTL and 54% of sQTL gene–trait associations lacked a corresponding eQTL association.

**Figure 4.**
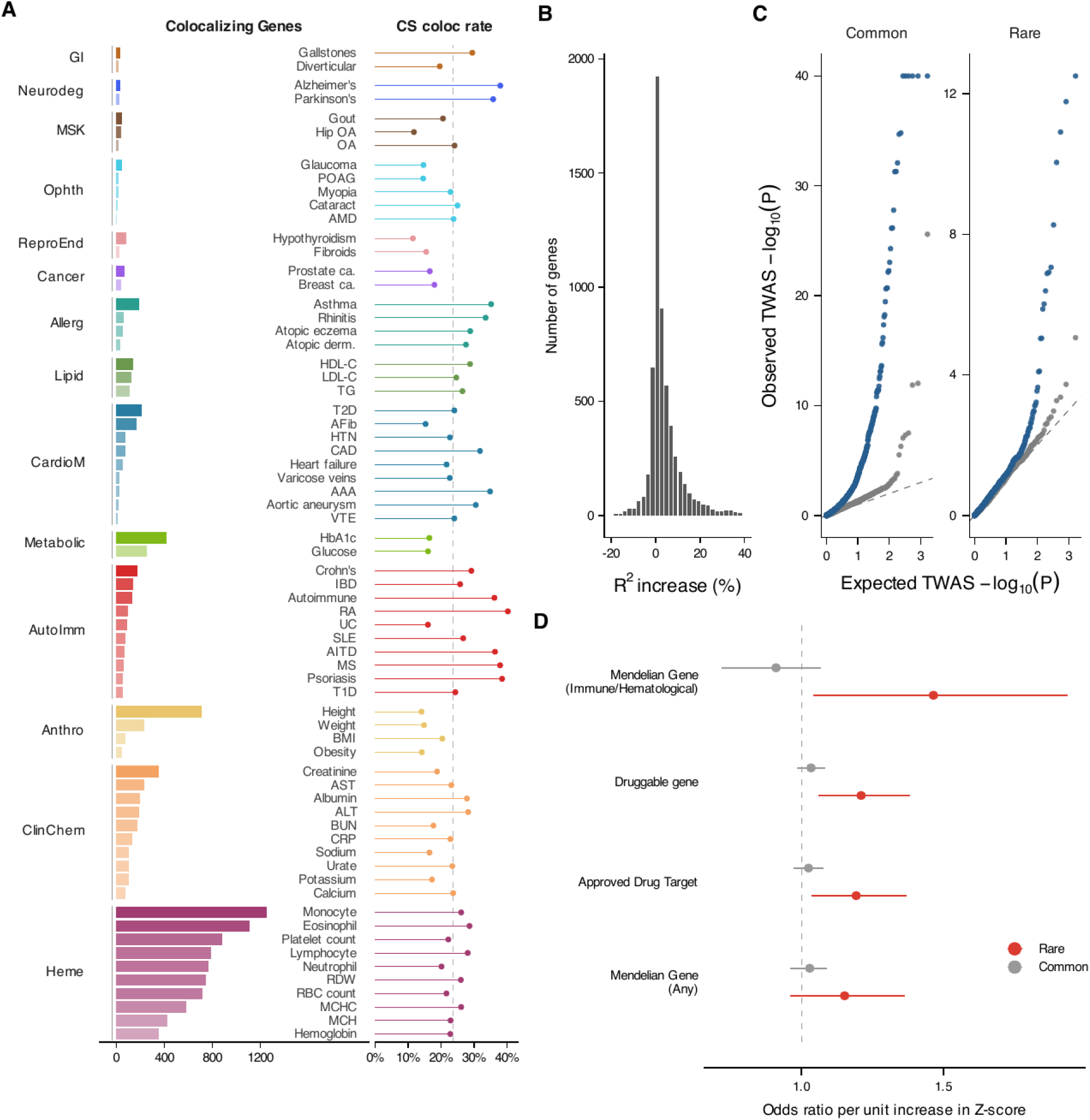
Rare-variant contribution to trait association at colocalized genes. **A.** Colocalization of molecular QTLs with GWAS by trait category. Left, the number of distinct genes with at least one QTL-GWAS colocalization for that trait colored by trait category. Right: the percentage of that traits GWAS credible sets colocalizing with any molecular QTL, dashed lines represent the median across displayed traits. Traits with five or fewer colocalizing genes were omitted. **B.** Distribution of R2 change in expression prediction from including rare variants across genes with a causal fine-mapped variant. **C.** Quantile-Quantile plots of conditional-TWAS association P values across nine blood traits shown separately for common-variant and rare conditional tests. Blue points show genes with colocalizations and grey show a matched null set based on expression level, constraint, rare-variant count. **D.** Enrichment of rare conditional TWAS signals across four clinically relevant gene sets adjusted for constraint and expression level. Points are the odds ratio per one unit increase in |Z| for rare (red) and common (grey) with 95% confidence intervals.

Importantly, colocalizations often converged across molecular layers. We identified 1,617 gene–trait pairs with colocalization support from multiple QTL classes, including 100 supported by all three molecular QTL types (Supplemental Figure 24,Supplemental Table 16). CD40 provided an illustrative example: distinct eQTL and pQTL credible sets supported colocalization of CD40 across seven GWAS involving inflammatory bowel disease, multiple sclerosis, rheumatoid arthritis, systemic lupus erythematosus, LDL cholesterol, and monocyte count (Supplemental Figure S25).

To complement our colocalization analyses, we next used AoU phenotype data (*17*) in a transcriptome-wide association study (TWAS) framework (*18*) (Methods). The major advantage of TWAS is the ability to integrate both common and rare QTLs. We first asked whether incorporating rare eQTL variants improves the prediction of genetically regulated expression. Across 7,226 genes harboring both common and rare eQTLs, models including rare variation exhibited improved cross-validated prediction accuracy relative to models restricted to common variants (mean ΔR² = 4%; Fig 4B). Further, prediction accuracy increased with increasing expression extremity, indicating that rare variation increases prediction of extreme molecular phenotypes.

We next asked whether the rare-variant component of molecular prediction improves prioritization of trait-associated genes beyond common-variant TWAS. Across 22 traits, we partitioned fine-mapped molecular predictors into common- and rare-variant components and tested the rare component conditional on the common component (Methods, Supplemental Table 17). Conditional rare-variant association signals were significantly enriched among colocalized gene-trait pairs relative to matched non-colocalized pairs (OR = 3.29, p-value = 4.1e-9) and significantly enriched for druggable genes (OR = 1.21, p-value = 0.004), approved drug targets (OR = 1.19, p-value = 0.013) from the Drug-Gene Interaction Database (*19*), and Mendelian disease genes (*20*) (OR = 1.47, p-value = 0.013). Together, these results showed that rare QTLs supplement common-variant TWAS to prioritize biologically and clinically-relevant effector genes.

### Molecular consequences of rare variation in known disease genes

Rare genetic variation is abundant in clinically relevant genes, yet the molecular consequences of most variants remain unresolved (*21*). Given the multiple genetic ancestry design of the AoU multi-omics cohort, we evaluated if there were rare QTLs that impact known disease genes.

Across 13,547 credible sets (46% resolved to single-variant credible sets), we identified 5,186 rare molecular QTL variants spanning 944 disease-associated genes annotated in ClinGen (*22*) (Fig. 5A top).

**Figure 5.**
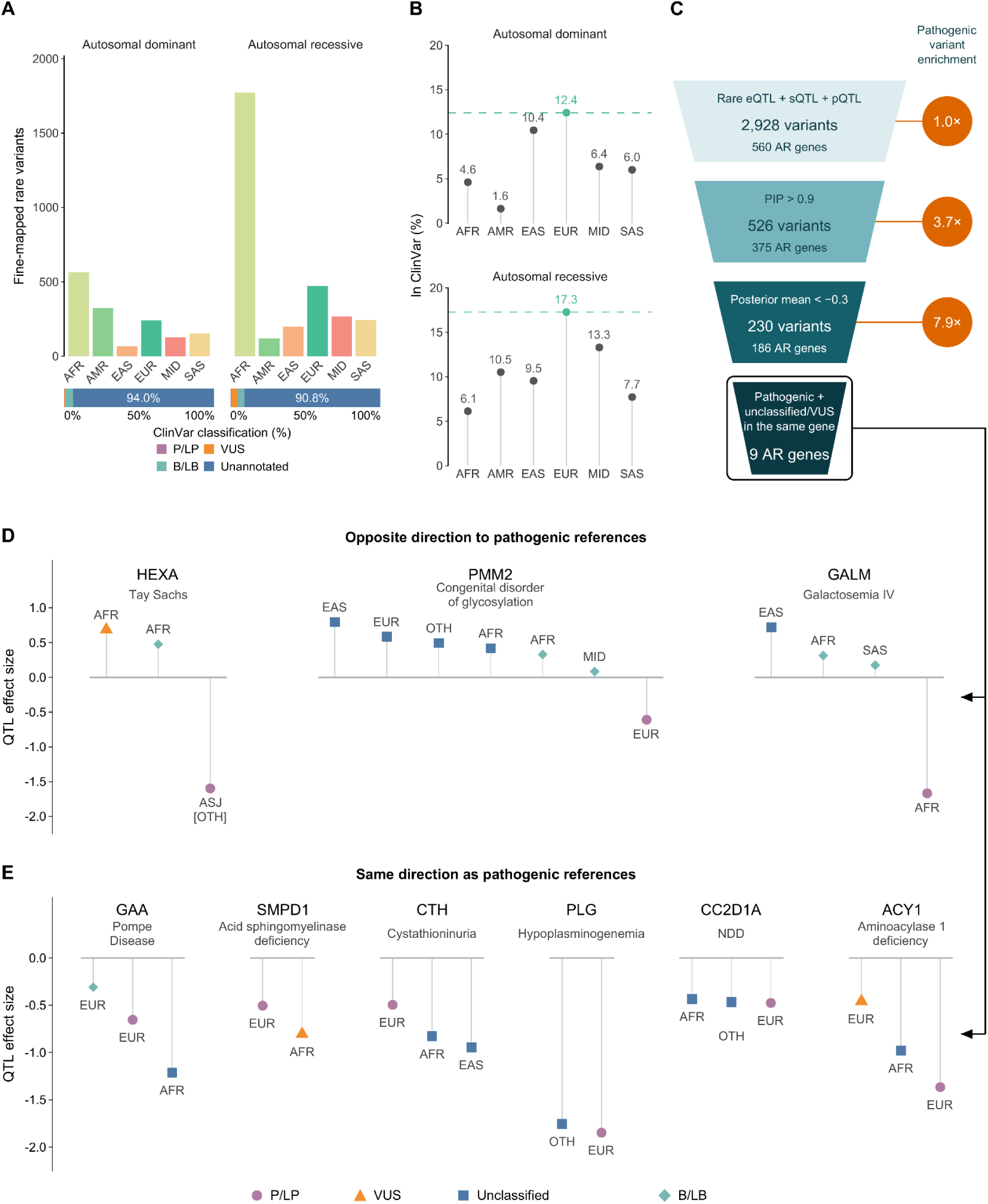
Rare molecular QTLs at ClinGen disease-associated genes. **A.** Numbers of rare fine-mapped expression (eQTL), splicing (sQTL), and protein (pQTL) variants at autosomal dominant and autosomal recessive disease genes with strong or definitive ClinGen associations. Variants have a maximum allele frequency <1% and are counted once across QTL types within each inheritance group. Bars group variants by the population with the highest allele frequency. Horizontal stacked bars show ClinVar classification proportions within each inheritance group. **B.** Percentage of variants with any ClinVar classification within each population and inheritance group. Variants are assigned to EUR when their EUR allele frequency is ≥75% of the maximum across the six displayed populations; otherwise, the highest-frequency population is used. Green points and dashed lines indicate EUR annotation coverage. **C.** Sequential prioritization of rare variants at autosomal recessive genes. Starting with 2,928 variants across 560 genes carrying P/LP, VUS, or no ClinVar annotation, filtering for PIP >0.9 retains 526 variants across 375 genes, with 3.7-fold enrichment of P/LP annotations (*P* = 1.05 × 10⁻⁶). Further filtering for posterior mean QTL effect <−0.3 retains 230 variants across 186 genes, increasing enrichment to 7.9-fold (*P* = 3.62 × 10⁻¹⁰). Enrichments are relative to the starting pool; unadjusted *P* values are from two-sided Fisher’s exact tests comparing each retained subset with the variants excluded from that pool. The final step identifies nine genes containing both a P/LP-reference variant and a distinct unclassified or VUS variant among rare high-PIP variants in either effect direction. **D., E.** Posterior mean QTL effects for reference and candidate variants at the nine selected genes, with selected benign comparators. Candidate effects are opposite in direction to the references in **D** and concordant in **E**. Colors and shapes indicate ClinVar classes; labels indicate ancestry annotations.

Strikingly 94.9% of these variants were either unclassified (92.1%) or annotated as variants of uncertain significance (2.8%) in ClinVar (*23*), with only 1.1% (42 variants) having an existing pathogenic classification (Fig 5A bottom). ClinVar annotations were also significantly less frequent in non-European genetic similarity groups for variants in autosomal dominant and recessive genes (OR = 0.33 and 0.38; P = 4.4 × 10⁻⁶ and 6.5 × 10⁻¹² respectively; two-sided Fisher’s exact tests; Fig 5B), demonstrating the lack of representation of group-enriched variation in current clinical variant databases. Despite the lack of annotation, known pathogenic variants had significantly greater molecular effects than benign variants (p-value = 0.00576) (Supplemental Figure S26). We also observed that the magnitude of the molecular effect varied systematically according to disease inheritance mode with the largest effects seen in the autosomal recessive genes (Supplemental Figure S27). We then asked whether rare causal eQTLs could produce loss-of-function-like effects. Of the 574 high confidence rare causal eQTLs (PIP > 0.9) affecting ClinGen disease genes, 18.3% had log₂(aFC) < −0.5 (7.8% with log₂(aFC) < −1), identifying candidate loss-of-function alleles (Supplemental Figure S28).

We next examined if specific high-effect variants could recover known pathogenic alleles and provide functional evidence for uncharacterized or VUS variants. As a proof of principle, we recovered a well-established pathogenic recessive Tay–Sachs indel which resulted in a strong reduction in *HEXA* expression(log2 aFC = −0.85) (Fig 5D, Supplemental Figure 29). Starting with 2,928 rare variants at autosomal recessive ClinGen genes with either no ClinVar annotation, a VUS annotation or a pathogenic/likely pathogenic assertion we applied a series of filters to prioritize the unclassified and VUS variants that were most likely to be pathogenic. First, restricting to likely causal molecular QTL variants (PIP > 0.9) yielded 526 variants across 375 genes and increased the proportion of variants with a pathogenic/likely pathogenic assertion by 3.7 fold compared to the original set. Further restricting to variants with a posterior mean QTL effect < −0.3 resulted in 230 variants across 186 genes and increased the enrichment to 7.9 fold compared to the original set (Fig 5C). Finally, within the 186 genes that remained we examined all of the rare high-PIP variants (PIP > 0.9) with either effect direction. This identified nine genes which contained both a variant with a pathogenic/likely pathogenic ClinVar assertion and a distinct unclassified or VUS variant (Fig 5C).

Comparison to the pathogenic references revealed different molecular effects for these nine genes. At *HEXA*, *GALM,* and PMM2 the unclassified/VUS variants were associated with increased expression or protein abundance (in contrast to the pathogenic references) and were more in line with the benign/likely benign counterparts(Fig. 5D). In contrast the unclassified or VUS variants at *GAA*, *SMPD1*, *CTH*, *PLG*, *CC2D1A* and *ACY1* all had negative effects matching the direction of the pathogenic references (Fig 5E). Several of them had effect magnitudes comparable to or greater than the pathogenic references, including an unclassified *GAA* eQTL and a *SMPD1* VUS pQTL.

For the genes where there was no known pathogenic variant amongst the fine-mapped signals we further refined the prioritisation using trait effects from AoU and predicted regulatory consequences. Examples included a VUS in *F10* with an INR (blood clotting time) effect in the top percentile of all variant effects and an unclassified variant in *ANGPTL3* which had a triglyceride effect in the bottom percentile (Supplemental Figure S30). These were in line with the bleeding phenotype of factor X deficiency and the low lipid levels seen in ANGPTL3 loss of function. At *ACVRL1*, a gene associated with hereditary haemorrhagic telangiectasia, PromoterAI predicted strongly reduced activity of a fine-mapped promoter variant (Supplemental Figure S31). *SPR* deficiency is known to impair brain dopamine and serotonin synthesis and result in dystonia so the neural regulatory predictions are particularly pertinent. Borzoi predicted 96% reduction in CTCF signal and 76% reduction in chromatin accessibility at the *SPR* deletion in the neural contexts (Supplemental Figure S32).

## Discussion

The AoU multi-omics cohort provides the scientific community with new avenues to study molecular mechanisms involved in human health and disease. Prior efforts in the UK Biobank have demonstrated the key value of multi-omics data when coupled to diverse phenotypic information from identifying the molecular effects of genetic variants (*5*) to discovering novel biomarkers (*24–26*). Further, such data provides multiple avenues to derisk drug targets by aiding in gene prioritization and establishing effect directionality (*27*). Here, the AoU multi-omics cohort expands on these opportunities by focusing on an ancestrally-diverse cohort in which both transcriptome and proteome data were generated to enable deeper insights into the effects of genetic variants across multiple major US ancestries.

The design of the AoU multi-omics cohort enables multiple facets of study from the expanded quantifications using Total RNA-sequencing, increased repertoire of 5,415 proteins to evaluating ancestry-enriched effects and cross-omic effects. We demonstrate several key insights from these data. First, we provide a comprehensive set of e/p/sQTL data expanding the discovery of independent QTLs for each gene. We also leverage the multi-ancestry design, which provides increased resolution of fine-mapped alleles (*28*, *29*), to provide credible sets for each QTL.

The expansive set of independent QTLs in the AoU multi-omics cohort enabled a deeper survey of the genetic architecture of molecular effects. By combining QTLs, we were able to identify dosage effects carried by each individual suggesting that locus-level allelic heterogeneity can achieve effects on par with gene deletions or duplications. The cohort of ∼10,000 individuals also allowed us to report on rare QTLs down to MAFs of 0.05%. The map of these rare effects provides a resource of larger effect variants influencing known disease genes and drug targets. Further, we demonstrate that these effects are more likely to replicate across transcriptome and proteome and increase power for gene prioritization approaches through TWAS analyses.

By focusing on rare eQTLs, we also provide a compendium of variants that may have imminent clinical utility. We observe 5,186 rare eQTLs impacting 922 disease-associated genes in ClinGen. While AoU is not a rare disease cohort, many healthy individuals may be carriers of Mendelian disease alleles. We demonstrate the ability of biobank-scale multi-omics to identify large-effect rare variants within rare disease genes that may support classification of pathogenicity for variants of uncertain significance or identify new potentially pathogenic alleles enriched in specific ancestry groups but absent from ClinVar.

The ancestry enrichment patterns reported here should be interpreted in the context of the genetic ancestry represented in the All of Us cohort. These data do not capture the full global or local spectrum of genetic ancestry, and a variant enriched in one ancestry group in this study may also occur in populations not represented or adequately sampled here. Thus, “ancestry enriched” describes relative frequency in the studied cohort and does not imply that a variant is restricted to, or exclusive to, that group.

Collectively, the AoU multi-omics cohort enables ongoing mechanistic investigations of health and disease. We demonstrate the significant value of generating multi-omics within an ancestrally diverse cohort. We expect that future investigators will benefit from use of these data in several ways from specific disease studies, exploration of omics relationships with diverse AoU demographic variables, through to accelerating biomarker discovery and therapeutic development.

## Data Availability

The All of Us Research Hub has a tiered data access data passport model with three data access tiers. The Public Tier dataset contains only aggregate data with identifiers removed. These data are available to the public through Data Snapshots (https://www.researchallofus.org/data/tools/data-snapshots/) and the public Data Browser (https://databrowser.researchallofus.org/). The Registered Tier curated dataset contains individual-level data, available only to approved researchers on the Researcher Workbench. At present, the Registered Tier includes data from EHRs, wearables and surveys, as well as physical measurements taken at the time of participant enrolment. The Controlled Tier dataset contains all data in the Registered Tier and additionally genomic data in the form of short- and long-read whole-genome sequencing,RNA-sequencing, proteomics, and genotyping arrays, previously suppressed demographic data fields from EHRs and surveys, and unshifted dates of events. At present, Registered Tier and Controlled Tier data are available to researchers at academic institutions, nonprofit institutions, industry affiliates,and both nonprofit and for-profit health care institutions. Researchers have the option to register for Registered Tier and/or Controlled Tier access by completing the All of Us Researcher Workbench access process and the *All of Us* Responsible Conduct of Research training, which includes identity verification and All of Us-specific training in research (https://www.researchallofus.org/register/). Researchers may create a new workspace at any time to conduct any research study, provided that they comply with all Data Use Policies and self-declare their research purpose. Data used in these analyses were released in the Curated Data Repository Version 9. Information on quality control of the data is available at on the Researcher workbench: https://support.researchallofus.org/hc/en-us/articles/50655639562900-All-of-Us-Genomics-Multi-omics-Quality-Report. Descriptions of the data-omics data available and the formats are provided: https://support.researchallofus.org/hc/en-us/articles/49999549117588-How-the-All-of-Us-Genomic-and-mulit-omics-data-are-organized.

This information is made accessible publicly on the All of Us Research Projects Directory at: https://allofus.nih.gov/protecting-data-and-privacy/research-projects-all-us-data.

## Acknowledgements

We gratefully acknowledge All of Us participants for their contributions, without whom this research would not have been possible. We also thank the National Institutes of Health’s All of Us Research Program for making available the participant data examined in this study. We thank Tara Dutka as well as current and former steering committee members Toral Contractor, Eric Dishman, Alyssa Cotler, Kelly Gebo, Gwynne Jenkins, Chris Lunt, Michelle McNeely, Dara Richardson-Heron, Karriem Watson, Michael Castro, Joni Rutter, Geoffrey S. Ginsburg, Rachele Peterson, Justin Hentges, and Sheri D. Schully for their service and contributions to the All of Us Research Program.

## Code Availability

All code for processing pipelines and analysis is available at https://github.com/AoU-Multiomics-Analysis

The project has been made public by All of Us as a featured workspace available here: https://workbench.verily.com/workspaces/multiomics-qtls-demo-project

## Author information

### All of Us Research Program - Consortium Contributors

**Steering Committee:** Habibul Ahsan^11^, Hoda Anton-Culver^12^, Deborah Banks^2^, Melissa A. Basford^10^, Randee Bloom^2^, Hugo Campos^2^, Mine S. Cicek^14^, Elizabeth Cohn^16^, Stephanie H Cook^17^, Linda Cottler^18^, Kathy Deerinwater^2^, Anjali Forber-Pratt^19^, Stacey Gabriel^9^, David Glazer^20^, Melissa Haendel^22^, Paul Harris^10^, Scott Joseph Hebbring^23^, Beverly Holmes^24^, Gail P Jarvik^8^, Jason Karnes^25^, Winston Koo^2^, Bruce Korf^26^, Monica Kraft^27^, Kate LeFauve^38^, KiTani Lemieux^29^, Mitchell R. Lunn^30^, April Brinkoetter McCrea^32^, Debra Dianne Murray^6^, Saer Oan^2^, Lucila Ohno-Machado^33^, Akinlolu Ojo^34^, Cathryn Peltz-Rauchman^35^, Steven Reis^5^, Edgar M Gil Rico^36^, Grisel Marie Robles-Schrader^37^, Linda Salgin^59^, Suaih Morales^2^, Edward Schlicksup^68^, Stephen Smith^38^, Jordan Smoller^39^, Andreas Theodorou^25^, Cheryl Thomas^40^, Eric Topol^41^, Phil Tsao^42^, Jen Uhrig^69^, Julia Moore Vogel^41^, Eboni Winford^43^, Katrina Yamazaki^44^, Stephan Zuchner^45^

**The Broad-LMM-Color Genome Center:** <u>Stacey Gabriel</u>^9^, <u>Niall Lennon</u>^9^, <u>Heidi L. Rehm</u>^9^, Scott Topper^1^, Namrata Gupta^9^, Konrad J. Karczewski^9^, Lee Lichtenstein^9^, Wenhan Lu^9^, Jeremy Guez^9^, Riley Grant^9^, Matt Solomonson^9^, Ben Neale^9^

**Baylor Hopkins Center for Clinical Genomics:** <u>Richard A. Gibbs</u>^6^, <u>Kimberly F. Doheny</u>^4^, <u>Eric</u> <u>Boerwinkle</u>^3^, Ginger A. Metcalf^6^, Donna M. Muzny^6^, Eric Venner^6^

**Northwest Genomics Center:** <u>Gail P. Jarvik</u>^8^, <u>Evan E. Eichler</u>^8^, <u>Chia-Lin Wei</u>^8^, Phillip E. Empey^5^, Christian Frazar^8^, Tina Lockwood^8^, Brian Shirts^8^, Joshua D. Smith^8^, Colleen Davis^8^

**The Biobank:** <u>Mine S. Cicek^14^</u>, Tony G. Bilyeu^14^, Ashley L. Blegen^14^, Jolene J. Lane^14^, Jeffrey G. Meyer^14^

**The Data and Research Center:** <u>Paul A. Harris</u>^10^, <u>Melissa A. Basford</u>^10^, <u>Dan M. Roden</u>^10^, Emmanuel Ankumah-Saikoom^10^, Robert J. Carroll^10^, Justin A. Cook^10^, Megan K. He^10^, Aymone Jeanne S. Kouame^10^, Lee Lichtenstein^9^, Jodell E. Linder^10^, Michael Lyons^10^, Brandy M. Mapes^10^, Kayla Marginean^10^, Megan Morris^10^, Jun Qian^10^, Ryan Samarakoon^10^, Samantha Stewart^10^, Chun-I Tang^10^, Yuanyuan Wang^10^, Consuelo H. Wilkins^10^

**The Participant Center (including Scripps Research Translational Institute, CareEvolution and Sage Bionetworks):** Sasri Dedigama^41^, Megan Doerr^66^, Vik Kheterpal^67^, Christine Suver^66^, Ashley Tate^67^, Julia Moore Vogel^41^, John Wilbanks^66^

**The Participant Technology Systems Center:** <u>Mark Begale</u>^48^, <u>Scott Sutherland</u>^48^, Jeff Drommerhausen^48^, John Durham^48^, Catherine E. W. Freeland^48^, Cecilia Gacek^48^, Gail Hamilton^48^, Aditya Naik^48^, Julie Shaw^48^, Daniel Uribe^48^

**Acknowledging former members of the Steering Committee:** Ivye L. Allen^2^, Katie Baca-Motes^41^, Laura Bartlett^46^, Debra Bartley-Rice^47^, Mark Begale^48^, Eric Boerwinkle^3^, Murray H Brilliant^23^, Jessica Burke^13^, Patricia Butts^2^, John Chaffins^32^, Katherine Chang^2^, Veena Channamsetty^49^, Carmen Chinea^50^, Cheryl Clark^15^, Vivian Colon-Lopez^51^, Karl Cooper^19^, Carolyn Cronin^2^, Martha Daviglus^52^, Liliana Lombardi Desa^2^, Charles E. Drum^19^, Mark Edmunds^53^, Dejen M. Eshete^2^, Adolph Falcón^36^, Miguel Flores^2^, Gretchen Funk^54^, Ali G. Gharavi^55^, David B. Goldstein^55^, Joannie Grand^13^, Philip Greenland^21^, Kristin Hall^56^, Kimiyo Harris-Williams^2^, Edina Harsay^2^, Richard Hochfelder^2^, Nazia Hussain^2^, Praduman Jain^48^, Ayanna Jenkins^47^, Christine C. Johnson^35^, Anna Johnston^2^, Parinda Khatri^43^, Linda Perez Laras^28^, Patricia Watkins Lattimore^40^, Brendan Lee^6^, Dessie Levy^2^, Megan Lewis^2^, Emily Makahi^31^, Patrick McGovern^32^, Martin Mendoza^57^, Wanda Montalvo^58^, Francisco Moreno^25^, Fatima Munoz^59^, Narayana Murali^60^, Christopher O’Donnell^42^, May Okihiro^61^, Greg Orr^56^, Andrea Ramirez^10^, Fornessa Randal^62^, Dan Roden^10^, Eric Schlueter^24^, Yashoda Sharma^27^, Hannah Sinemus^5^, Vicki Smith^41^, Teshia Solomon^25^, Louisa Stark^63^, Steven Steinhubl^41^, Christine Suver^64^, Gregory Talavera^59^, Amy Taylor^49^, Ronnie Tepp^65^, Steve Thibodeau^14^, Scott Topper^1^, Vanessa Uzoh^2^, David Wellis^53^, Consuelo Wilkins^10^, Amanda Wilson^46^, Joyce Winkler^2^, Alicia Y. Zhou^1^

**Acknowledging in memoriam:** Stephen Mikita^2^, Deborah A Nickerson^8^, Lauren Ryan^1^, Karl Surkan^2^

**Affiliations:** ^1^Color Health; ^2^Participant Steering Committee Member; ^3^The University of Texas Health Science Center at Houston; ^4^Johns Hopkins University School of Medicine; ^5^University of Pittsburgh; ^6^Baylor College of Medicine; ^7^Laboratory for Molecular Medicine; ^8^University of Washington; ^9^Broad Institute; ^10^Vanderbilt University Medical Center; ^11^University of Chicago; ^12^University of California, Irvine; ^13^The MITRE Corporation; ^14^Mayo Clinic; ^15^Brigham and Women’s Hospital; ^16^The Feinstein Institutes for Medical Research; ^17^New York University; ^18^University of Florida; ^19^American Association on Health and Disability; ^20^Verily Life Sciences; ^21^Northwestern University; ^22^University of North Carolina School of Medicine; ^23^Marshfield Clinic Research Institute; ^24^Cooperative Health; ^25^University of Arizona; ^26^University of Alabama at Birmingham; ^27^Icahn School of Medicine at Mount Sinai; ^28^San Juan Bautista School of Medicine; ^29^Xavier University; ^30^Stanford University School of Medicine; ^31^Waianae Coast Comprehensive Health Center; ^32^HungryHeart Media, Inc. (Wondros); ^33^Yale School of Medicine; ^34^University of Kansas Medical Center; ^35^Henry Ford Health; ^36^National Alliance for Hispanic Health; ^37^Pyxis Partners; ^38^NORC; ^39^Partners Healthcare System; ^40^Delta Research and Educational Foundation; ^41^Scripps Research Translational Institute; ^42^U.S. Department of Veterans Affairs; ^43^Cherokee Health Systems; ^44^California State University Los Angeles; ^45^University of Miami School of Medicine; ^46^National Library of Medicine; ^47^Jackson-Hinds Comprehensive Health Center; ^48^Vibrent Health; ^49^Community Health Center; ^50^HRHCare; ^51^University of Puerto Rico Comprehensive Cancer Center; ^52^University of Illinois at Chicago; ^53^San Diego Blood Bank; ^54^FiftyForward; ^55^Columbia University Irving Medical Center; ^56^Walgreens; ^57^National Institutes of Health; ^58^University of Pennsylvania; ^59^San Ysidro Health; ^60^Geisinger Health System; ^61^University of Hawai’i; ^62^Asian Health Coalition; ^63^University of Utah School of Medicine; ^64^Emory University; ^65^HCM Strategists LLC; ^66^Sage Bionetworks; ^67^CareEvolution; ^68^IBM; ^69^RTI

**Note:** Underlining indicates the grant’s Principal Investigators

## Methods

Reported results with group counts under 20 are disclosed under a granted exception to the Data and Statistics Dissemination Policy from the All of Us Resource Access Board.

### Proteomics Processing

The proteomics processing workflow is designed to generate protein expression profiles from 10,056 blood plasma samples using the Olink Explore HT platform. These samples were systematically organized into 61 individual Olink batches, each consisting of two plates, and were processed across two primary rounds. To address potential batch effects and ensure longitudinal consistency, the workflow incorporated approximately 127 replicate samples, including 25 specific bridge samples designated for software-based bridging analysis.

### Reference median-based normalization

To remove technical artefacts we used a reference median based normalisation approach. Control samples were excluded prior to normalisation. A single plate was selected as the reference and assay specific median NPX values were calculated from that plate. The NPX values from all other plates were then adjusted to these reference medians using the Olink normalisation procedure, resulting in a harmonised set of values across plates with the relative biological variation being preserved. The choice of reference plate had minimal impact on downstream analysis.

### Outlier sample removal

Sample-level proteomic quality control was performed using a network connectivity approach applied to the reference median normalized Olink NPX matrix. Briefly, pairwise biweight midcorrelations were calculated across all samples, and the resulting adjacency matrix was used to estimate sample connectivity(*30*). Connectivity values were converted to z scores, and samples with connectivity z scores less than −3 were designated as outliers and removed prior to downstream analysis.

### RNA-sequencing Processing

#### Read alignment and expression quantification

Reads were extracted from input CRAM files using Picard SamToFastq v2.27.1 and aligned to the hg38 reference genome using STAR v2.7.11b (*31*). The reference genome used did not include the ALT, HLA or decoy contigs and gene annotations were based on GENCODE v48 (*32*). WASP (*33*) filtering was enabled during alignment to reduce reference mapping bias. Duplicate reads were marked using Picard MarkDuplicates v2.27.1. Gene level raw counts and RNA-seq QC metrics were generated using RNA-SeQC 2 v2.4.3 (*34*) with a collapsed GENCODE v48 GTF annotation.

#### Expression normalization

Raw count data were transposed into a sample by gene matrix and filtered to keep only those genes which had a count >6 in at least 20% of samples. The filtered counts were then normalised using the TMM method (in edgeR (*35*)) and converted to counts per million (CPM).

To stabilize variance and approximate normality, expression values for each gene were rank-based inverse normal transformed across samples. The normalized expression matrix was then merged with gene-level TSS coordinates to generate a BED-formatted file containing genomic positions and normalized expression values for each gene across samples.

#### Splicing Quantification

Per sample splice junction files generated with regtools were jointly clustered across samples using LeafCutter v0.2.9 (*36*). Junctions were clustered into intron clusters using the leafcutter_cluster_regtools.py workflow with a minimum of 30 reads per cluster, a minimum junction to cluster read ratio of 0.001 and a maximum intron length of 500kb. This generated the per-intron count tables across all samples for downstream splicing quantification.

Intron clusters were then assigned to genes by intersecting the intron boundaries with the exon annotations. Under a strict assignment scheme a cluster was only assigned to a gene if both splice sites fell at annotated exon boundaries. Introns with insufficient representation or low information content were removed prior to the QTL analysis (those with zero usage in more than 50% of samples, low variability across individuals and those on the Y chromosome).

### QTL identification and covariate calculation

#### Genotype Principal Component Calculation

Joint srWGS variants were filtered to biallelic variants, an AC threshold of 10 and a call rate of 95% was applied, and the VCFs were converted to PLINK(*37*) for downstream QTL analysis. Genetic PCs were calculated using SNPRelate after pruning with SNPs with an LD threshold = 0.2 and MAF = 0.01. The number of components to retain was determined using the Gavish–Donoho(*38*) method for optimal hard thresholding, which estimates the effective dimensionality of the data based on the singular value spectrum.

#### Phenotype Principal Component Calculation

Phenotype PCs for the covariates were calculated via PCA on the sample by molecular phenotype matrix using PCAtools v2.22.4. The number of components to retain was determined using the Gavish-Donoho method for optimal hard thresholding (this method estimates the effective dimensionality of the data from the singular value spectrum). The requisite number of PCs were then extracted from the rotated PCA matrix (each PC representing a major axis of variation in expression across the samples).

#### cis-QTL Identification

cis-QTL mapping was performed using TensorQTL(*39*). The mapping window was defined as 1 Mb up and down-stream of the transcription start site for each feature. All variants with a MAC >= 10 were used as input within each cohort. Phenotype and genotype principal components were used as covariates. A gene was considered to have a significant QTL if the corresponding FDR value was less than 0.05. QTL identification was performed within each ancestry and in a pooled fashion using all samples.

### Ancestry Group Definitions

Ancestry group assignment was defined using measures of genetic similarity of each participant to global reference populations using the Human Genome Diversity Project(*13*, *40*, *41*), 1000 Genome Project, and gnomAD (https://support.researchallofus.org/hc/en-us/article_attachments/52524243515540 Appendix G). The following acronyms are used to describe genetic similarity to a reference population AFR (1KGP-HGDP-AFR-like); AMR (1KGP-HGDP-AMR-like); EAS (1KGP-HGDP-EAS-like); EUR (1KGP-HGDP-EUR-like); MID (1KGP-HGDP-MID-like); SAS (1KGP-HGDP-SAS-like); and REM, not belonging to one of the other ancestries or is an admixture. Due to uneven sampling across global populations within the HGDP and 1000 Genomes reference panels, inferences regarding genetic background and admixed ancestry may exhibit reduced precision for participants originating from underrepresented groups, warranting careful interpretation of these estimates.

### QTL cross-ancestry meta analysis and replication

#### Cross-ancestry meta-analysis

To assess the consistency of molecular QTL signals across ancestry groups, we performed cross-ancestry meta-analysis using METASOFT (*42*), excluding the pooled and admixed cohorts. For each molecular trait and ancestry group, we extracted the effect size estimate and standard error for the lead QTL variant and used these as input to METASOFT to assess the consistency of genetic effects across ancestries and estimate the posterior probability that an effect was present within each population. METASOFT estimates ancestry-specific M-values, which represent the posterior probability that a genetic effect exists within a given ancestry group. Associations were considered present when the corresponding M-value exceeded 0.9. We applied this analysis to eQTLs, sQTLs, and pQTLs and summarized both the number of ancestry groups supporting each association (M-value > 0.9) and the distribution of ancestry-specific M-values across populations.

#### Fine-mapping

To further pinpoint causal genetic variants, we applied Bayesian fine-mapping with SusieR (*12*) to significant cis-QTLs (FDR < 0.05). For each QTL, we extracted variants within a cis-window of +/-1 Mb of the transcription start site for that gene. Inverse-normal-transformed phenotypes were used for fine-mapping. Both the phenotype and genotype matrices were residualized with respect to the covariate matrix. Variants with no alternate alleles were removed, and missing genotype dosages were imputed to the variant mean. SuSiE was run with a maximum of 10 single-effect components (L=10), with residual and prior variances estimated from the data (estimate_residual_variance=TRUE, estimate_prior_variance=TRUE), a scaled prior variance of 0.1, and univariate z-scores computed (compute_univariate_zscore=TRUE). Credible set purity was assessed using a minimum absolute correlation threshold of 0.5 (min_abs_corr=0.5). In downstream reporting, we kept non-overlapping credible sets and excluded low-purity credible sets (minimum absolute correlation < 0.5).

#### Ancestry Enrichment Calculation

To quantify the level of ancestry enrichment amongst the fine-mapped variants we limited the analysis to variants with strong posterior evidence of causality (PIP > 0.9). For each variant we identified the ancestry group with the highest allele frequency and then compared the number of alleles and the number of allele counts in that group to the number of alleles and number of allele counts in all the other ancestry groups combined within the greater AoU genome sequencing cohort. An ancestry enrichment odds ratio was then calculated using Fisher’s exact test on the resulting contingency table, giving a measure of the degree to which the variant was enriched in its most frequent ancestry relative to all the other ancestry groups. Odds ratios > 5 were considered to indicate ancestry-specific enrichment.

#### Allelic Fold Change

Allelic fold changes (*14*) were estimated for all variants identified as causal (PIP > 0.9) through eQTL fine-mapping using log2(CPM) expression values rather than inverse normal-transformed expression values, as the latter do not preserve an interpretable expression scale. Principal components were recalculated from the log2(CPM) expression matrix using the same procedure employed for QTL mapping. Allelic fold changes were then estimated using the aFC software package https://github.com/secastel/aFC.

#### Cumulative eQTL Burden effects

To estimate the cumulative effect of multiple independent regulatory variants on expression we assumed additive effects in log2 expression space. For each gene the independently fine-mapped variants were split into expression increasing and expression decreasing sets based on the sign of their estimated allelic fold change. Within each direction the allelic fold changes of all causal variants were then summed in sequence, beginning with the variant of highest effect, to give an estimate of the cumulative log2 expression change. This gives an estimate of the expected heterozygous expression level under an additive model and across sequential number of variants.

### eQTL Burden Model

To quantify the regulatory burden carried by each individual, we first estimated the expected expression effect of fine-mapped eQTL variants on expression for each eGene. Because allele frequencies differ across ancestry groups, expected population-specific expression shifts calculated separately for each gene and ancestry using the observed allele frequencies and allelic fold change estimates under an additive model. Specifically, the expected expression shift was computed as ∑ᵢ 2pᵢβᵢ, where pᵢ is the ancestry-specific allele frequency and βᵢ is the estimated log₂ allelic fold change.

For each individual-gene pair, the predicted expression effect was centered by subtracting the ancestry-specific expected population shift. Centered effects were converted to percent expression change relative to the population expectation as (2^Δ − 1) × 100, where Δ denotes the centered log2 expression effect. Genes were subsequently classified according to their predicted expression perturbation, including deletion-like and duplication-like categories, and the median number of genes per individual across a range of predicted expression values.

### eQTL Burden enrichment

To assess enrichment of dosage-like QTL effects, we fitted separate generalized linear models for deletion-like and duplication-like QTL counts. For each annotation, we modeled the winsorized number of deletion-like or duplication-like QTL effects using a quasi-Poisson regression with a log link. Annotation status or score was included as the primary predictor, with, log(1 + Mean TPM), the number of credible sets, and the presence of any rare variants included as covariates

The annotation coefficient was exponentiated to obtain an incidence rate ratio (IRR). For IRRs below one, fold depletion was calculated as 1/IRR;for IRRs above one, fold enrichment was calculated as the IRR. Associations with *p* < 0.05 were reported as fold changes, whereas nonsignificant associations were not labeled.

### Cross-ome colocalization

Using independently fine-mapped signals from each molecular assay, we performed pairwise colocalization analysis with fastENLOC(*15*) to identify genes and variants with evidence of shared causal effects across transcriptional and post-transcriptional processes. Unlike other Bayesian colocalization methods, fastENLOC estimates colocalization priors directly from the fine-mapping results based on the total number of signals and variants analyzed. Because molecular features were not perfectly shared across QTL assays, we used only variants tested in cis-windows of the intersecting set of features for each pairwise comparison.

fastENLOC reports evidence for colocalisation at multiple levels: gene level colocalisation probability (GLCP), region level (credible set) colocalisation probability (RCP) and signal level (variant) colocalisation probability (SCP). For each pairwise comparison and each of the colocalisation metrics we have applied Bayesian false discovery rate correction and only events passing the appropriate significance threshold were retained.

### Trait colocalization

GWAS credible sets were analyzed against eQTL, sQTL, and pQTL datasets using fastENLOC(*15*) and a complementary CLPP-based approach(*43*).

FastENLOC was run independently for each GWAS analysis and QTL layer. Signal-level regional colocalization probabilities (RCP) and locus colocalization probabilities (LCP), together with gene-level regional and locus colocalization probabilities (GRCP and GLCP), were retained for downstream analysis.

To prevent duplicate counting of loci represented in multiple GWAS studies of the same trait, credible sets were merged across studies using variant-membership similarity. Credible sets were assigned to the same consensus locus when the number of shared variants divided by the total number of unique variants across both credible sets was at least 0.90. Only credible sets from different studies of the same trait were merged.

CLPP was calculated for each GWAS credible-set–QTL-signal pair by multiplying the GWAS PIP and QTL PIP for each variant shared between the two signals, then summing these products across all shared variants.

FastENLOC and CLPP evidence was harmonized at the signal, credible-set, and gene levels. Bayesian false-discovery-rate thresholds were calculated separately for each GWAS–QTL analysis pair at an FDR level of 0.05. RCP thresholds were estimated over signal-level probabilities, whereas GRCP and GLCP thresholds were estimated over gene-level probabilities. Probabilities were considered significant when they exceeded the threshold corresponding to a cumulative Bayesian FDR of 0.05.

Stringent colocalization evidence was defined by the union of RCP ≥ 0.5, CLPP ≥ 0.05, RCP Bayesian-FDR significance, or GLCP Bayesian-FDR significance. Colocalization was evaluated separately for each QTL layer and across QTL layers. In cross-layer analyses, a consensus locus or gene was counted once if it met the relevant criterion in at least one QTL layer

### TWAS Model Benchmarking

To assess the contribution of rare variants to transcriptomic prediction we compared cross-validated TWAS prediction models trained on all cis-variants to models trained after the removal of rare variants. For each gene individuals were split into cross-validation folds (10 folds), molecular phenotypes were rank normalised separately within each training and test set to prevent information leakage across folds. Expression principal components were estimated from the training samples within each fold and projected onto the test samples (the held out samples). The fold specific expression PCs were then combined with the genetic PCs and used as covariates.

Within each training fold we fine-mapped the cis effects using SuSiE and estimated the posterior mean effect sizes for variants in the credible sets. We trained two models per gene: one using all the cis variants and one using only those variants with an allele frequency >1%. Genetic prediction in the held out samples was the dosage weighted sum of the SuSiE posterior mean effect sizes. Expression in the held out samples was residualised using the covariate effects estimated from the corresponding training set and performance was assessed by comparing predicted to observed residualised expression across folds.

### TWAS association analysis

TWAS scores were calculated by combining the GWAS summary statistics with the SuSiE fine-mapping results for each molecular trait. For each gene, the GWAS variants that overlapped the genomic interval defined by the fine-mapped variants were pulled from the tabix-indexed summary statistics. Variants were kept if the chromosome, position, reference allele and alternate allele all matched between the two datasets; duplicate variant records and any variants that had a mismatch in allele were removed. Variant level GWAS Z-scores were then calculated by taking the GWAS effect estimate and dividing it by its standard error.

The TWAS score was calculated as the sum across variants of each variant’s SuSiE posterior mean weight multiplied by its GWAS Z-score. Its variance was calculated by combining the variant weights with the LD correlation matrix, such that each pair of weights was multiplied by the LD correlation between the corresponding variants and these products were summed. The TWAS Z-score was then calculated by dividing the TWAS score by the square root of its variance. Two-sided association P-values were obtained from the squared TWAS Z-score using a chi-squared distribution with one degree of freedom. LD matrices were computed from standardized genotype dosages after mean imputation of missing values.

### PromoterAI variant-effect predictions

Predicted effects of promoter variants were taken from PromoterAI (*44*), a deep-learning model that scores non-coding variants near transcription start sites (TSS). Precomputed scores were obtained for all three possible single-nucleotide substitutions at every position within ±500 bp of each annotated TSS, scored independently for each transcript.

For the ACVRL1 locus, scores were restricted to the transcript ENST00000419526.6, giving predictions for all 3,003 possible substitutions across the 1,001 positions within ±500 bp of its TSS (chr12:51,912,370). Within each window, every possible substitution at every position was plotted against its position relative to the fine-mapped variant, and substitutions with an absolute predicted effect of at least 0.5 were highlighted. The fine-mapped promoter variant chr12:51,912,316 C>T received a PromoterAI score of −0.739, placing it within the most extreme 1% of predicted effects for this transcript (1st percentile −0.688), consistent with the reduction in ACVRL1 expression observed for the alternate allele.

### Borzoi variant-effect predictions

Allele-specific regulatory predictions were generated with Borzoi (*45*), using the pretrained borzoi-replicate-0 weights. For the SPR variant chr2:72,887,998 GCGCC>G (a 4-bp deletion), a 524,288-bp window of hg38 sequence centered on the variant was one-hot encoded and passed through the model, which predicts 7,611 human genomic tracks over 6,144 output bins of 32 bp (a 196,608-bp predicted window). The alternate sequence was constructed by replacing the reference allele at the variant offset and padding the 3′ end with reference sequence so that both alleles presented the model with an identically sized input; predicted signal was mapped back to reference coordinates, the 4-bp shift being smaller than the 32-bp output resolution.

Output tracks were grouped by assay using the Borzoi target descriptions: CTCF and cohesin (RAD21, SMC3) ChIP–seq; chromatin accessibility (DNase and ATAC); the activating marks EP300, H3K27ac, H3K4me1 and H3K4me3; CAGE and RNA; and the broad or repressive marks H3K36me3, H3K27me3 and H3K9me3. Within each group, tracks whose description matched a neural context (brain, cerebellum, cortex, neuron, hippocampus, astrocyte, glia or nervous system) were retained when at least three such tracks were available, and all tracks in the group were used otherwise; predicted signal was then averaged across the retained tracks. This yielded brain-specific groups for CTCF (n = 6 tracks), accessibility (n = 77), H3K27ac (n = 5), H3K4me1 (n = 11), H3K4me3 (n =15), CAGE (n = 38), RNA (n = 39), H3K36me3 (n = 10), H3K27me3 (n = 11) and H3K9me3 (n =12), and pan-tissue groups for cohesin (n = 22). Reference and alternate group-averaged predictions were exported as bigWig tracks at 32-bp resolution.

The effect of the variant on each mark was quantified as the percentage change in mean predicted signal between alleles, 100 × (alternate − reference) / reference, computed within ±96 bp of that mark’s reference-track peak, with the peak located within ±750 bp of the variant.

Base-resolution contribution scores were computed by input-gradient attribution. The prediction objective was the summed predicted CTCF ChIP–seq signal over the five 32-bp bins centered on the variant, and the gradient of this objective with respect to the one-hot encoded reference input was multiplied elementwise by that input to give the contribution of the observed base at each position. Contributions are reported over a ±28-bp window centered on the variant

