## Supplemental Figures for "Multi-omics in the All Of Us Research Program expands mapping genetic mechanisms in health and disease"


| | | |
|---|---|---|
| S1 | GTEx RNA-seq comparison | 2 |
| S2 | Olink Precision and QC | 3 |
| S3 | Olink subcellular location enrichment among detected proteins | 4 |
| S4 | QTL discovery by population and combined analysis | 5 |
| S5 | Cross ancestry QTL meta-analysis | 6 |
| S6 | eQTL replication with external consortia | 7 |
| S7 | sQTL replication with external consortia | 8 |
| S8 | pQTL detection rate by LOD | 9 |
| S9 | GTEx noncoding RNA comparison | 10 |
| S10 | GTEx hemoglobin expression comparison | 12 |
| S11 | Credible sets per molecular trait by population and combined analysis | 13 |
| S12 | Credible-set sizes and differences from AFR | 14 |
| S13 | Lead-variant PIP and causal resolution across analyses | 16 |
| S14 | eQTL credible-set discovery across sample sizes | 18 |
| S15 | Fine-mapping annotation enrichment | 19 |
| S16 | AoU-cohort wide allele frequency by ancestry enrichment | 20 |
| S17 | Strongest aFC by allele frequency per gene | 21 |
| S18 | Extreme QTL predictions are enriched for protein underliers | 22 |
| S19 | Validated dosage-like events by gene category | 23 |
| S20 | Cross-QTL colocalization is robust across thresholds | 24 |
| S21 | Coloc rate by molecular pair | 25 |
| S22 | Colocalization between specific credible sets is more restricted than sharing at the gene level | 26 |
| S23 | Cross-omic colocalization becomes more likely as allele frequency decreases | 27 |
| S24 | QTL-trait colocalization identifies genes with convergent evidence | 28 |
| S25 | CD40 has convergent evidence for colocalization across QTL types | 29 |
| S26 | eQTL effects of ClinVar variants | 30 |
| S27 | Disease inheritance pattern by effect size | 31 |
| S28 | Rare eQTL variant counts by aFC threshold | 32 |
| S29 | HEXA eQTL fine-mapping | 33 |
| S30 | Trait prioritization for F10, ANGPTL3, SERPINF2, and IFT80 | 34 |
| S31 | ACVRL1 fine-mapping and PromoterAI predictions | 35 |
| S32 | SPR fine-mapping and Borzoi regulatory predictions | 37 |

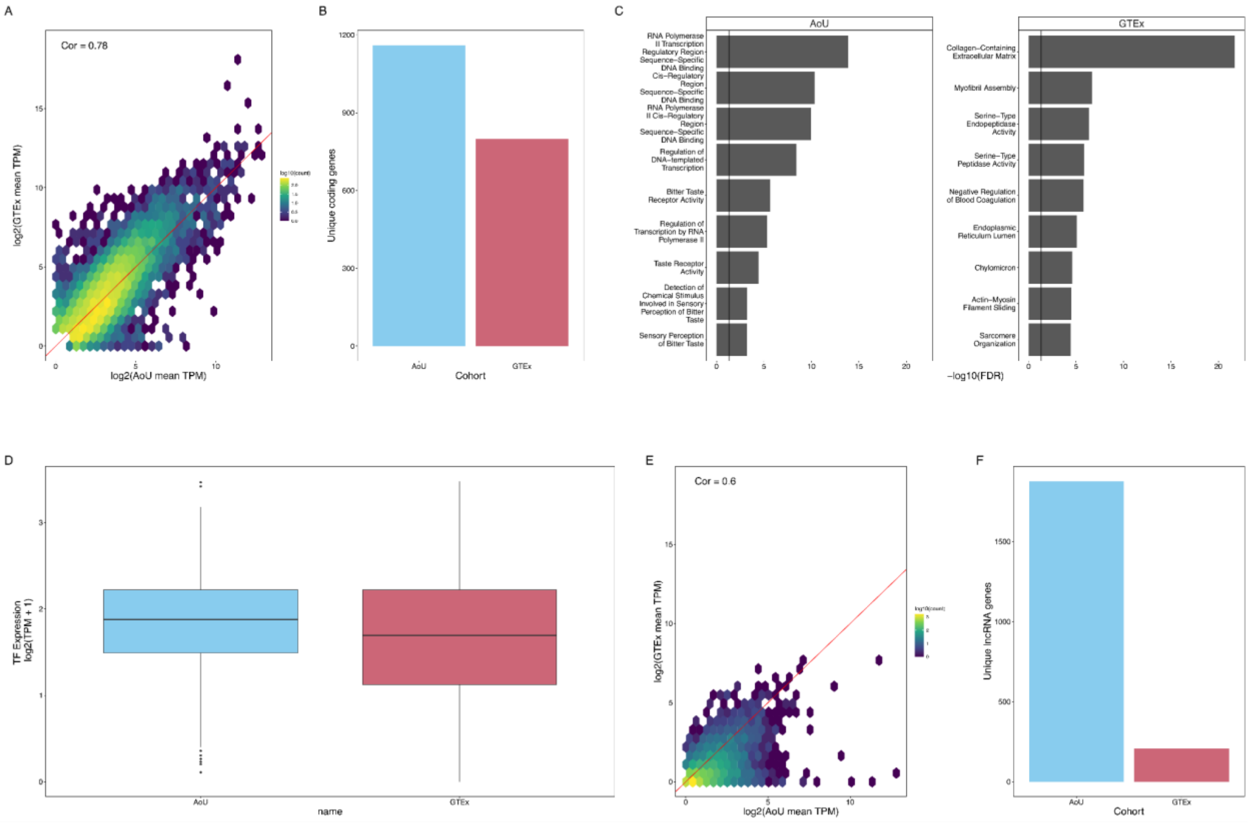


**Supplemental Figure S1. GTEx RNA-seq comparison**

5,000 individuals from the AoU RNA-sequencing data set were aligned to GENCODE v45 to benchmark against GTEx whole blood RNA seq. Mean expression per gene was computed across all samples in each cohort as transcripts per million (TPM); only genes quantified in both cohorts are shown. (A) Hexagonally binned density of protein-coding gene expression in All of Us (AoU; x axis, log2(mean TPM + 1)) against GTEx v10 whole blood (y axis, log2(mean TPM + 1)), restricted to protein-coding genes with a mean TPM greater than 1 in at least one cohort. Hexagon fill encodes the log10 of the number of genes falling in each bin; the red line is the identity line (y = x). Inset text gives the Pearson correlation coefficient between the two log-transformed means (r = 0.78). (B) Number of the protein-coding genes in A that were detected in exactly one cohort, defined as a mean TPM greater than 1 in one cohort and below 1 in the other; bar height is the count of unique genes specific to All of Us and to GTEx, respectively. (C) Gene Ontology (GO) enrichment of the two cohort-specific protein-coding gene sets defined in B, computed with Enrichr against the GO Molecular Function, GO Cellular Component and GO Biological Process 2025 term libraries. The nine most significant terms per cohort, ordered by Enrichr's adjusted P value, are shown. (D) Expression of transcription-factor (TF) genes in the two cohorts from genes in the Human Transcription Factors list (v1.01), and had a mean TPM greater than 1 in at least one cohort; the y axis is log2(mean TPM + 1) and each box shows the median and interquartile range with whiskers extending to the most extreme value within 1.5× the interquartile range (points beyond are plotted individually).

Expression was compared between cohorts with a two-sided paired t-test on the log2(mean TPM + 1) values and by ordinary least-squares regression of GTEx on All of Us log2(mean TPM + 1). (E) As in A, for long non-coding RNA (lncRNA) genes, restricted to lncRNAs with a mean TPM greater than 0.1 in at least one cohort (r = 0.60]). (F) As in B, for the lncRNA genes in E: the number of lncRNAs with a mean TPM greater than 1 in one cohort and below 1 in the other.

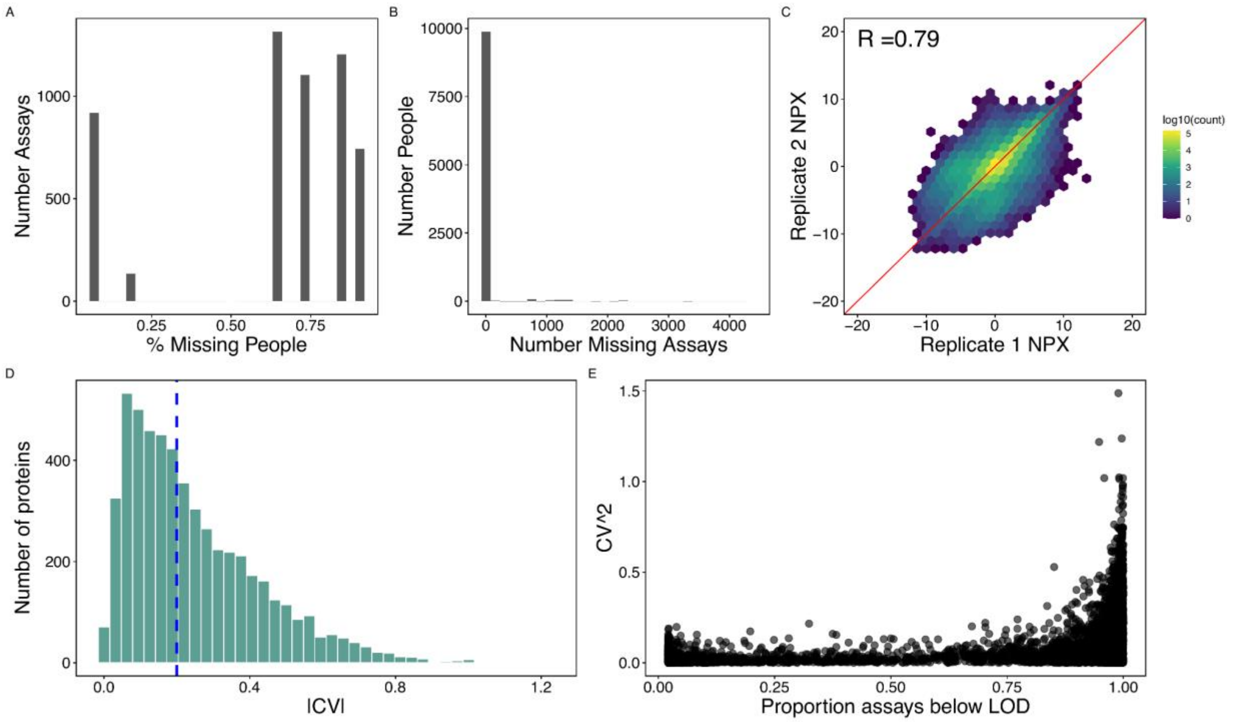


**Supplemental Figure S2. Olink Precision and QC**

Distribution across Olink assays of the fraction of participants with a missing normalized protein expression (NPX) value; each observation is one assay. (B) Distribution across participants of the number of assays with a missing NPX value; each observation is one participant. (C) Measurement reproducibility, assessed using bridge samples for participants run on two separate plates. Hexagonally binned density of NPX for the first replicate (x axis) against the second (y axis); inset text is the Pearson correlation across all pooled pairs (R = 0.79). (D) Distribution across assays of the absolute coefficient of variation (|CV|) between replicate pairs, computed within each bridge participant and summarized as the median across participants; the dashed blue line marks the median across assays. (E) Per-assay $|CV|^2$ against the proportion of samples in which that assay fell below its limit of detection; each point is one assay. Assays detected above LOD in nearly all samples show uniformly low replicate variability, whereas variability rises sharply among assays below LOD in more than ~75% of samples.

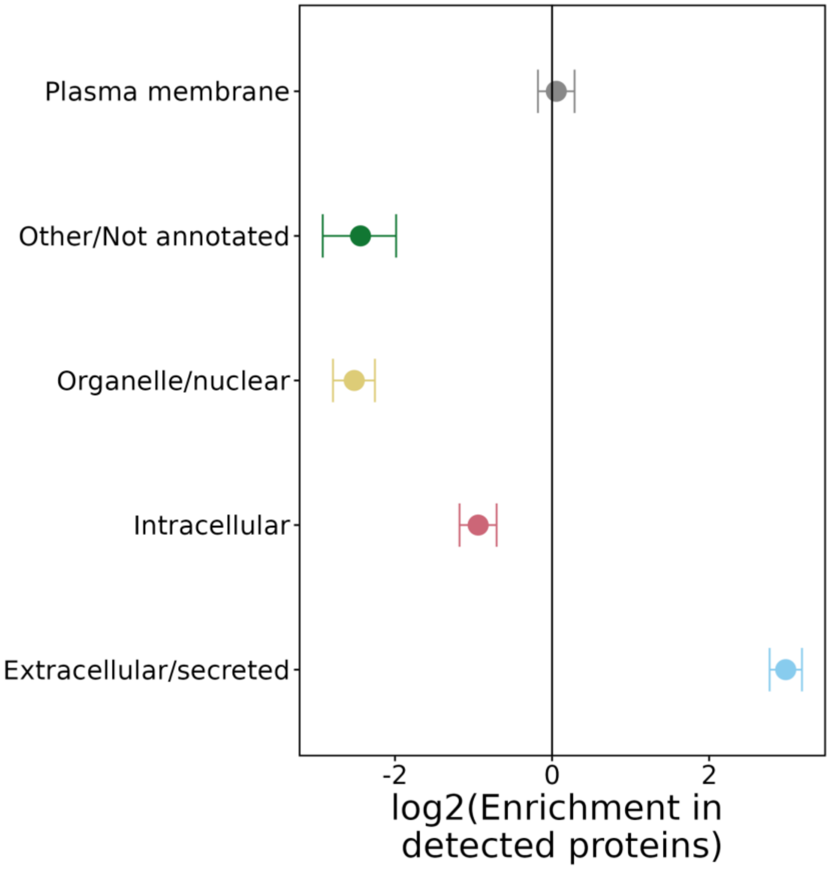


**Supplemental Figure S3. Olink subcellular location enrichment among detected proteins**

For each subcellular location annotated in the Olink Explore HT library content description, a logistic regression was fitted across all assays modelling membership of that location as a function of the proportion of samples in which the protein was measured above its limit of detection. Points give the resulting odds ratio on a log2 scale and horizontal lines the 95% confidence interval; the vertical line marks no association, and positive values indicate enrichment among well-detected proteins. Extracellular and secreted proteins were strongly enriched among well-detected assays (OR 7.8, 95% CI 6.8–9.07, $P < 2.2e-16$), whereas proteins annotated to organelles or the nucleus (OR 0.17) and to the intracellular compartment (OR 0.52) were correspondingly depleted; plasma-membrane proteins showed no association (OR 1.03, $P = 0.651$).

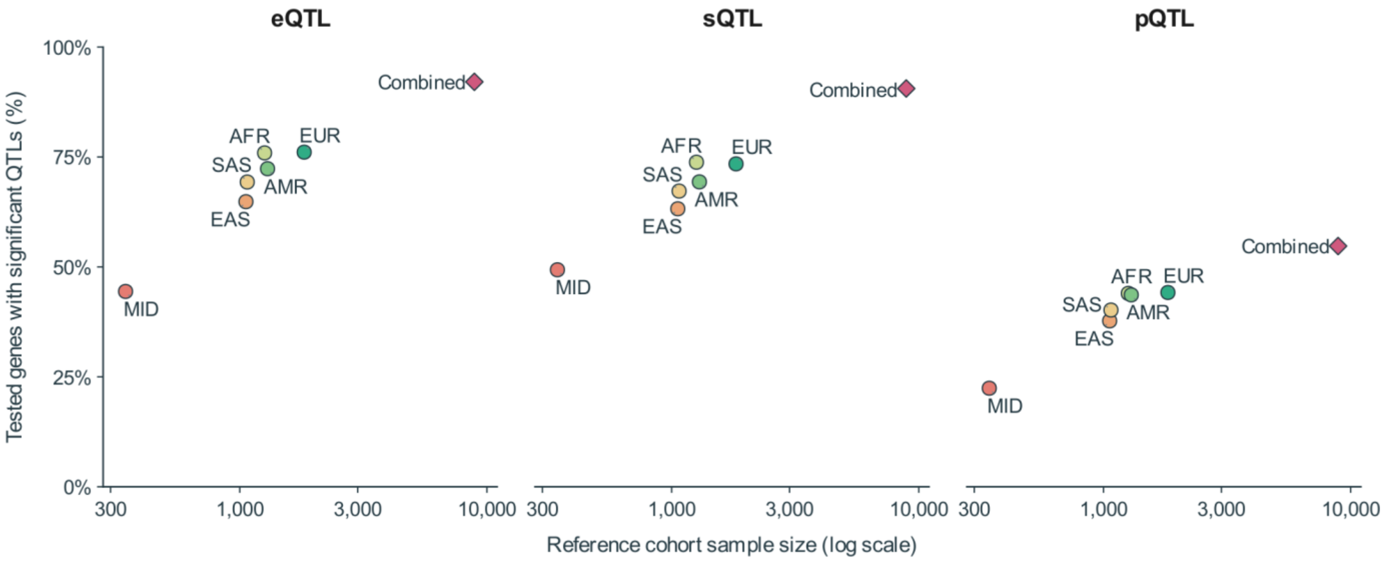


**SUPPLEMENTAL FIGURE S4**

**QTL discovery by population and combined analysis**

**Figure legend**

Left, expression quantitative trait loci (eQTLs); middle, splicing QTLs (sQTLs); right, protein QTLs (pQTLs). Points show the proportion of tested genes with a significant cis-QTL at a Benjamini-Hochberg false discovery rate below 0.05, calculated separately within each population and assay. Circles show population-specific analyses; diamonds show the combined analysis. Combined, 8,895; AFR, 1,260; AMR, 1,295; EAS, 1,059; EUR, 1,822; SAS, 1,072; MID, 345.

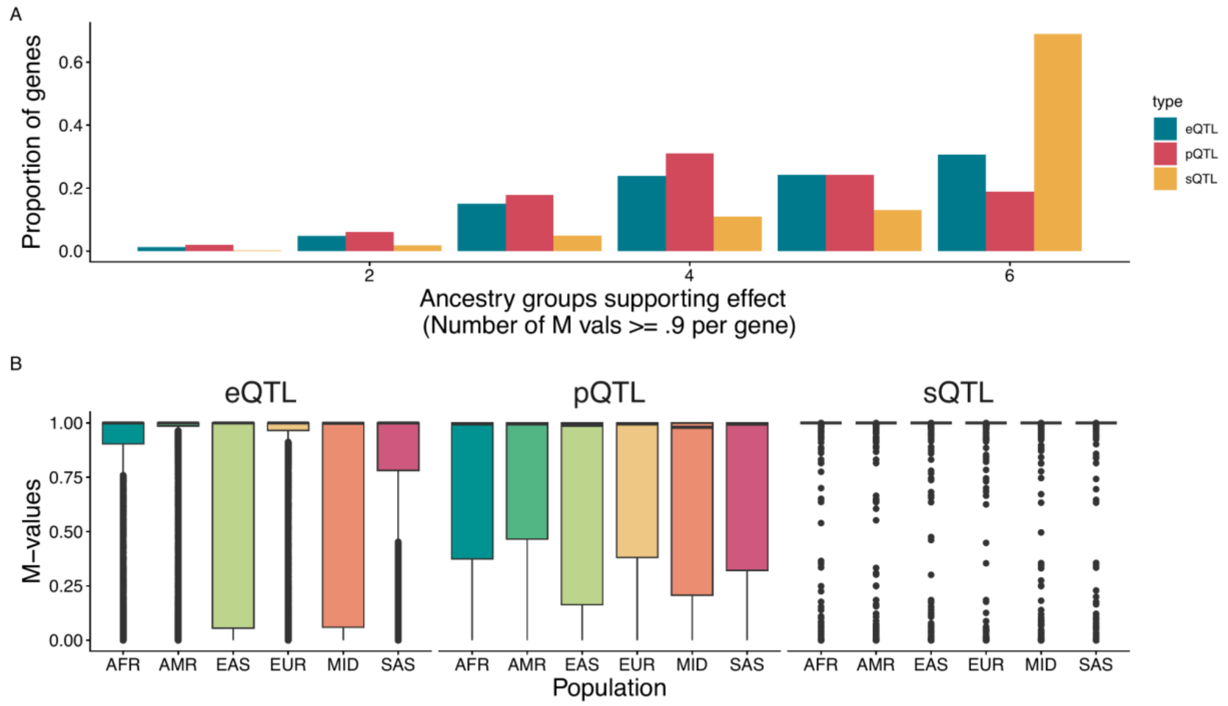


**Supplemental Figure S5. Cross ancestry QTL meta-analysis**

Per-ancestry cis-QTL permutation results were combined across the six genetic-ancestry groups by random-effects meta-analysis (METASOFT), and reproducibility was summarised using M-values, the posterior probability that a QTL effect is present in a given ancestry group. (A) Proportion of genes supported in k ancestry groups, where support is defined as $M \geq 0.9$, shown separately for expression (eQTL), protein (pQTL) and splicing (sQTL) QTLs. Proportions are calculated within each assay, over genes with at least one supporting ancestry group. Splicing QTLs were the most uniformly shared, with the majority of sGenes supported in all six ancestry groups, whereas expression and protein QTLs were more often supported in a subset of groups. (B) Distribution of M-values per ancestry group. Boxes show the median and interquartile range, whiskers extend to 1.5 × IQR, and points beyond are plotted individually. Analyses were restricted to genes with a lead-variant effect estimate in all six ancestry groups, and M-values are defined by MetaSoft only for genes whose meta-analysis reaches $P < 1 \times 10^{-7}$.

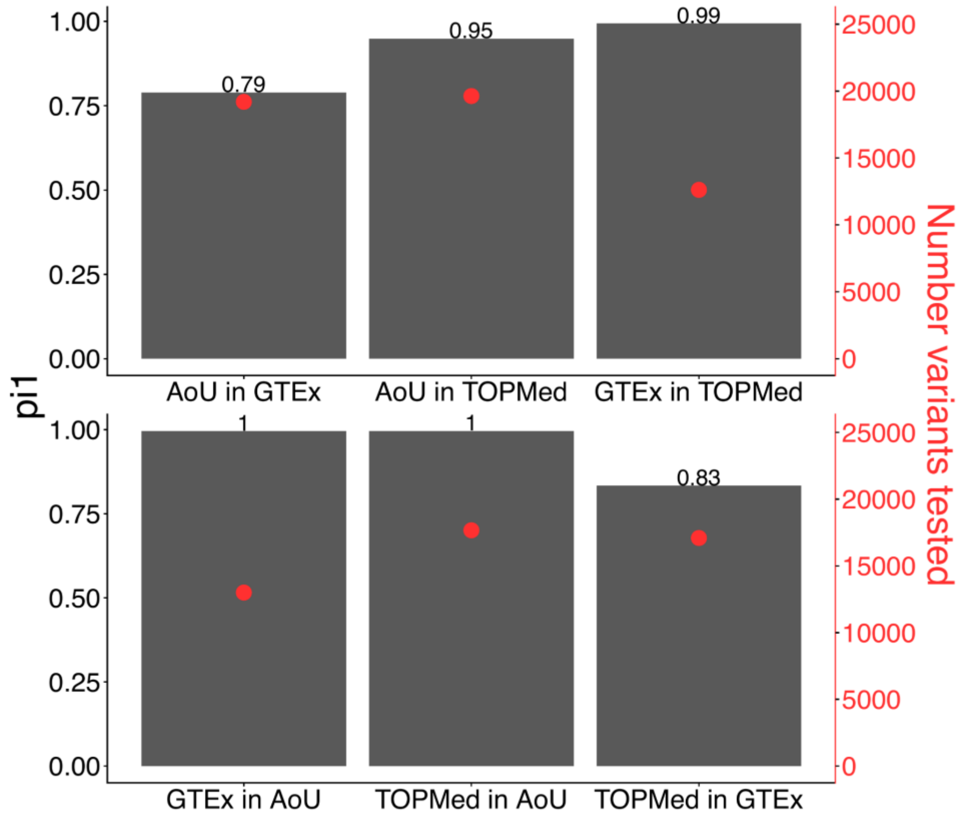


**Supplemental Figure S6. eQTL replication with external consortia**

Storey's $\pi_1$ statistic quantifying the proportion of eGenes discovered in one dataset that show a replication signal in another, for all six ordered pairs of the three datasets (All of Us, GTEx and TOPMed). Each panel label reads "discovery in replication": for example, "AoU in GTEx" is the fraction of All of Us eGenes that replicate in GTEx. Columns pair reciprocal comparisons  column 1, All of Us ↔ GTEx; column 2, All of Us ↔ TOPMed; column 3, GTEx ↔ TOPMed  with the top row showing one direction and the bottom row the reverse. Gray bars and the numbers above them give $\pi_1 = 1 - \hat{\pi}_0$, estimated from the distribution of replication-dataset gene-level permutation P values across the discovery eGenes (left axis). Red points give the number of discovery eGenes entering each estimate (right axis). Analyses were restricted to genes tested in both datasets of a pair; discovery eGenes were defined at a permutation q value < 0.05.

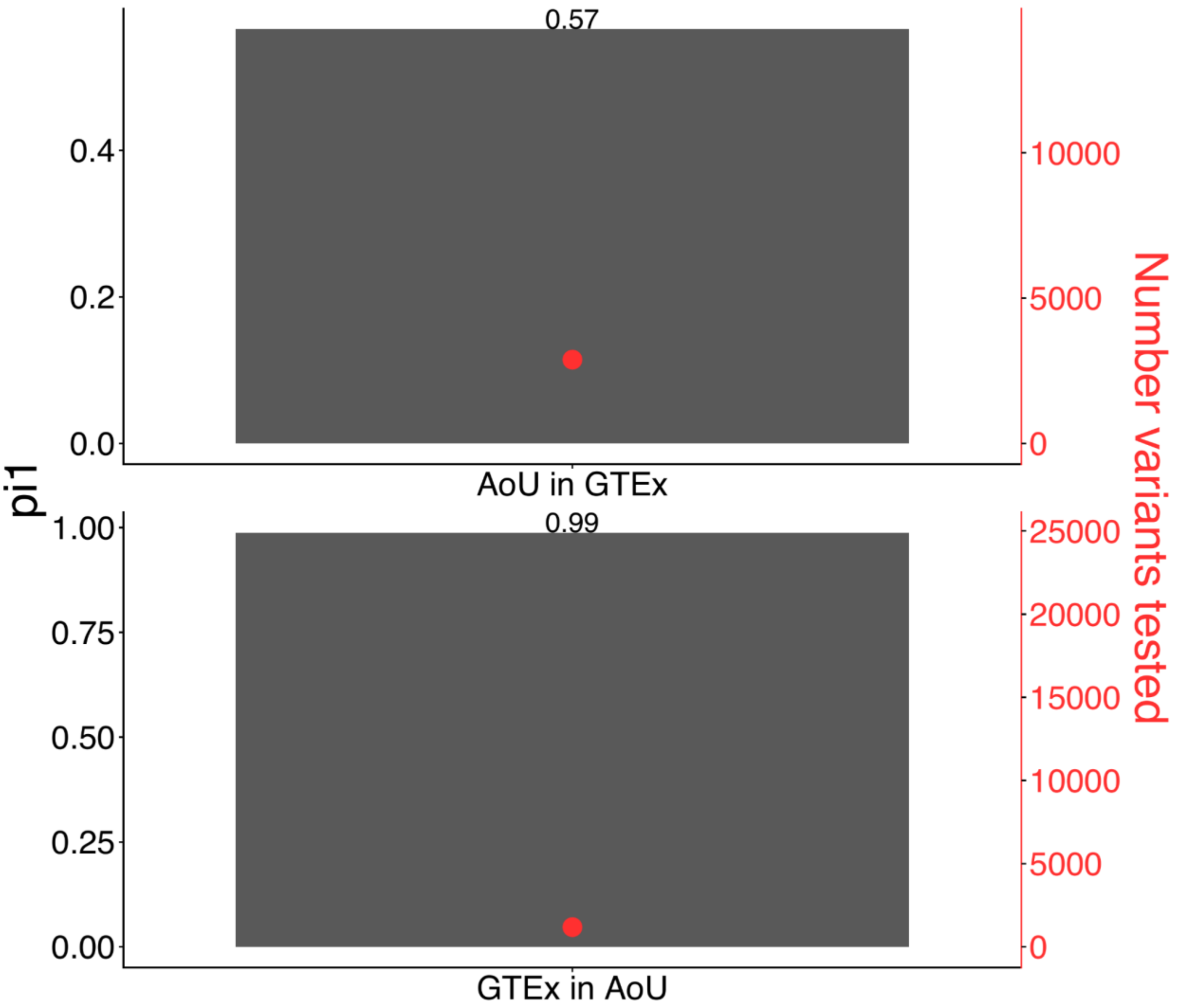


**Supplemental Figure S7. sQTL replication with external consortia**

Storey's $\pi_1$ statistic quantifying the proportion of sGenes discovered in one dataset that show a replication signal in another, for all six ordered pairs of the two datasets (All of Us, GTEx). Each panel label reads "discovery in replication": for example, "AoU in GTEx" is the fraction of All of Us eGenes that replicate in GTEx. Columns pair reciprocal comparisons column 1, All of Us ↔ GTEx. Gray bars and the numbers above them give $\pi_1 = 1 - \hat{\pi_0}$, estimated from the distribution of replication-dataset gene-level permutation P values across the discovery sGenes (left axis). Red points give the number of discovery eGenes entering each estimate (right axis).

Analyses were restricted to genes tested in both datasets of a pair; discovery sGenes were defined at a permutation q value < 0.05.

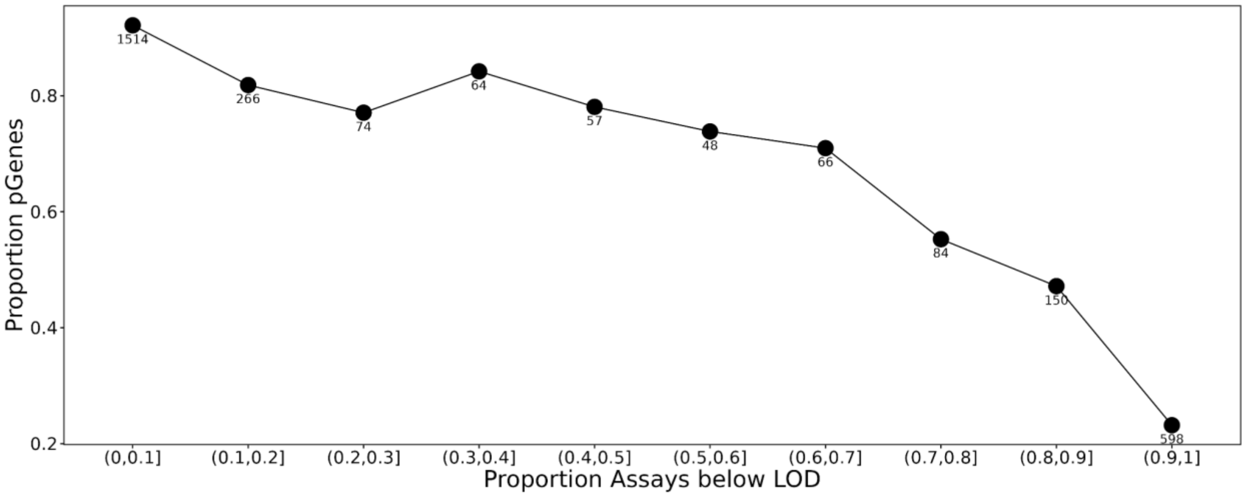


**Supplemental Figure S8. pQTL detection rate by LOD**

For each Olink assay, the proportion of samples whose NPX value fell below that assay's limit of detection (LOD) was computed across all participants, and assays were grouped into ten equal-width bins of this proportion. Points give the proportion of proteins in each bin with a significant cis-pQTL (pGene), defined as a permutation q-value < 0.05 in the combined-ancestry tensorQTL cis analysis; numbers beside each point give the number of proteins contributing to that bin. Proteins measured above LOD in nearly all samples were the most likely to have a detectable cis-pQTL (96.5% in the (0, 0.1] bin), falling to 26% among proteins below LOD in more than 90% of samples. Sensitivity of the proteomic assay is therefore a determinant of cis-pQTL discovery.

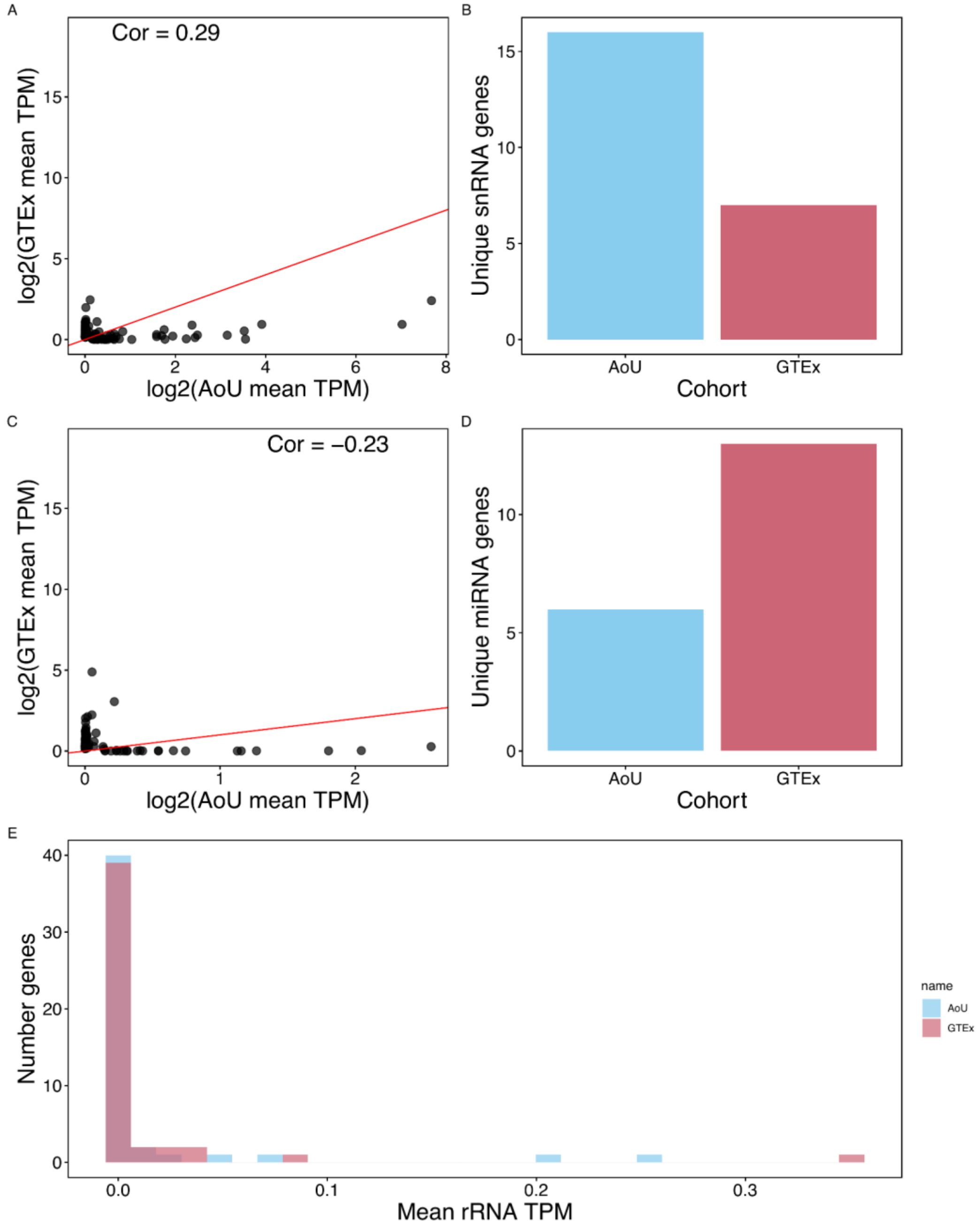

A
Cor = 0.29
log2(GTEx mean TPM)
log2(AoU mean TPM)
B
Unique snRNA genes
AoU
GTEx
Cohort
C
Cor = −0.23
log2(GTEx mean TPM)
log2(AoU mean TPM)
D
Unique miRNA genes
AoU
GTEx
Cohort
E
Number genes
Mean rRNA TPM
name
AoU
GTEx

**Supplemental Figure S9. GTEx noncoding RNA comparison**

Mean expression per gene was computed across all samples in each cohort as transcripts per million (TPM), genes were matched between cohorts, and gene biotype was assigned from the GENCODE v45 annotation using 5000 AoU samples aligned to GENCODE v45. (A) Mean expression of small nuclear RNA (snRNA) genes in All of Us (AoU; x axis, log2(mean TPM + 1)) against GTEx whole blood (y axis, log2(mean TPM + 1)), restricted to snRNAs with a mean TPM greater than 0.1 in at least one cohort. Each point is one gene and the red line is the identity (y = x); inset text gives the Pearson correlation between the two log-transformed means (r = 0.29). (B) Number of the snRNA genes in A detected in exactly one cohort, defined as a mean TPM greater than 1 in one cohort and below 1 A. Sixteen snRNAs were specific to All of Us and seven to GTEx. (C, D) As in A and B, for microRNA (miRNA) genes (r = −0.23); six were specific to All of Us and thirteen to GTEx. (E) Distribution of mean ribosomal RNA (rRNA) gene TPM in each cohort, no expression filter was applied, and essentially all rRNA genes fall below 0.1 TPM in both cohorts.

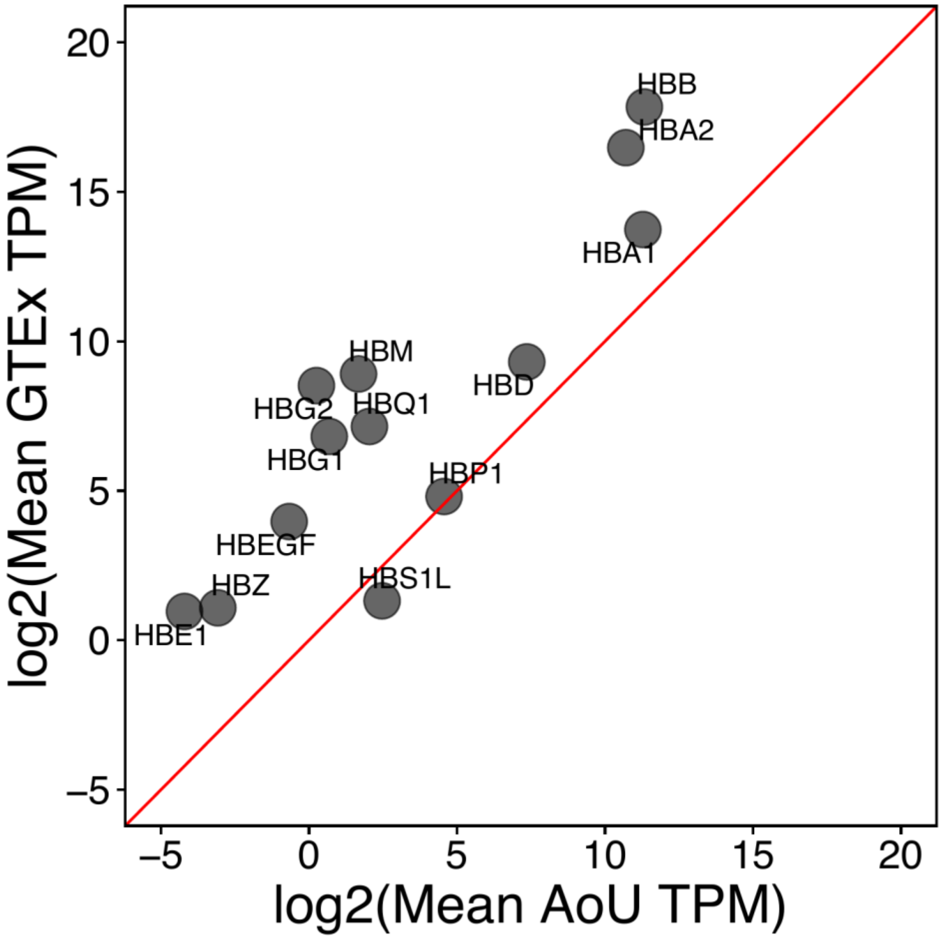


**Supplemental Figure S10. GTEx hemoglobin expression comparison**

Mean expression per gene was computed across all samples in each cohort as transcripts per million (TPM), genes were matched between cohorts on the Ensembl gene. Hamegoblin subunit genes (n = 13) are plotted as log2 mean TPM in All of Us (AoU; x axis) against GTEx whole blood (y axis); each point is one gene, labelled with its symbol, and the red line is the identity (y = x).

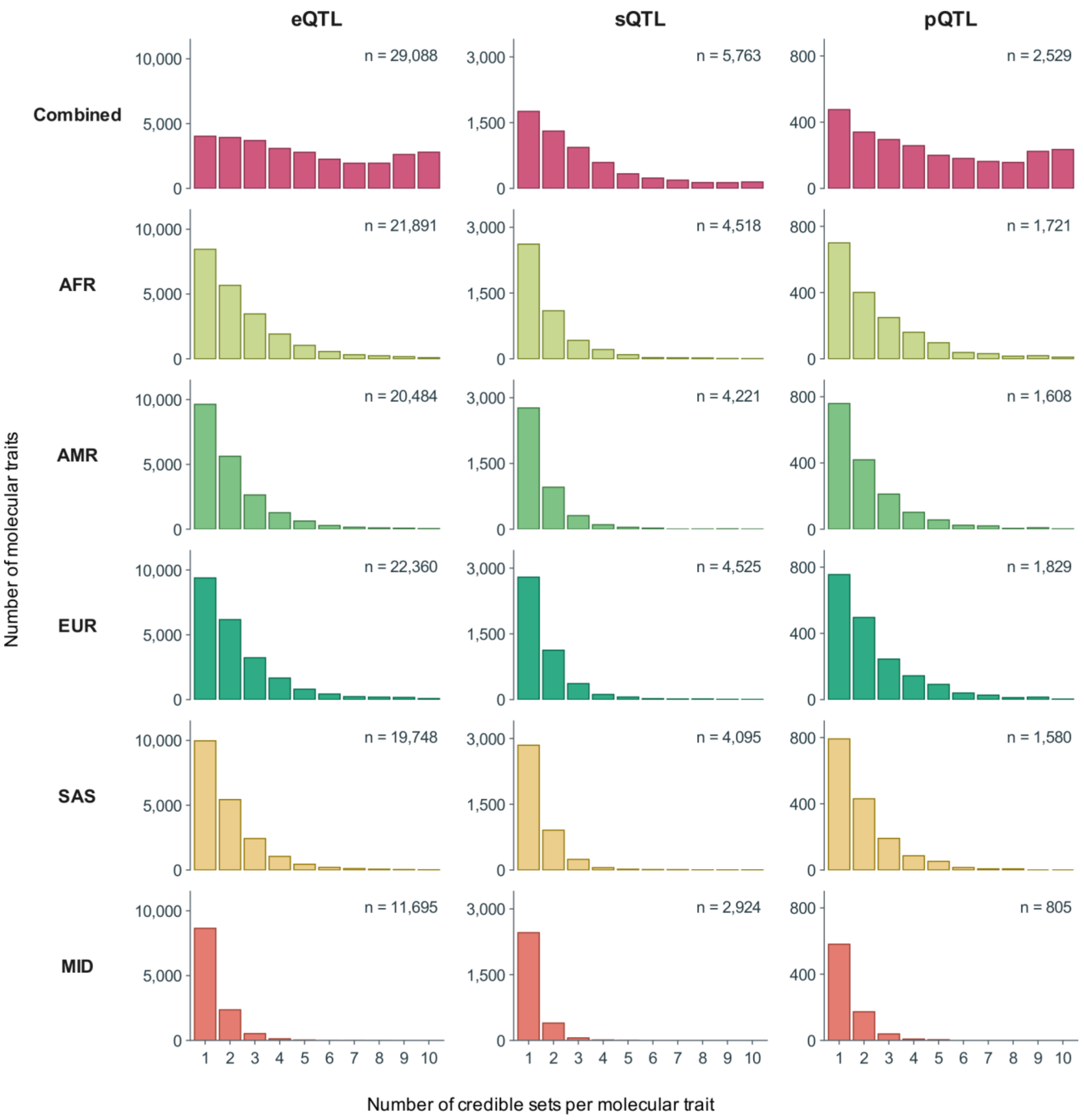


**SUPPLEMENTAL FIGURE S11 | FIGURE LEGEND**

**Credible sets per molecular trait by population and combined analysis**

Histograms show the number of credible sets per molecular trait for expression quantitative trait loci (eQTLs), splicing QTLs (sQTLs), and protein QTLs (pQTLs).

Each bar gives the number of molecular traits with the indicated number of distinct credible sets. The full observed range of 1-10 sets is shown. The n label gives the number of traits with at least one credible set in that panel.

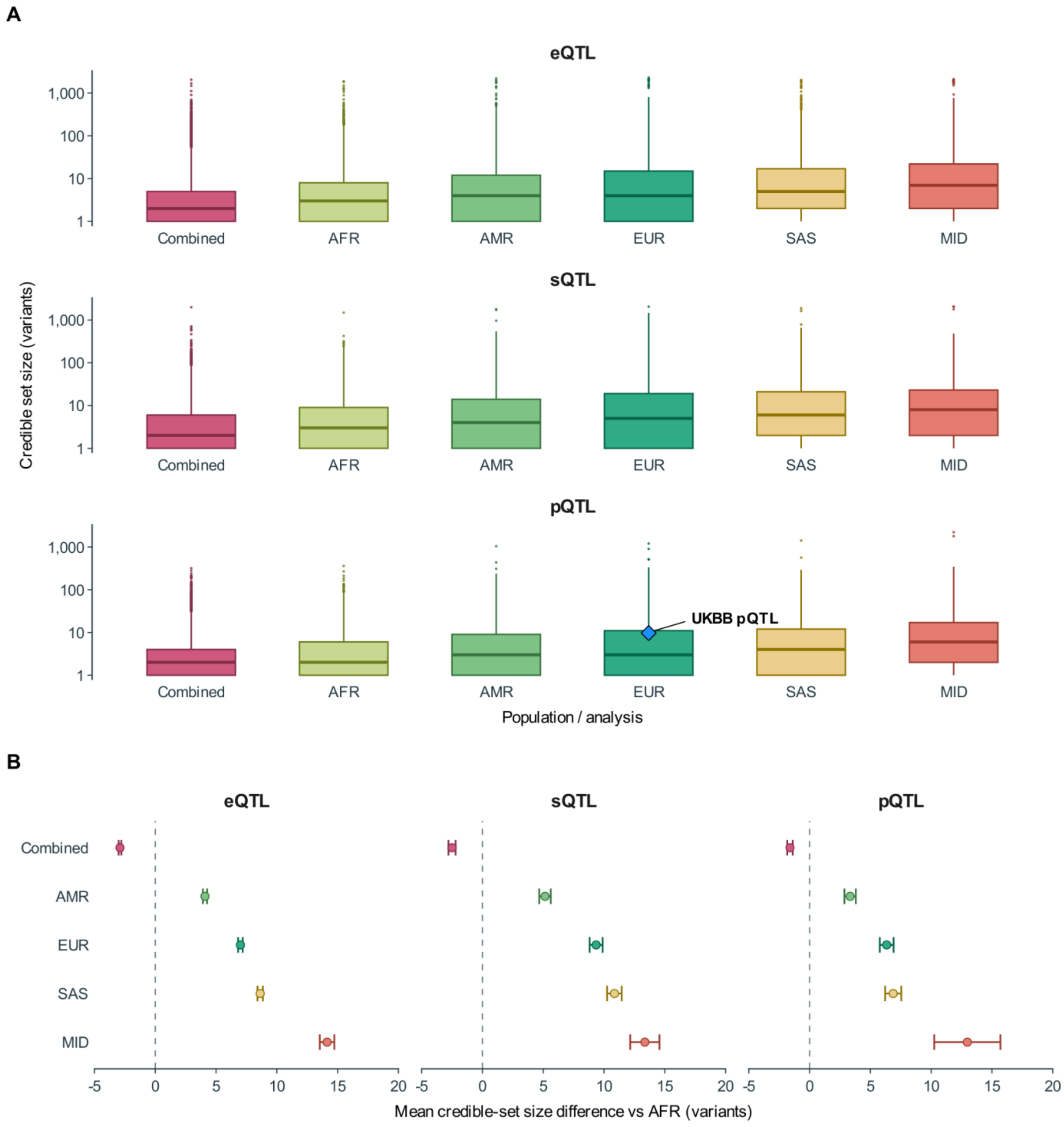
A
eQTL
sQTL
pQTL
Credible set size (variants)
1,000
100
10
1
Combined
AFR
AMR
EUR
SAS
MID
UKBB pQTL
Population / analysis
B
eQTL
sQTL
pQTL
Combined
AMR
EUR
SAS
MID
-5
0
5
10
15
20
Mean credible-set size difference vs AFR (variants)

### SUPPLEMENTAL FIGURE S12 | FIGURE LEGEND

#### Credible-set sizes and differences from AFR

(A) Credible-set-size distributions for expression QTLs (eQTLs), splicing QTLs (sQTLs), and protein QTLs (pQTLs), for the combined analysis and five population groups. Boxes show the median and interquartile range of log10-transformed credible-set sizes; whiskers extend to the most extreme value within 1.5 times the interquartile range on that scale. Points beyond the whiskers show outliers. The blue diamond above EUR in the pQTL plot is the retained UK Biobank (UKBB) reference mean of 9.7 variants.

(B) Difference in arithmetic mean credible-set size between each analysis group and AFR, in variant counts. Points are group mean minus AFR mean; horizontal bars show plus or minus one standard error (SE). The dashed line marks the AFR reference of zero. Negative values indicate smaller mean credible sets than AFR.

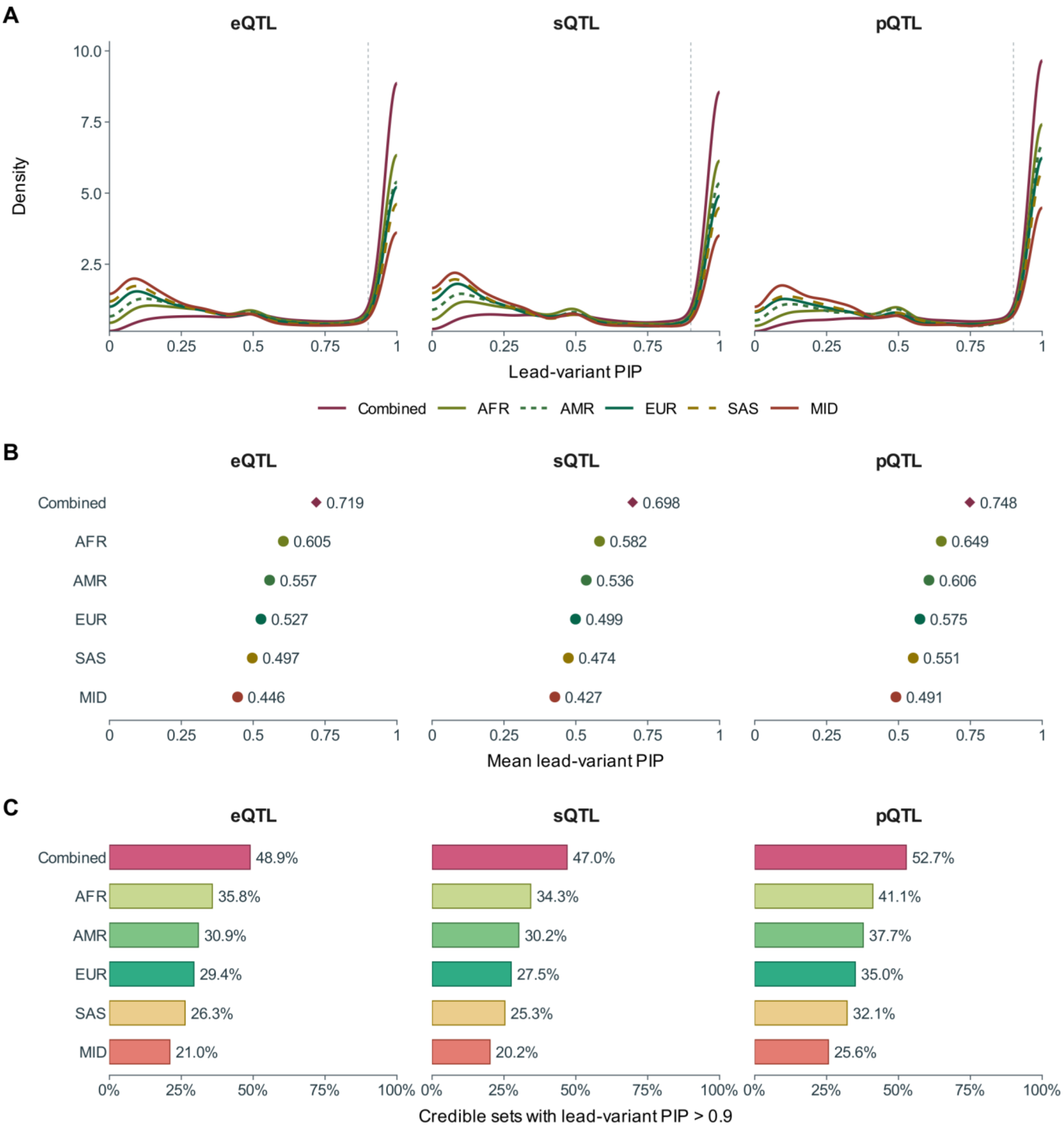
A
eQTL
sQTL
pQTL
Density
10.0
7.5
5.0
2.5
0
0.25
0.5
0.75
1
Lead-variant PIP
Combined
AFR
AMR
EUR
SAS
MID
B
eQTL
sQTL
pQTL
Combined
AFR
AMR
EUR
SAS
MID
0.719
0.605
0.557
0.527
0.497
0.446
0.698
0.582
0.536
0.499
0.474
0.427
0.748
0.649
0.606
0.575
0.551
0.491
Mean lead-variant PIP
C
eQTL
sQTL
pQTL
Combined
AFR
AMR
EUR
SAS
MID
48.9%
35.8%
30.9%
29.4%
26.3%
21.0%
47.0%
34.3%
30.2%
27.5%
25.3%
20.2%
52.7%
41.1%
37.7%
35.0%
32.1%
25.6%
0%
25%
50%
75%
100%
Credible sets with lead-variant PIP > 0.9

**SUPPLEMENTAL FIGURE S13 | FIGURE LEGEND**

**Lead-variant PIP and causal resolution across analyses**

(A) Distributions of lead-variant posterior inclusion probability (PIP) for expression quantitative trait loci (eQTLs), splicing QTLs (sQTLs), and protein QTLs (pQTLs). Curves identify Combined, AFR, AMR, EUR, SAS, and MID analyses. Lead PIP is the maximum variant PIP within each credible set. Each distinct set contributes one value. The gray dashed line marks PIP = 0.9.

(B) Arithmetic mean lead-variant PIP for each analysis group and QTL type. Each credible set has equal weight. Numbers give means rounded to three decimal places. Diamonds show Combined; circles show populations.

(C) Percentage of credible sets with a causally resolved variant, defined here as a lead-variant PIP strictly greater than 0.9. The denominator is all credible sets in that group and QTL type. Labels give percentages.

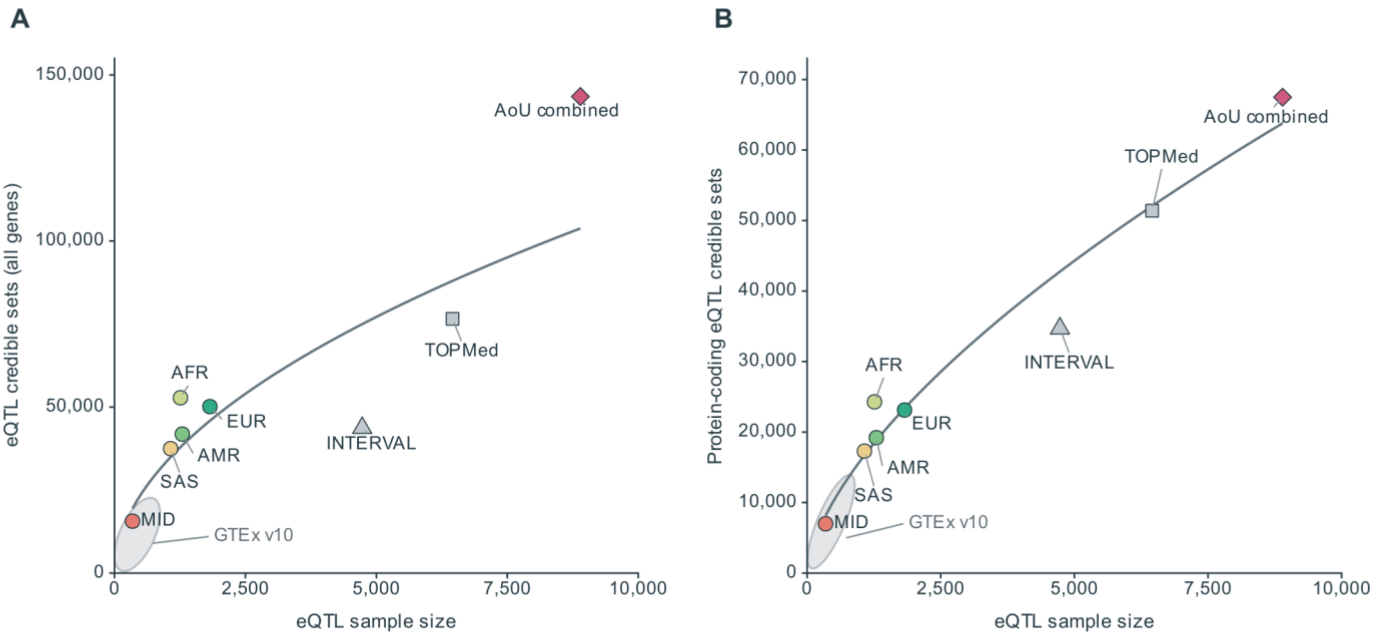


**SUPPLEMENTAL FIGURE S14**

**eQTL credible-set discovery across sample sizes**

**Figure legend**

(A) Total expression quantitative trait locus (eQTL) credible-set discovery across all available target genes. (B) Discovery within 10,881 protein-coding genes with at least one credible set in each of the All of Us (AoU) combined, TOPMed, and INTERVAL analyses. Protein-coding genes were defined using GENCODE v48. Gray squares and triangles show TOPMed (n = 6,454) and INTERVAL (n = 4,729), respectively. The light gray regions enclose the 50 GTEx v10 tissue-analysis positions (n = 77-816 per tissue), with 2% padding in display coordinates.

B

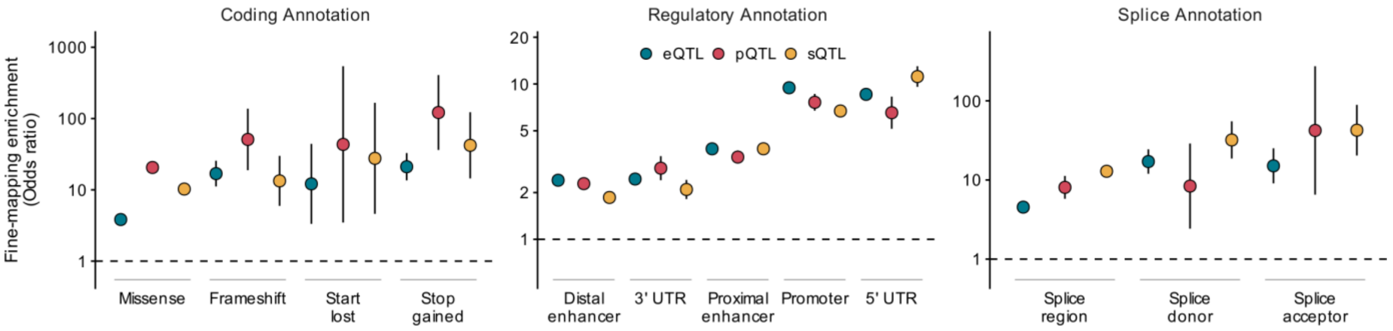


**SUPPLEMENTAL FIGURE S15**

**Fine-mapping annotation enrichment**

**Figure legend**

Fine-mapping enrichment for coding, regulatory, and splice annotations in eQTLs, pQTLs, and sQTLs. Each annotation is modeled separately within each assay by logistic regression of annotation status on variant posterior inclusion probability (PIP). Points show exp(beta_PIP), the modeled odds ratio for the contrast between PIP = 1 and PIP = 0. Vertical intervals show exp(beta_PIP plus or minus 1.96 times its standard error), giving 95% Wald confidence intervals. Dashed horizontal lines mark an odds ratio of one.

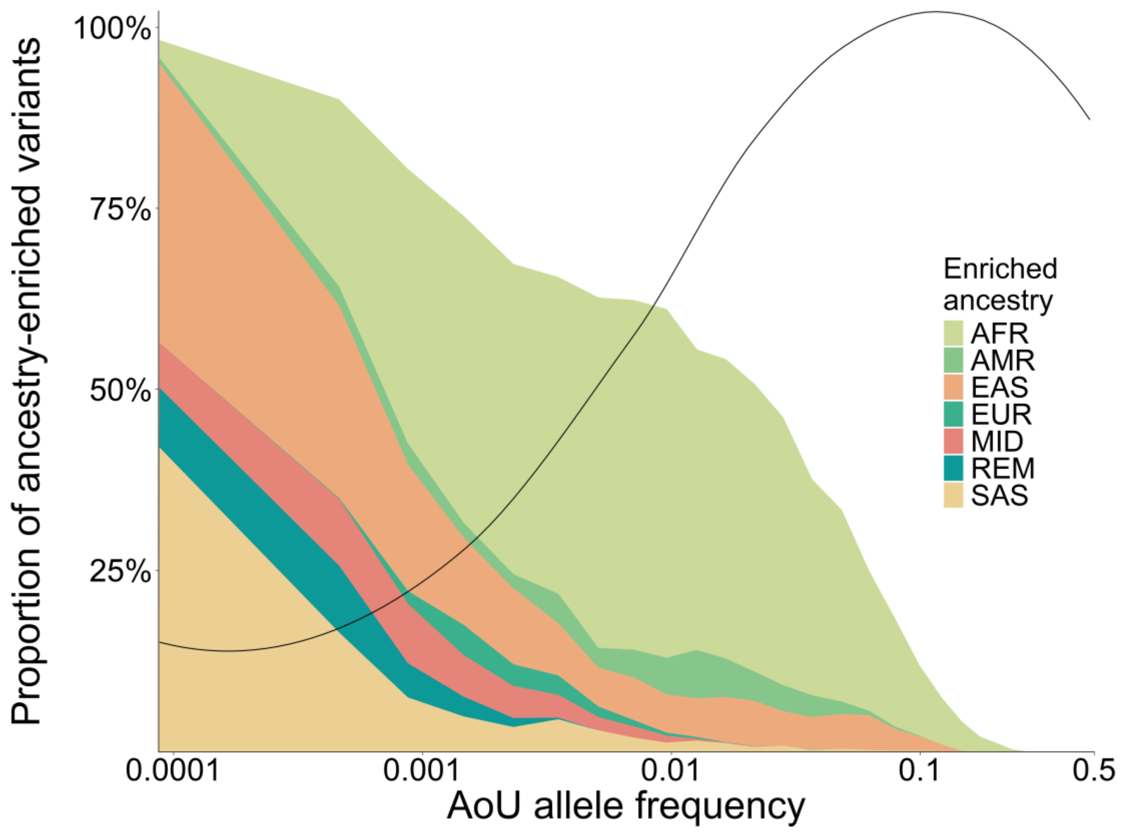


**Supplemental Figure S16. AoU-cohort wide allele frequency by ancestry enrichment**

Fine-mapped causal variants (SuSiE PIP > 0.9) were divided into 30 bins of equal size by their minor allele frequency in the full All of Us cohort, and each bin is plotted at the geometric mean of its allele-frequency range (x axis, log10 scale). Stacked areas show the proportion of variants in each bin that are ancestry-enriched  defined, as in Figure 2B, as an ancestry-enrichment odds ratio greater than 5 with an FDR-adjusted P < 0.05  coloured by the ancestry group in which the variant is enriched (AFR, AMR, EAS, EUR, MID, SAS and REM), so that the total height of the stack is the overall proportion of variants in that bin that are ancestry-enriched. The black line, plotted on the same y axis, is the proportion of variants in each bin observed at non-zero allele frequency in all seven ancestry groups, smoothed with a centred five-bin rolling mean and then with a LOESS fit. n = 69,931 fine-mapped expression variants across the 30 plotted bins. Of the variants plotted, 23,519 (33.6%) were ancestry-enriched, most often in AFR (14,311), followed by EAS (4,001), SAS (2,076), AMR (1,196), MID (974), REM (615) and EUR (346).

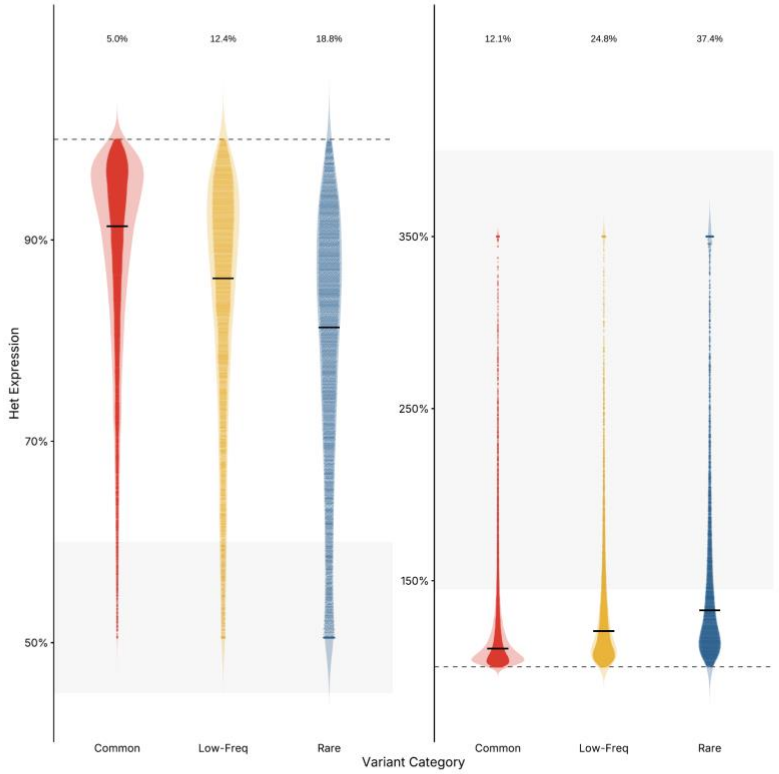


**Supplemental Figure S17. Strongest aFC by allele frequency per gene**

For each gene, the fine-mapped variant with the strongest allelic fold change (aFC) was selected and converted to an expected heterozygous expression level, (1 + 2^log2 aFC) / 2, expressed as a percentage of the population-typical level, so that 100% denotes no effect. Genes are grouped on the x axis by the allele-frequency class of that strongest-effect variant  common (MAF > 5%), low-frequency(MAF >1% and MAF < 5%) and rare (MAF < 1%) and shown separately for variants that decrease expression (left) and increase it (right). Violins show the distribution with individual genes overlaid as points, and the black horizontal bar marks the median. The dashed line marks 100%, i.e. no change. The shaded band marks the dosage-like range: 45–60% of expected expression on the left, equivalent to the effect of a heterozygous deletion, and 145–400% on the right, equivalent to a heterozygous duplication. Percentages above each violin give the proportion of genes in that frequency class whose strongest-effect variant falls within the shaded band. Genes whose strongest variant is rare were substantially more likely to reach a dosage-like state than those with a common strongest variant, in both directions: 18.8% versus 5.0% for expression-decreasing variants, and 37.4% versus 12.1% for expression-increasing variants, with low-frequency variants intermediate in both cases (12.4% and 24.8%).

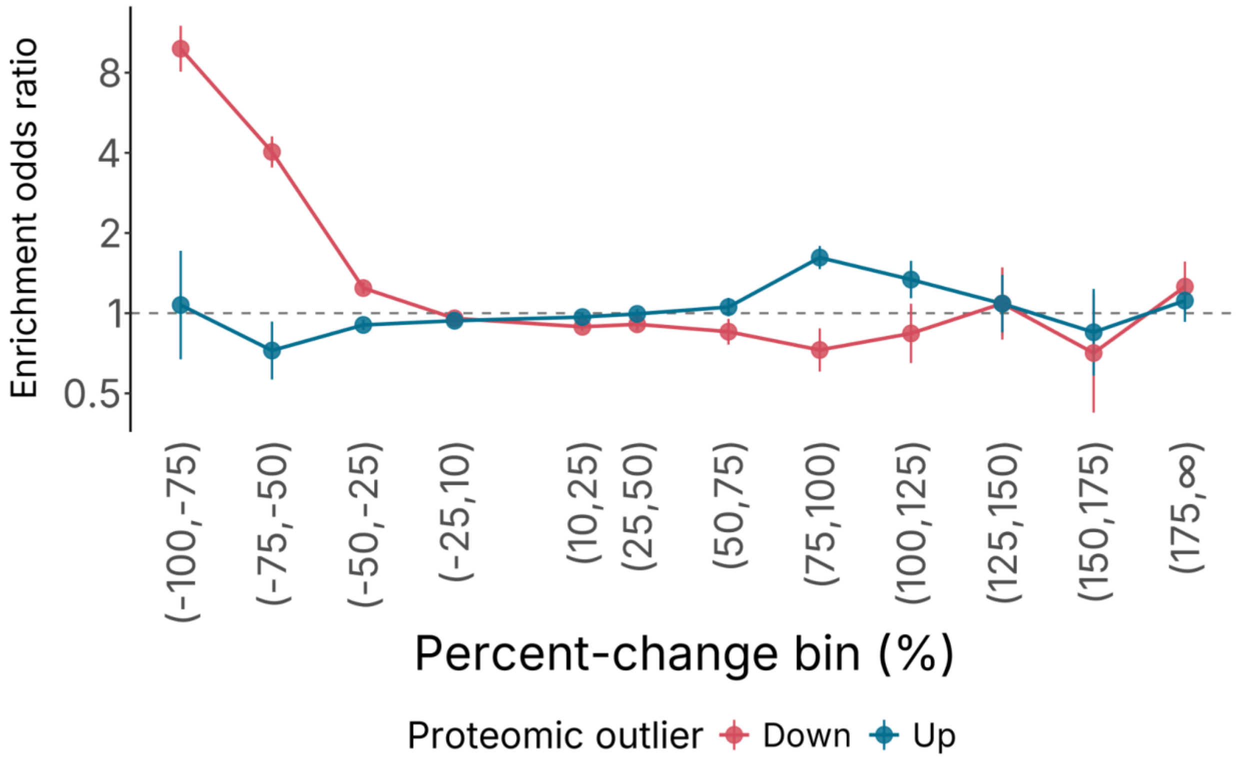


**Supplemental Figure S18. Extreme QTL predictions are enriched for protein underliers**

Proteomic outlier enrichment across predicted percent-change bins. Points show odds ratios for under (red) and over (blue) proteomic outliers compared to a reference bin (-10% to 10% predicted change); vertical bars indicate the 95% confidence intervals. The dashed horizontal line marks an odds ratio of 1, corresponding to no enrichment. Bin labels show the predicted percent-change ranges, with the final bin representing values above 175%. Proteomic outliers were defined using protein-abundance z-scores for each protein–sample pair. Samples with a z-score > 4 were classified as overliers, whereas samples with a z-score < −4 were classified as underliers.

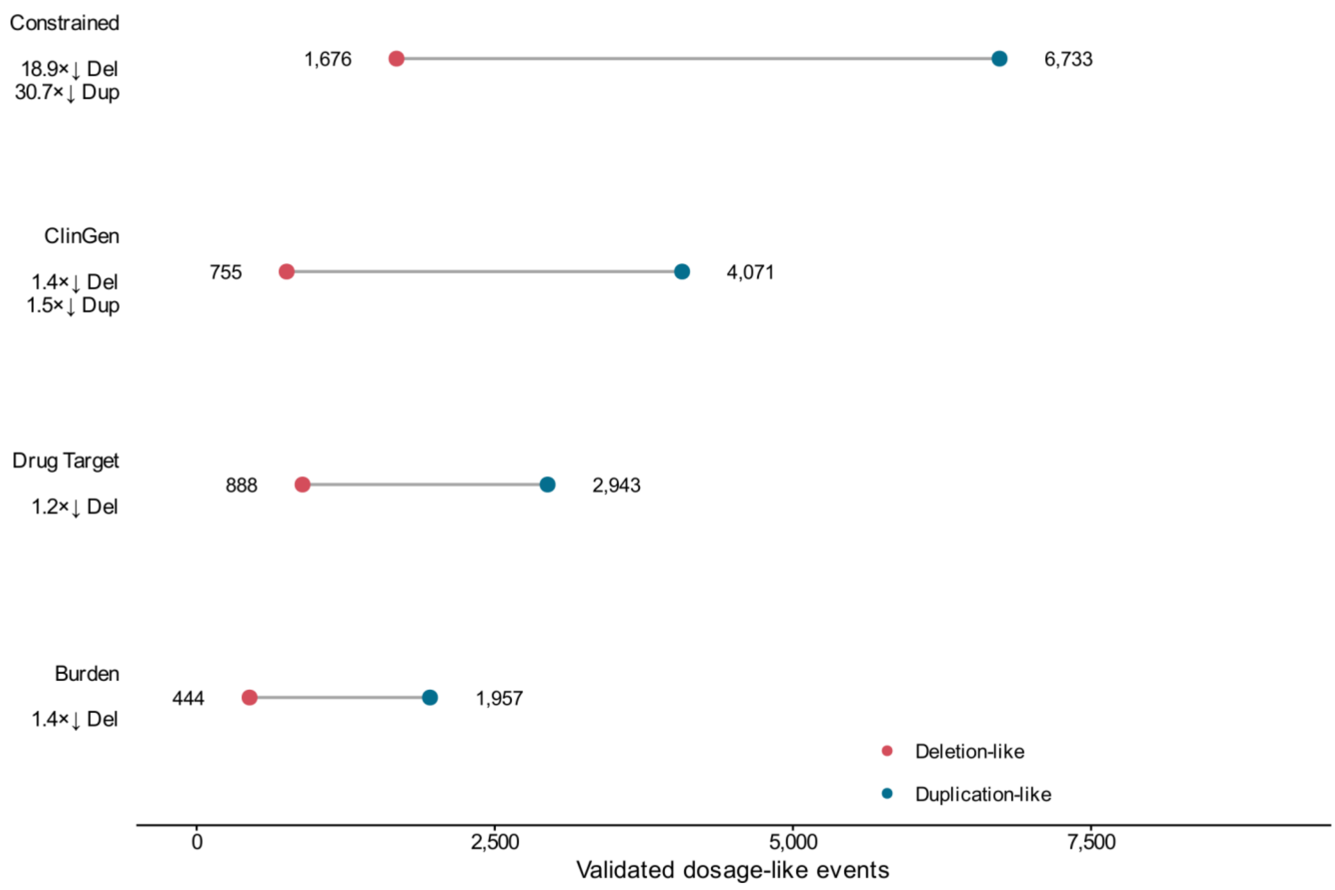


**SUPPLEMENTAL FIGURE S19**

**Validated dosage-like events by gene category**

**Figure legend**

Counts of validated dosage-like events across the Constrained, ClinGen, Drug Target, and Burden gene categories. Red points show expression-underlier, deletion-like events; blue points show expression-overlier, duplication-like events. Gray segments join the two counts within each category. The number of events is printed beside each point. The category labels retain the fold-reduction annotations for deletion-like (Del) and duplication-like (Dup) events calculated from an incidence rate ratio, enrichments are only shown when a category-dosage like event pair is significant enriched or depleted (FDR < .05).

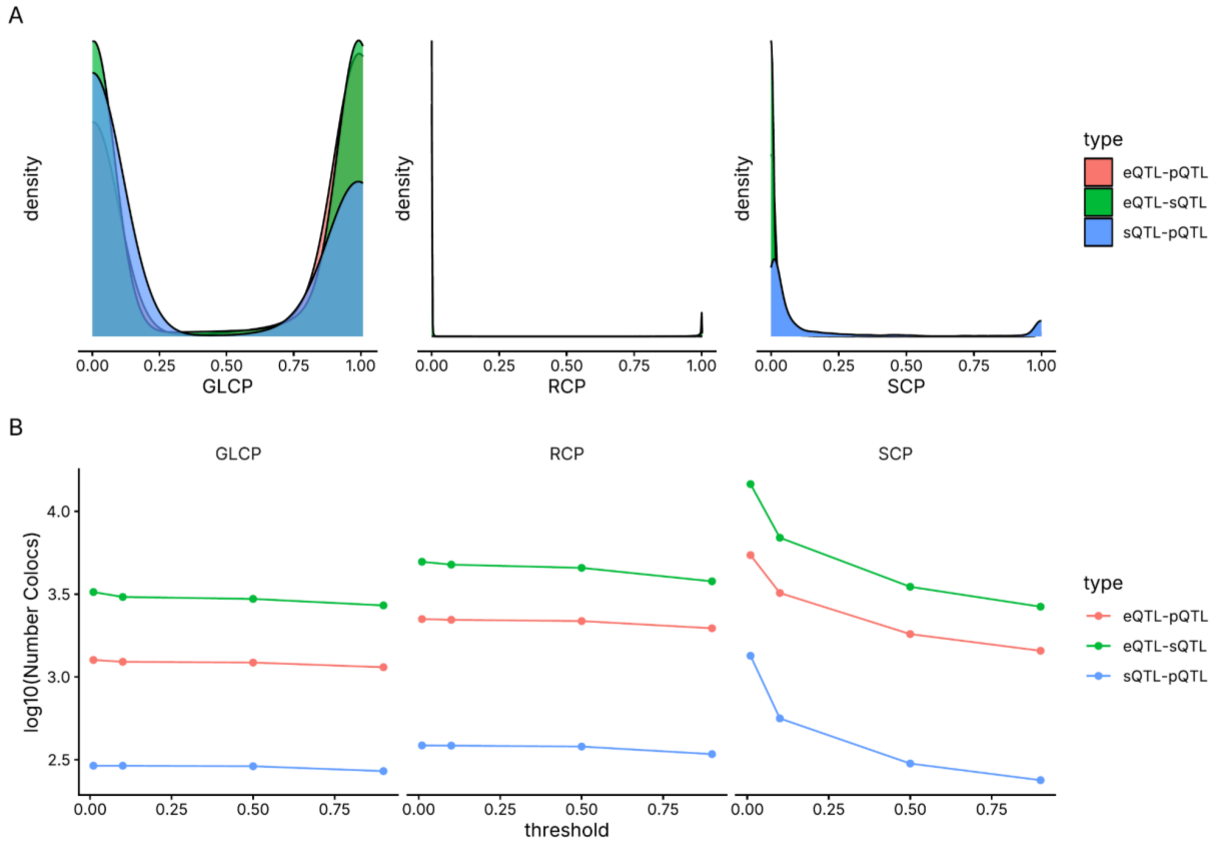


**Supplemental Figure S20. Cross-QTL colocalization is robust across thresholds**

Kernel density estimates of the FastENLOC colocalization posteriors, coloured by assay pair (eQTL–pQTL, eQTL–sQTL, sQTL–pQTL), at each of the three levels at which FastENLOC reports colocalization: the gene-level colocalization probability (GLCP; one record per gene), the regional colocalization probability (RCP; one record per credible-set pairing) and the SNP-level colocalization probability (SCP; one record per variant). (B) Number of colocalizations recovered as a function of an arbitrary posterior cutoff.

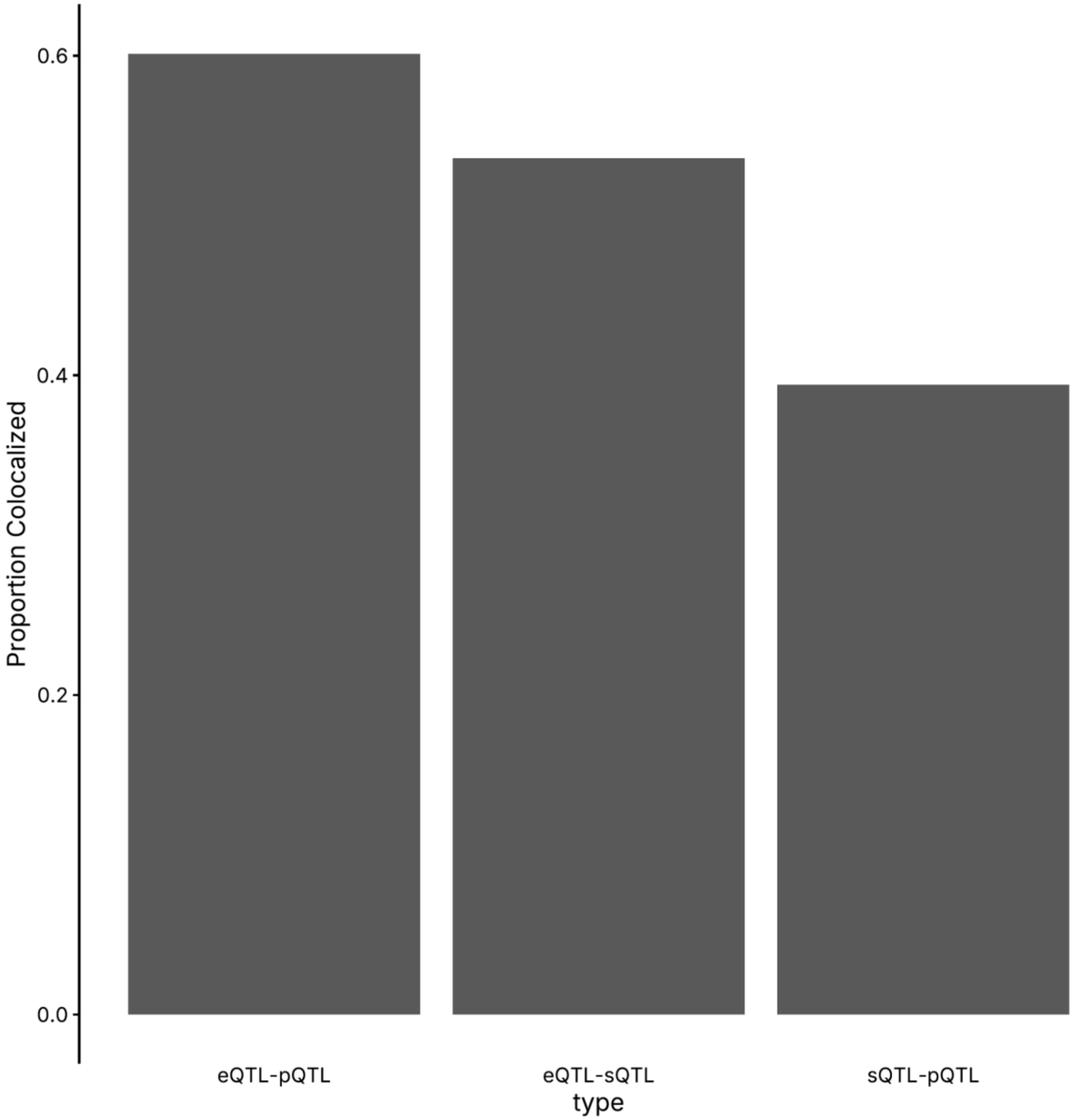


**Supplemental Figure S21. Coloc rate by molecular pair**

Bars give, for each assay pair, the proportion of gene-level FastENLOC records called colocalized at a Bayesian FDR of 5%. The denominator is every gene tested for that pair, unlike Figure 3A genes are not restricted to those fine-mapped in all three assays. Between 39.4% and 60.2% of tested genes colocalized depending on the pair: eQTL–pQTL 60.2% (1,265 of 2,102), eQTL–sQTL 53.6% (3,060 of 5,709) and sQTL–pQTL 39.4% (298 of 756). Pooled across all three pairs the rate is 54.0% (4,623 of 8,567).

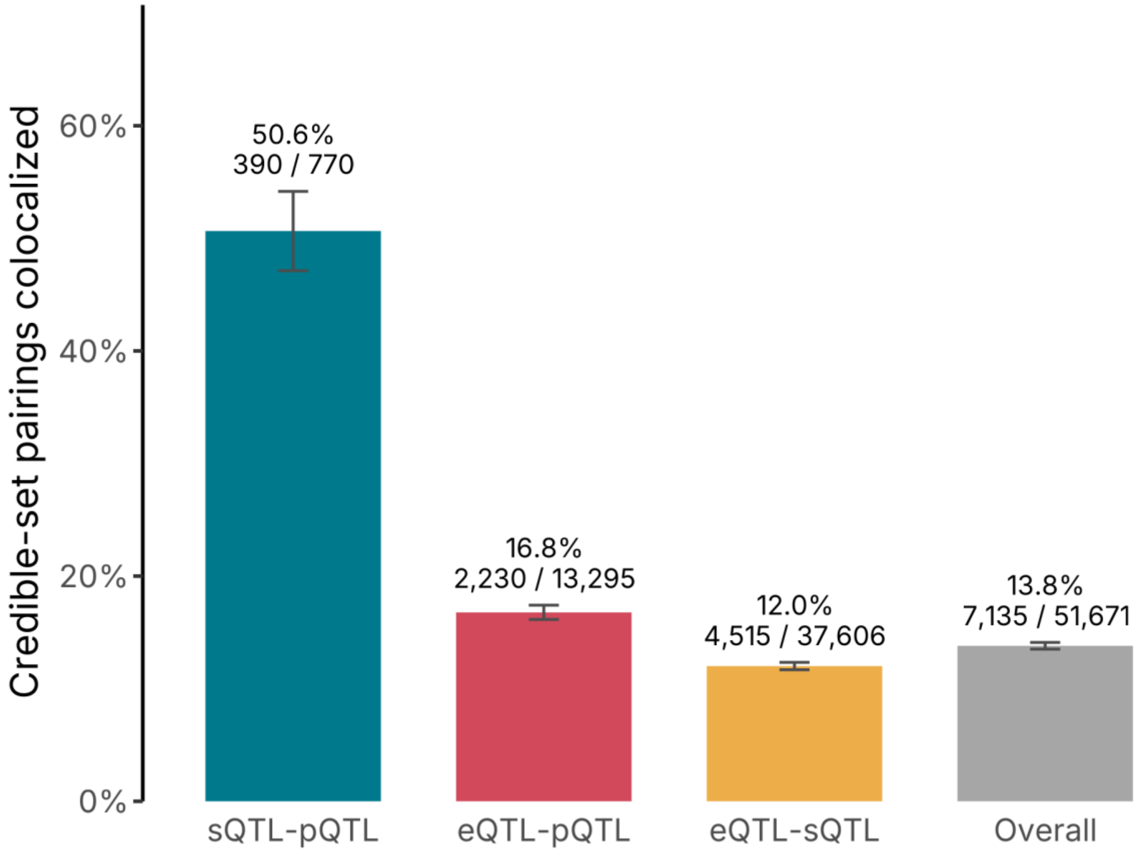


**Figure S22. Colocalization between specific credible sets is far more restricted than sharing at the gene level.** For each pair of molecular phenotypes, bars give the percentage of tested credible-set pairings called colocalized; the grey bar pools all three pairs. Labels gives the percentage and the colocalized/tested counts, and error bars are 95% confidence intervals. Sharing is highest for sQTL–pQTL (50.6%, 390/770), followed by eQTL–pQTL (16.8%, 2,230/13,295) and eQTL–sQTL (12.0%, 4,515/37,606), with an overall rate of 13.8% (7,135/51,671).

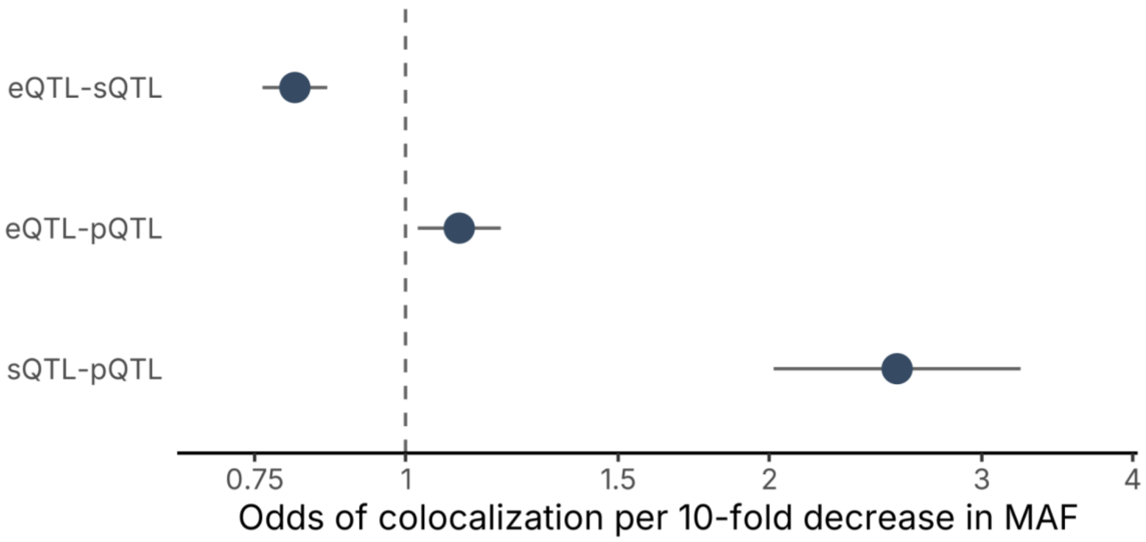


**Supplemental Figure S23. Cross-omic colocalization becomes more likely as allele frequency decreases**

For each pair of molecular phenotypes, points give the odds ratio for a variant being called colocalized per 10-fold decrease in minor allele frequency, with lines showing 95% confidence intervals; the dashed line marks the null of 1, and values above it indicate that rarer variants colocalize more often. The relationship is strongest for sQTL–pQTL (OR 2.55, 95% CI 2.02–3.23, $P = 6.0 \times 10^{-15}$; n = 1,942 variants) and also positive for eQTL–pQTL (OR 1.11, 95% CI 1.02–1.20, P = 0.011; n = 62,744), while eQTL–sQTL shows the opposite direction (OR 0.81, 95% CI 0.76–0.86, $P = 2.1 \times 10^{-11}$; n = 203,599)

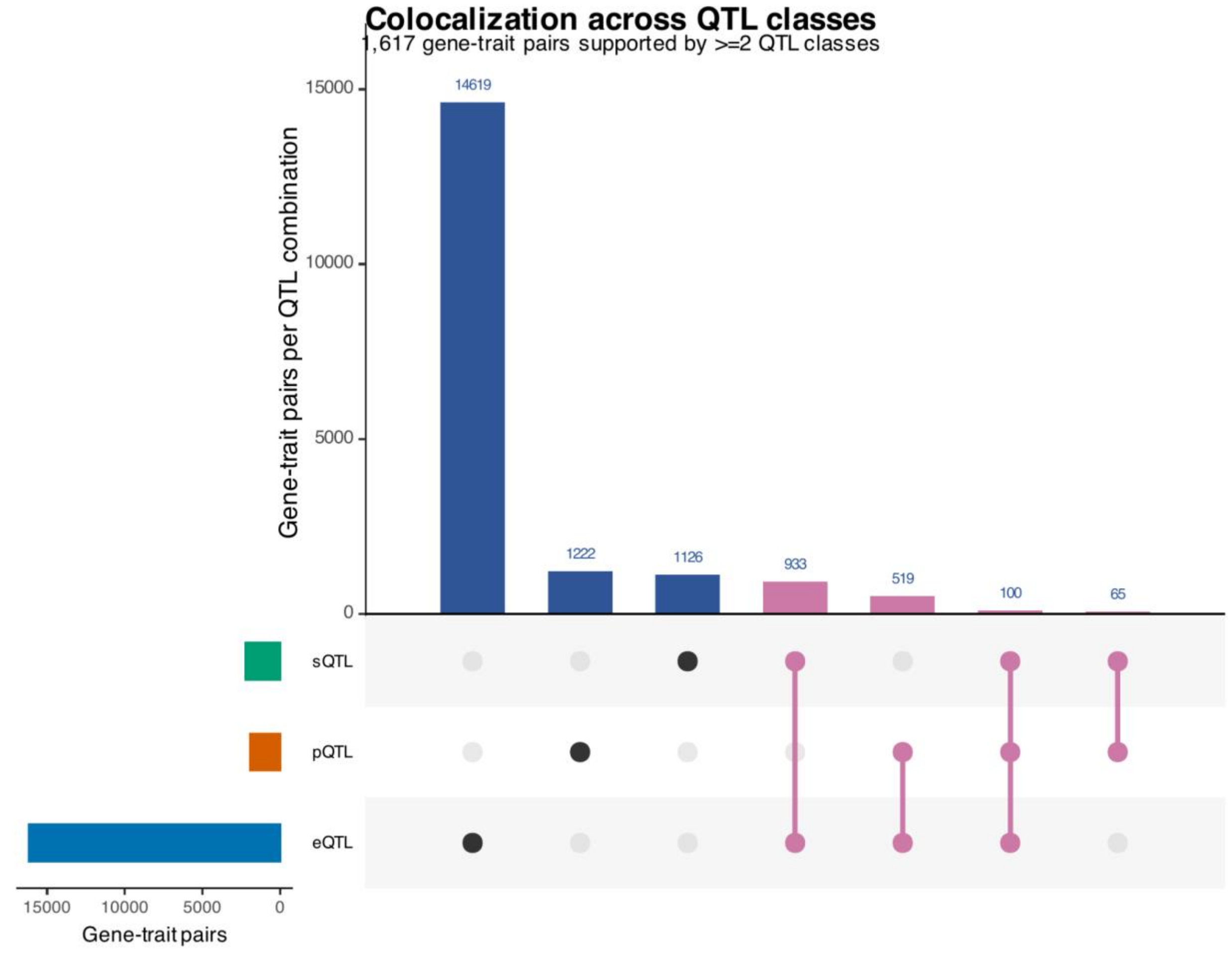


**Supplemental Figure S24. QTL-trait colocalization identifies genes with convergent evidence**

UpSet plot showing the overlap of colocalizing gene–trait pairs across eQTL, pQTL, and sQTL datasets. Each main bar represents an exact QTL-class combination, and the left-hand bars show the total number of gene–trait pairs represented in each molecular layer. Pink bars indicate gene–trait pairs supported by at least two QTL classes. In total, 1,617 unique gene–trait pairs showed multilayer colocalization support, including 100 supported by all three QTL classes. Gene–trait pairs were counted once per QTL class, even if supported by multiple credible sets or loci.

A gene was included if it was supported by at least one of the reported colocalization metrics: FastENLOC RCP ≥ 0.5, CLPP ≥ 0.01 or ≥ 0.05, FastENLOC signal-level RCP Bayesian-FDR support, or gene-level GLCP Bayesian-FDR support.

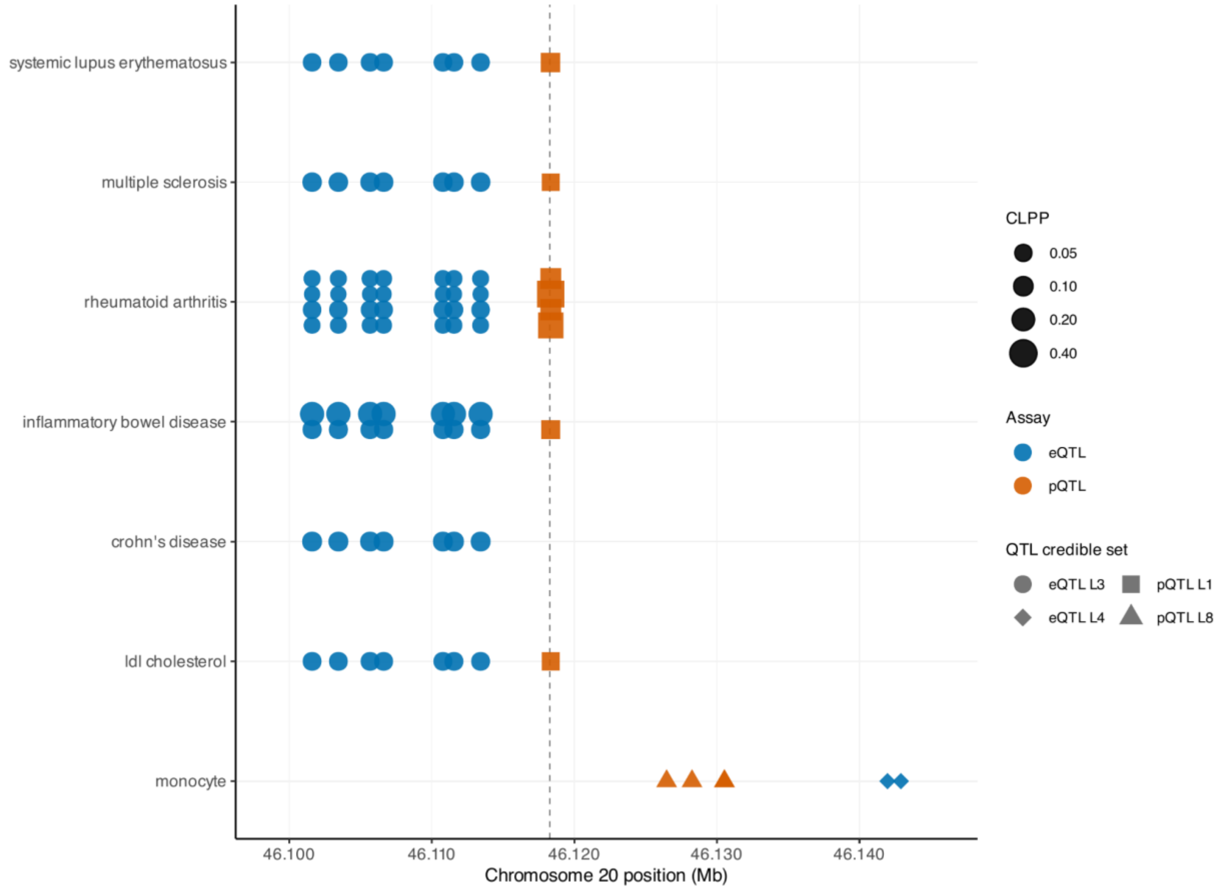


**Supplemental Figure S25. CD40 has convergent evidence for colocalization across QTL types**

Fine-mapped CD40 QTL variants contributing to colocalization signals across seven GWAS traits: systemic lupus erythematosus, multiple sclerosis, rheumatoid arthritis, inflammatory bowel disease, Crohn's disease, LDL cholesterol, and monocyte count. Each row represents a trait, and the x-axis shows chromosome 20 genomic position. Point color denotes molecular assay, point shape denotes the QTL credible set, and point size denotes the signal-level CLPP. If multiple GWAS were tested per trait, the row has multiple rows for each tested GWAS. The dashed vertical line marks the CD40 transcription start site. The displayed signals comprise eQTL credible sets L3 and L4 and pQTL credible sets L1 and L8.

For each CD40 colocalization signal, CLPP was calculated as the sum of the products of GWAS and QTL variant posterior inclusion probabilities across shared variants. The plot includes all fine-mapped variants belonging to QTL credible sets participating in a CD40 signal with CLPP ≥ 0.01; point size reflects the corresponding signal-level CLPP and is therefore shared across variants from the same signal.

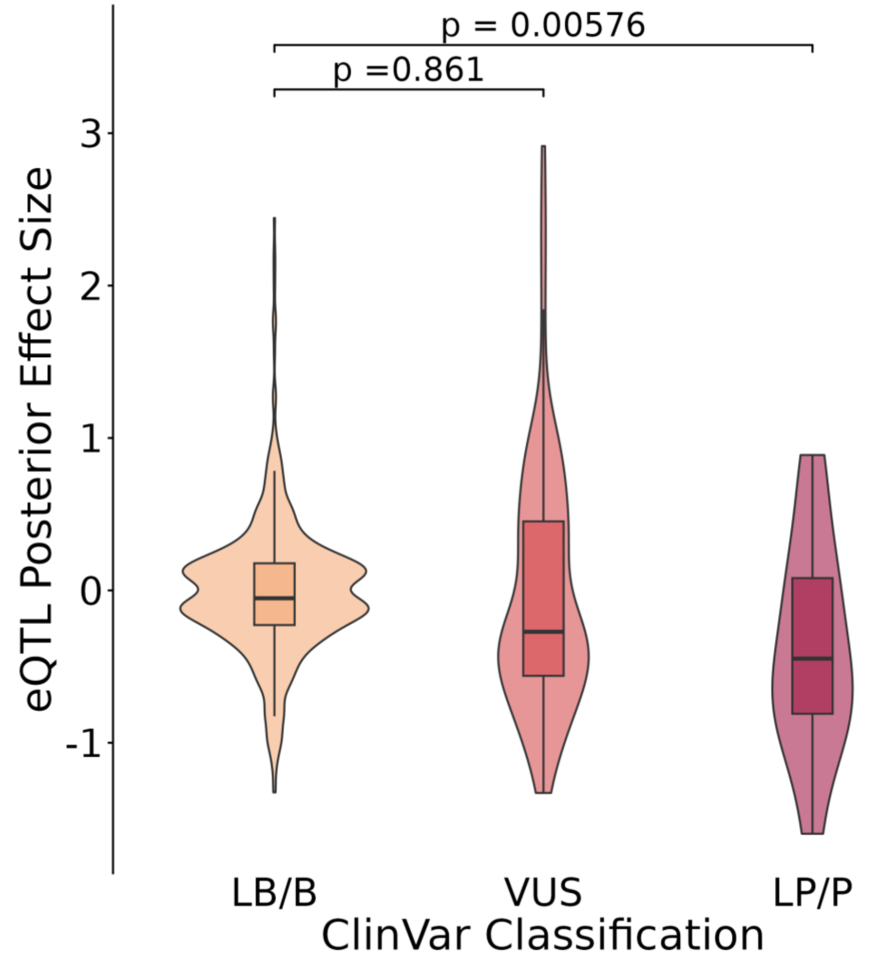


**Supplemental Figure S26. eQTL effects of ClinVar variants**

Distribution of eQTL posterior mean effect sizes for fine-mapped variants that are also present in ClinVar. Each variant was assigned its most severe ClinVar classification across all records, collapsed into three bins: likely benign or benign (LB/B), uncertain significance (VUS), and likely pathogenic or pathogenic (LP/P). Variants classified only as "uncertain risk allele" were excluded. Violins show the kernel density of the posterior mean effect size; overlaid boxplots show the median and interquartile range, with whiskers extending to 1.5 × IQR and outliers omitted. P values are from a linear model of posterior mean effect size on ClinVar class (lm(posterior_mean ~ class)) with LB/B as the reference level. Pathogenic and likely pathogenic variants showed a shift toward reduced expression relative to benign variants ($P = 5.8 \times 10^{-3}$), whereas VUS did not differ from benign ($P = 0.86$).

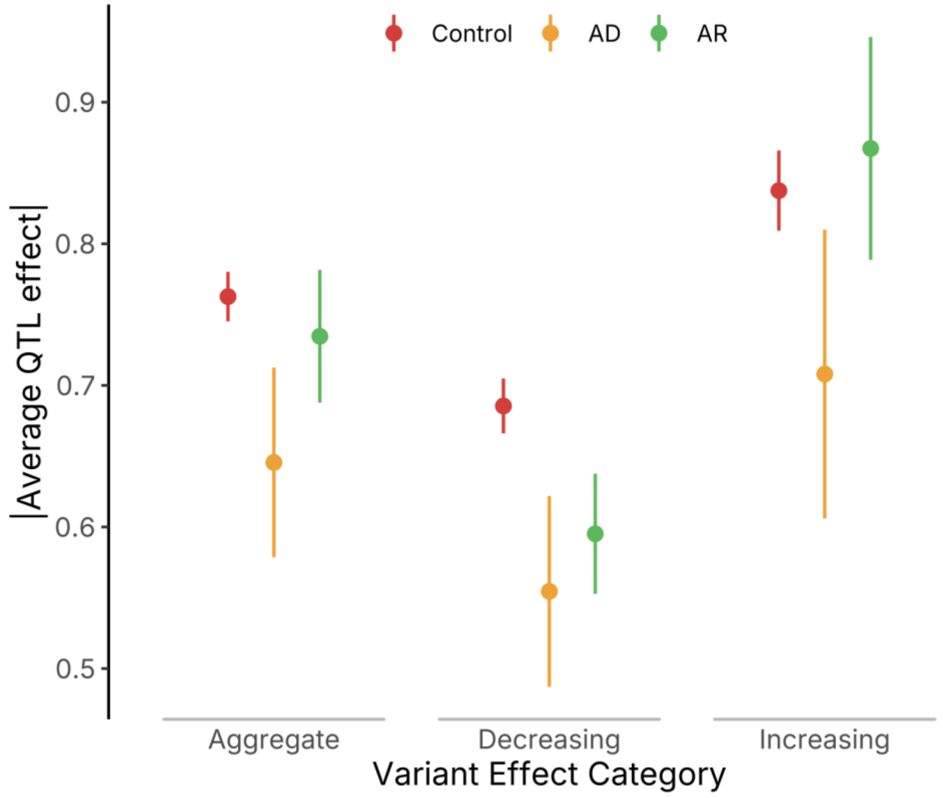


**SUPPLEMENTAL FIGURE S27**

**Disease inheritance pattern by effect size**

**Figure legend**

Mean absolute SuSiE posterior-mean expression-QTL effects for the Control ( sHet < .01 & not in ClinGen) , autosomal dominant (AD), and autosomal recessive (AR) gene groups. Rare variants with posterior inclusion probability greater than 0.9 are summarized in aggregate and after separation by effect direction. The y axis measures absolute posterior-mean effect size. Vertical bars show 95% confidence intervals calculated as the mean plus or minus 1.96 times the standard error, with standard error equal to the sample standard deviation divided by the square root of the group size. Rare fine-mapped eQTLs in autosomal dominant (AD) genes had 15.4% smaller mean absolute effects than controls (adjusted P = 0.0037), whereas autosomal recessive (AR) genes showed no significant overall difference (P = 0.37). Expression-decreasing effects were smaller in both AD and AR genes (19.2% and 13.3% lower; P = 0.0037 and 0.0018, respectively), while expression-increasing effects were smaller in AD genes (P = 0.032) but showed no significant difference in AR genes (P = 0.52).

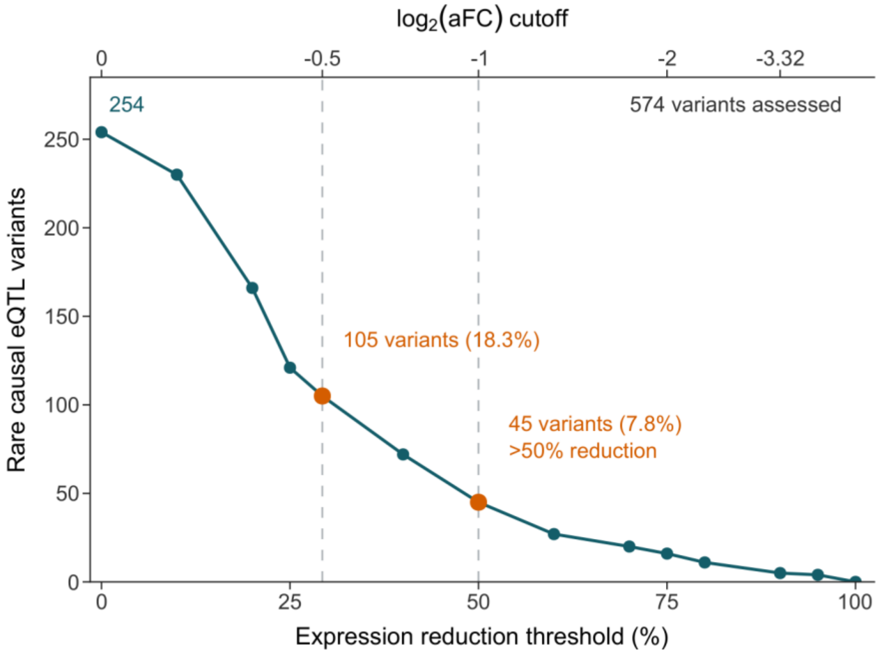


**SUPPLEMENTAL FIGURE S28**

**Rare eQTL variant counts by aFC threshold**

**Figure legend**

The curve shows the number of distinct rare eQTL variants whose estimated allelic expression reduction exceeds each threshold. The analysis includes 574 variants at 453 ClinGen genes with Strong or Definitive disease associations, PIP >0.9, and maximum population allele frequency <1%. Expression reduction is 100*(1 - 2^log2_aFC), relative to the reference allele. Dashed reference lines and orange points mark log2(aFC)=-0.5 (approximately 29.3% reduction; 105 of 574 variants, 18.3%) and log2(aFC)=-1 (50% reduction; 45 of 574 variants, 7.8%).

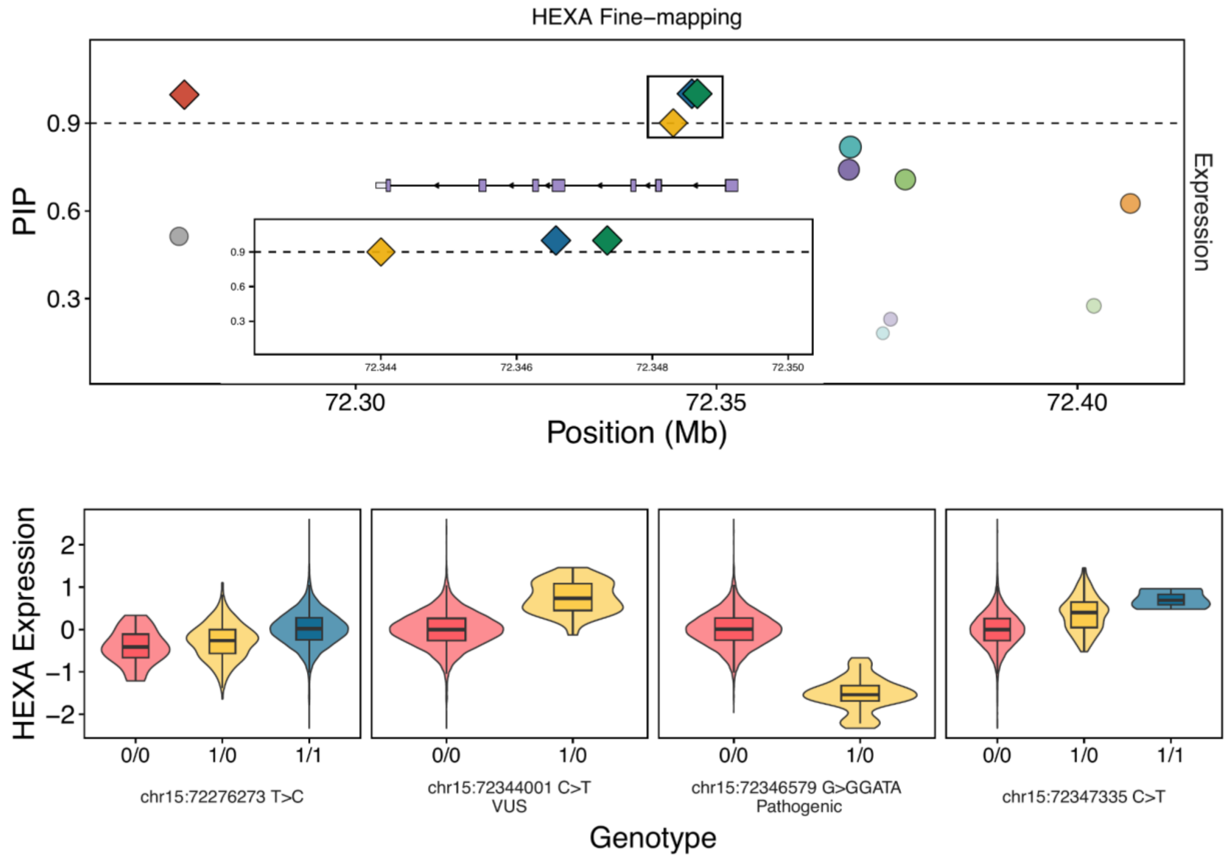


**Supplemental Figure S29. HEXA eQTL fine-mapping**

SuSiE fine-mapping of the cis-eQTL signal for HEXA in the combined-ancestry All of Us expression data. Each point is a variant, plotted by genomic position (x axis, chr15, Mb) against its posterior inclusion probability (PIP, y axis); fill denotes credible-set membership, diamonds are variants with PIP ≥ 0.9 and circles those below, and the dashed horizontal line marks PIP = 0.9. The window shown spans chr15:72.27–72.408 Mb. The black rectangle marks the region expanded in the inset, which shows the HEXA-206 transcript model, exons as light boxes, coding sequence as darker boxes, introns as arrowed lines indicating minus-strand orientation , above the same fine-mapping data restricted to chr15:72.3425–72.350 Mb. (Bottom) Covariate-adjusted HEXA expression by genotype for four fine-mapped variants. For each variant, HEXA expression was regressed on the full set of eQTL covariates with genotype excluded, and the residuals of that model are plotted; individuals are grouped by imputed genotype (0/0, 1/0, 1/1) and those with a missing genotype call were excluded. Violins show the distribution and the overlaid boxplots the median and interquartile range. From left to right the variants are chr15:72276273 T>C, chr15:72344001 C>T (ClinVar variant of uncertain significance), chr15:72346579 G>GGATA (ClinVar pathogenic) and chr15:72347335 C>T; the latter three fall within the region expanded in the inset, while chr15:72276273 lies approximately 70 kb away and corresponds to the leftmost high-PIP variant in the top panel.

Heterozygous carriers of the pathogenic four-base insertion at chr15:72346579 showed reduced HEXA expression relative to non-carriers, consistent with, whereas the other three fine-mapped variants were each associated with increased expression.

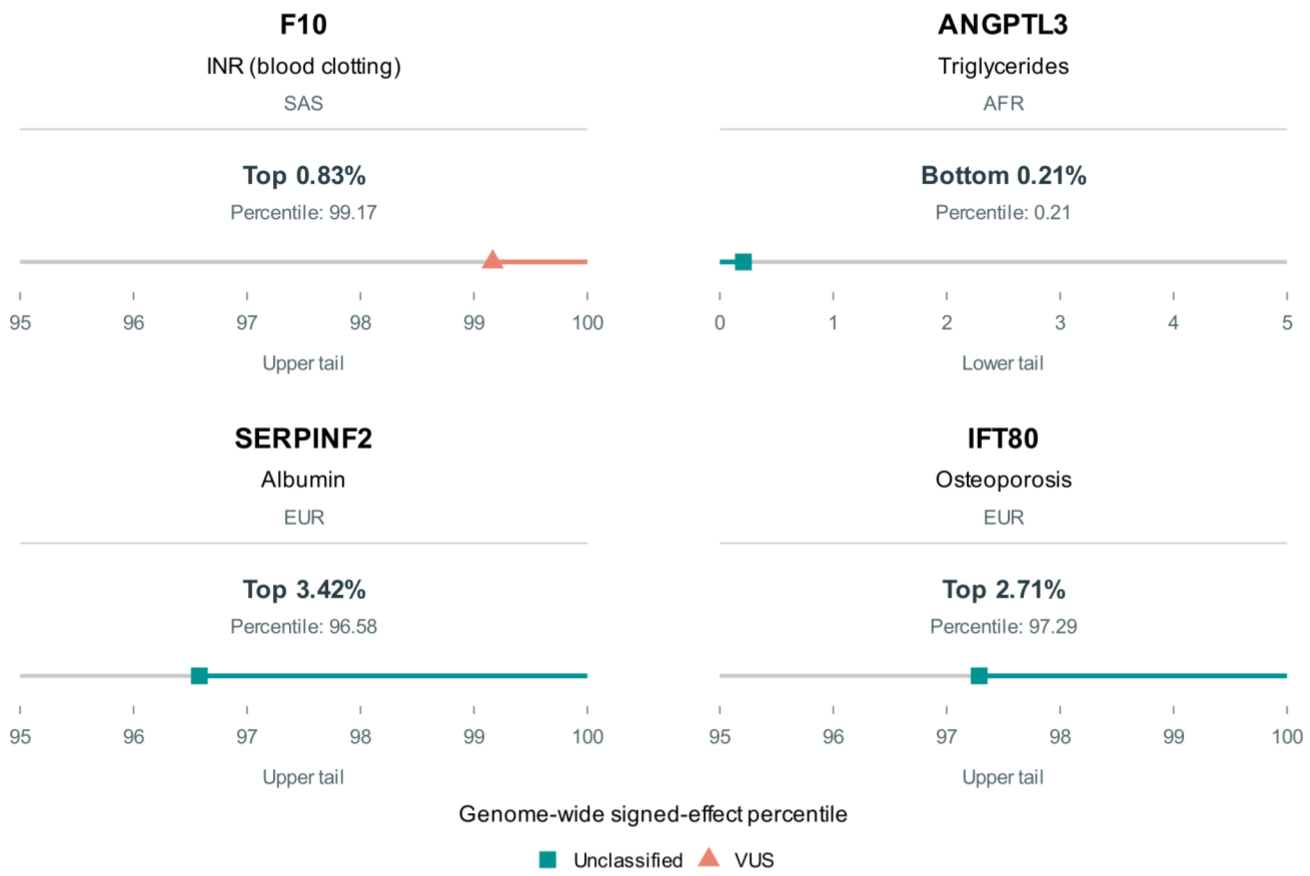


**SUPPLEMENTAL FIGURE S30**

**Trait prioritization for F10, ANGPTL3, SERPINF2, and IFT80**

**Figure legend**

Genome-wide signed-effect percentile ranks for four selected gene-trait examples: F10 with INR (international normalized ratio; blood clotting) in SAS, ANGPTL3 with triglycerides in AFR, SERPINF2 with albumin in EUR, and IFT80 with osteoporosis in EUR. Markers show each signed-effect percentile; colored segments extend from the marker to the tail endpoint. F10, SERPINF2, and IFT80 show the upper 95th-100th percentile range; ANGPTL3 shows the lower 0th-5th percentile range. Upper-tail percentages equal 100 minus the percentile; lower-tail percentages equal the percentile. Orange triangles denote a variant of uncertain significance (VUS); teal squares denote an unclassified variant.

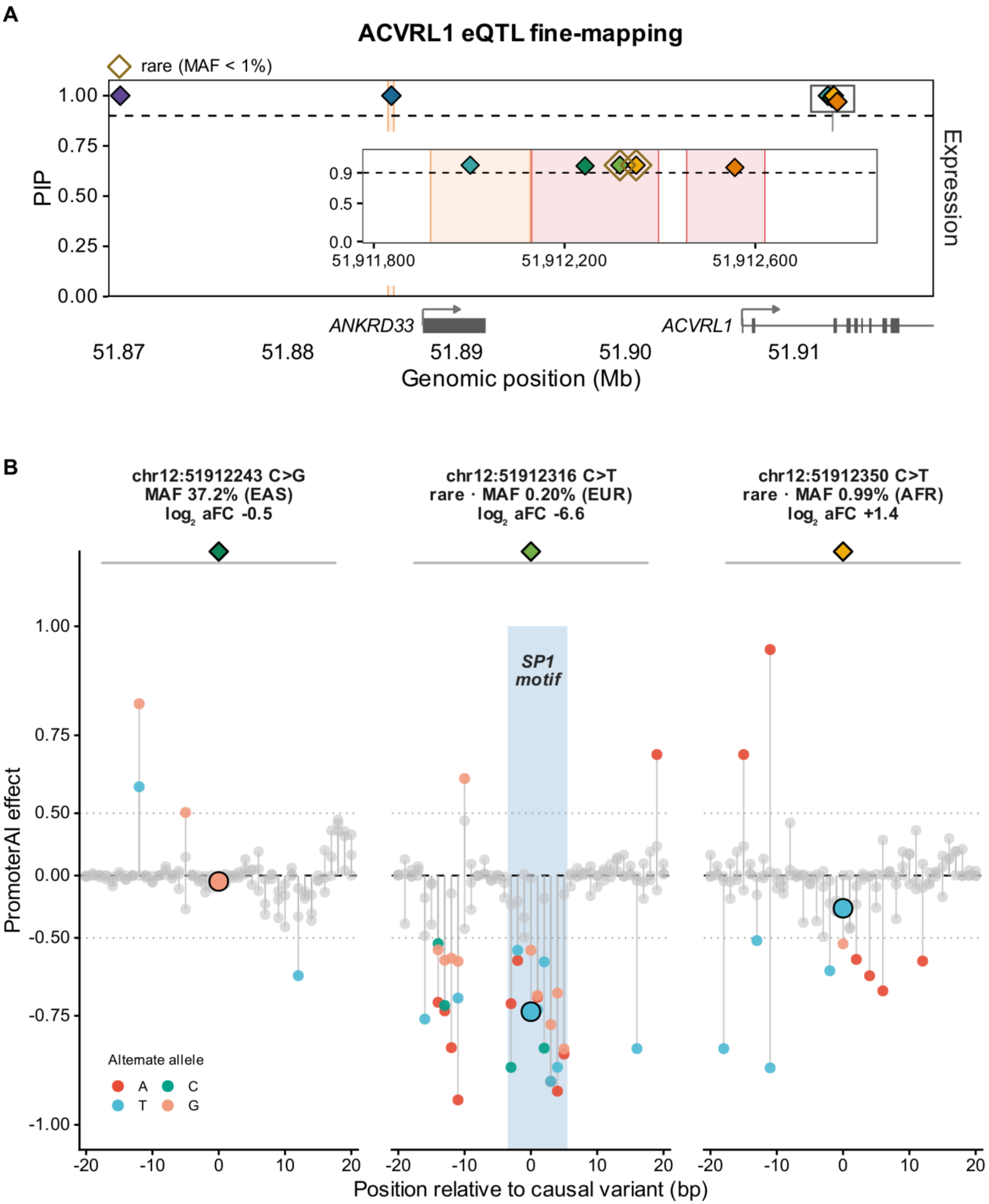
A
ACVRL1 eQTL fine-mapping
rare (MAF < 1%)
PIP
Expression
ANKRD33
ACVRL1
51,911,800
51,912,200
51,912,600
Genomic position (Mb)
B
chr12:51912243 C>G
MAF 37.2% (EAS)
log2 aFC -0.5
chr12:51912316 C>T
rare · MAF 0.20% (EUR)
log2 aFC -6.6
chr12:51912350 C>T
rare · MAF 0.99% (AFR)
log2 aFC +1.4
SP1 motif
PromoterAI effect
Alternate allele
A
C
T
G
Position relative to causal variant (bp)

**SUPPLEMENTAL FIGURE S31 | FIGURE LEGEND**

**ACVRL1 fine-mapping and PromoterAI predictions**

(A) Expression quantitative trait locus (eQTL) fine-mapping at ACVRL1 on chromosome 12 (hg38). Points show variant posterior inclusion probability (PIP) by genomic position, colored by credible set. Diamonds mark PIP at least 0.9; open gold outlines identify rare high-PIP variants (maximum minor allele frequency, MAF, below 1%). The dashed line marks PIP = 0.9. The inset expands the promoter region; shaded bands mark ENCODE candidate cis-regulatory elements. Gene models are shown below.

(B) PromoterAI predictions for ACVRL1 transcript ENST00000419526.6 within 20 bp on each side of chr12:51,912,243 C>G, 51,912,316 C>T, and 51,912,350 C>T. Header text show MAF/ancestry and log2 allelic fold change (aFC): 37.2% EAS/-0.5, 0.20% EUR/-6.6, and 0.99% AFR/+1.4, respectively. Points show predicted effects of single-nucleotide substitutions; those with absolute effect at least 0.5 are colored by alternate base and weaker effects are gray. Outlined circles identify the focal substitutions. The y axis uses the signed-square transform; horizontal guides mark zero and effects of -0.5 and +0.5. Blue shading marks the manually annotated SP1 motif around chr12:51,912,316.

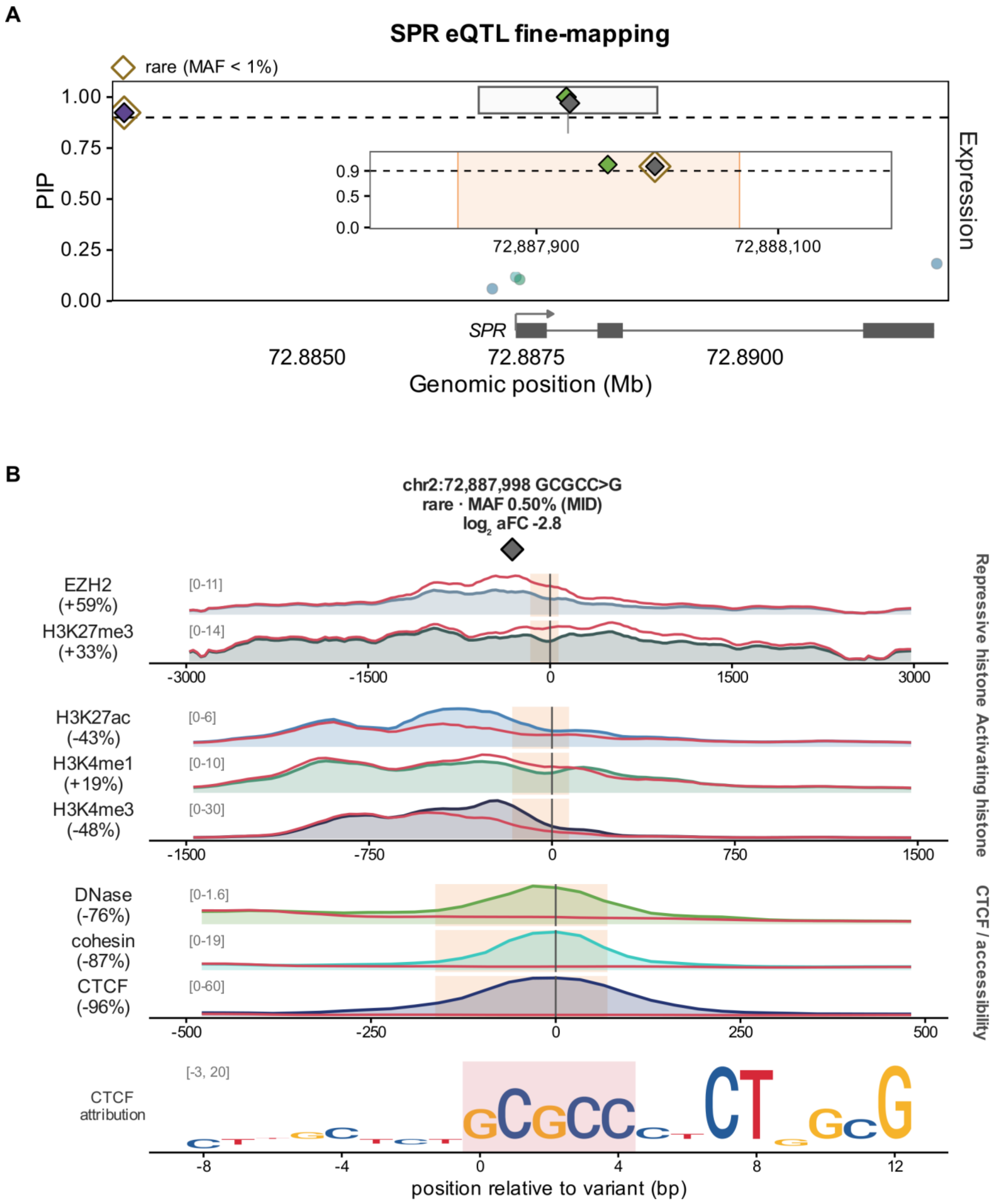
A
SPR eQTL fine-mapping
rare (MAF < 1%)
PIP
1.00
0.75
0.50
0.25
0.00
0.9
0.5
0.0
72,887,900
72,888,100
Expression
SPR
72.8850
72.8875
72.8900
Genomic position (Mb)
B
chr2:72,887,998 GCGCC>G
rare · MAF 0.50% (MID)
log2 aFC -2.8
EZH2 (+59%)
[0-11]
H3K27me3 (+33%)
[0-14]
-3000
-1500
0
1500
3000
Repressive histone
H3K27ac (-43%)
[0-6]
H3K4me1 (+19%)
[0-10]
H3K4me3 (-48%)
[0-30]
-1500
-750
0
750
1500
Activating histone
DNase (-76%)
[0-1.6]
cohesin (-87%)
[0-19]
CTCF (-96%)
[0-60]
-500
-250
0
250
500
CTCF / accessibility
CTCF attribution
[-3, 20]
-8
-4
0
4
8
12
position relative to variant (bp)

**SUPPLEMENTAL FIGURE S32 | FIGURE LEGEND**

**SPR fine-mapping and Borzoi regulatory predictions**

(A) Expression quantitative trait locus (eQTL) fine-mapping at SPR on chromosome 2 (hg38). Points show variant posterior inclusion probability (PIP) by genomic position, colored by credible set. Diamonds mark PIP at least 0.9; open gold outlines identify rare high-PIP variants (maximum minor allele frequency, MAF, below 1%). The dashed line is PIP = 0.9. The inset expands the selected regulatory region; shaded bands mark ENCODE candidate cis-regulatory elements. Gene models are shown below.

(B) Borzoi predictions for chr2:72,887,998 GCGCC>G. The header gives MAF 0.50% in MID and log2 allelic fold change (aFC) -2.8. Colored reference curves and filled areas are compared with red alternate-allele curves. Displayed tracks are EZH2 (+59%), H3K27me3 (+33%), H3K27ac (-43%), H3K4me1 (+19%), H3K4me3 (-48%), DNase (-76%), cohesin (-87%), and CTCF (-96%). Percentage labels give the relative change in mean predicted signal within 96 bp of each reference peak, located within 750 bp of the variant. Only tracks with at least 10% absolute change are retained. Display windows are 3 kb, 1.5 kb, and 500 bp on each side of the variant. Orange shading marks the overlapping ENCODE element; the bottom logo shows CTCF contribution scores with altered bases highlighted.
