## Supplemental Methods for "Multi-omics in the All Of Us Research Program expands mapping genetic mechanisms in health and disease"

### Olink Explore HT processing of AoU samples

#### Sample Receipt

Upon Receipt, plasma samples were stored at -20ºC until processing. These samples only underwent a single freeze thaw event, unless reprocessing was needed.

#### Incoming Sample QC and Source Plate Preparation

Prior to library preparation, 192 samples (consisting of 172 experimental samples and 20 controls) were verified and retrieved from storage and thawed  Controls consisted of several essential control reagents from the Olink Explore HT Controls kit (Olink, Catalog # 98003), including the Olink Explore Sample Control (Olink, Part. No 88011) and the Olink Explore Plate Control (Olink, Part. No 87010), along with the Olink Explore Negative Control (Olink, Part. No 87009). Initial sample transfer from parent matrix racks was performed on an Agilent Bravo liquid handling platform (Agilent Technologies, Model SG2049B001). Samples and control reagents were dispensed into a barcoded Eppendorf 384-well sample source plate. Following plate sealing and centrifugation, the sample plate was stored at +4°C prior to subsequent downstream processing.

#### Sample Dilution

To achieve the target concentrations, automated step-wise dilution of the sample source plate was conducted. Dilution plates were prepared on the Agilent Bravo liquid handling platform (Agilent Technologies) utilizing Olink Target 96/Explore Sample Diluent (Olink, Part. No 84032) in Eppendorf 384-well plates. Automated dilution was performed using an SPT Labtech Mosquito LV liquid handling system (SPT Labtech). The dilution program yielded intermediate 1:10 and 1:100 dilutions in a sequence of automated transfer and pause steps. Intermittent plate sealing and thorough mixing were performed after each dilution transfer on an Eppendorf MixMate (Eppendorf), followed by brief centrifugation. Dilution plates were stored at +4°C and utilized within a strict three-hour window.

#### Incubation Mix Preparation and Hybridization

Incubation mixes were prepared using probes and reagents from the Olink Explore HT Probes kit (Olink, Catalog # 98001), including Olink Explore HT Forward Probes 1–8 (Olink, Part. Nos 88030, 88032, 88034, 88036, 88038, 88040, 88042, 88044), Olink Explore HT Reverse Probes 1–8 (Olink, Part. Nos 88031, 88033, 88035, 88037, 88039, 88041, 88043, 88045), and the Olink Explore Incubation Solution (Olink, Part. No 87004). These probe sets were manually combined in a 96-well midi plate and mixed on an Eppendorf MixMate (Eppendorf). The incubation mixes were reverse-pipetted into a 384-well Eppendorf reagent source plate. Reagent and diluted sample transfers into Eppendorf 384-well incubation plates 1–4 were executed on the SPT Labtech Mosquito LV liquid handler (SPT Labtech). The liquid handling systems transferred 0.6 µL and 0.2µLof sample per well to assemble the hybridization reactions across eight distinct blocks. The incubation plates were immediately sealed and incubated for 20 hours at +4°C to permit target probe hybridization.

#### Index Plate Dispense and PCR Amplification

Following hybridization, the incubation plates were prepared for PCR amplification. Unique barcodes were dispensed from an Olink Index Plate C (Olink, Part. No 88016) (or alternative Olink Index Plates A and B, Olink, Part. No 88016/other) into the incubation plates on the Agilent Bravo liquid handling platform (Agilent Technologies, Model SG2049B001). PCR Master Mix was prepared in a 50 mL Falcon tube using reagents from the Olink Explore HT Sample Prep kit (Olink, Catalog # 98023 or 98009), which included Olink Explore HT PCR Solution (Olink, Part. No 88001), Olink Explore HT PCR Additive (Olink, Part. No 88003), Olink Explore HT PCR Enzyme A (Olink, Part. No 88006), and Olink Explore HT PCR Enzyme B (Olink, Part. No 88007). For samples processed after June 2024, Olink Explore HT PCR Enzyme B (Olink, Part. No 88097) The PCR Master Mix was dispensed into a 96-well midi plate and distributed to the four incubation plates via the Agilent Bravo platform. Sealed and mixed incubation plates were transferred to Eppendorf Mastercycler PCR instruments (Eppendorf) exactly 10 minutes after master mix addition and amplified.

#### Library Pooling and Automated Purification

Amplicons from the four post-PCR incubation plates were pooled into an intermediate 384-well plate and further combined into final 96-well PCR pool plates using the Agilent Bravo platform (Agilent Technologies). To clean and purify the library, automated purification was executed on an Agilent Bravo liquid handling platform integrated with a Benchcel workstation (Agilent Technologies). The automated purification process utilized Beckman Coulter AMPure XP beads (Beckman Coulter), 70% ethanol, and nuclease-free water to perform bead binding, washing, and drying. The purified library blocks were eluted in 45 µL of nuclease-free water and transferred to labeled, clear-capped 2D matrix tubes in a matrix rack.

#### Post-Purification Quality Control

The purified library blocks underwent strict quality control assessment. Size distribution and library integrity were analyzed using the High Sensitivity D1000 ScreenTape Assay (Agilent Technologies) on an Agilent TapeStation instrument (Agilent Technologies) according to the manufacturer's instructions. Purified library blocks (2 µL) were combined with 2 µL of High Sensitivity D1000 Buffer and loaded alongside an electronic ladder. Electropherogram results were analyzed to confirm the presence of a single or double peak around 150 bp, indicating a successful and purified library.

#### Final Pooling, Normalization, and Handoff

To prepare the libraries for sequencing, equal 10 µL aliquots from the eight purified library blocks were combined into a final pooled library tube. This final pooling was performed either manually or automatedly on a Hamilton liquid handler (Hamilton Company) using the WalkUpSeq column pooling protocol. The final pooled library was manually diluted with 745 µL of nuclease-free water to meet the loading specifications of the designated sequencer, the Illumina NovaSeq 6000 S4. For a NovaSeq 6000 S4 run, the library was diluted with 745 µL of nuclease-free water.

#### Watchmaker RNA protocol for AoU samples

Samples were retrieved from freezer storage and thawed over ice. Once thawed, 18µL of sample was transferred directly from the stock tube into the processing plate. Sample only underwent a single freeze thaw event, unless reprocessing was necessary.

#### rRNA and Globin Depletion

Selective depletion of cytoplasmic and mitochondrial ribosomal RNA (rRNA), as well as globin mRNA (specifically HBA1/2, HBB, HBD, HBM, HBG1/2, HBE1, HBQ1, and HBZ), was performed using the Watchmaker Polaris Depletion Kit (Watchmaker Genomics, Catalog # 7K0077-086). For probe hybridization, 18 µL of normalized RNA was combined with 7 µL of Depletion Master Mix containing HMR (Human/Mouse/Rat) Depletion Probes. Thermal cycling for hybridization was executed on an Eppendorf MasterCycler using a profile of 77°C for 2 minutes, followed by 65°C for 15 minutes. Immediately following hybridization, 35 µL of Digestion Master Mix was added to the reaction, and the mixture was incubated at 37°C for 10 minutes to enzymatically degrade the DNA-RNA hybrids.

#### Post-Depletion Cleanup

Following enzymatic digestion, the depleted RNA was purified using AMPure XP SPRI beads (Beckman Coulter, Catalog # A63881) at a 1.8x volume over volume (v/v) ratio. The beads were washed twice with 70% ethanol, followed by a 5-minute air dry at ambient temperature. 27 uL of fragmentation and priming buffer was added to the samples  and the reaction was incubated at 80°C for 5 minutes followed by a 12°C hold

#### cDNA Synthesis

First-strand cDNA synthesis was initiated by adding 10 µL of First-Strand Master Mix from the Watchmaker RNA Library Prep Kit (Watchmaker Genomics, Catalog # 7K0078-096). Reverse transcription was performed on a thermal cycler using a program of 25°C for 10 minutes, 42°C for 15 minutes, and 70°C for 15 minutes. Following first-strand synthesis, second-strand synthesis and dATP tailing (A-tailing) were performed. A 15 µL aliquot of Second-Strand/A-Tailing Master Mix was added to the first-strand reaction. Complementary second-strand synthesis was carried out at 42°C for 5 minutes, followed immediately by dATP tailing at 62°C for 10 minutes.

#### Adapter Ligation and Indexing

Adapter ligation was performed by adding Duplex Unique Molecular Identifiers (UMIs; Integrated DNA Technologies) and Watchmaker Ligation Master Mix (Watchmaker Genomics) to the A-tailed cDNA, followed by incubation at 20°C for 15 minutes. Post-ligation purification was carried out using a 0.7x v/v SPRI bead ratio to remove adapter dimers, with final elution in 22 µL of 10 mM Tris-HCl. For library amplification, unique 8-base dual-index sequences (P5 and P7 primers; Integrated DNA Technologies) and Equinox PCR Amplification Master Mix were added to the purified adapter-ligated cDNA. Thermal cycling was performed using the following profile:

an initial denaturation at 98°C for 45 seconds, followed by 10 cycles of 98°C for 15 seconds, 55°C for 30 seconds, and 72°C for 30 seconds.

#### Final Library Purification and Quality Control

Amplified libraries underwent a final purification using a 0.7x v/v SPRI bead ratio and were eluted in 10 mM Tris-HCl. Final library concentration and yield were determined via PicoGreen fluorometric analysis using the Invitrogen Quant-It Broad Range dsDNA Kit (Invitrogen, Catalog # Q33130) on a Molecular Devices Gemini XPS plate reader.

#### Final Library Normalization, Pooling, and Sequencing Preparation

The final purified libraries were normalized to a concentration of 2 ng/uL using 10 mM Tris-HCl (pH 8.0) on a Hamilton STARlet liquid handler, based on an assumed average library fragment size of 400 bp. To verify and confirm molarity, quantitative PCR (qPCR) was performed on a QuantStudio 7 system. Validated libraries were then pooled into sets of up to 94-plex. Prior to sequencing handoff, the final pools were verified to be within a concentration range of 1–3 nM. All library construction, cleanup, and quality control steps were automated on the Agilent Bravo liquid handling platform.
